# A rapid review of the effectiveness of interventions for addressing digital exclusion in older adults

**DOI:** 10.1101/2024.03.21.24304670

**Authors:** Alesha Wale, Jordan Everitt, Toby Ayres, Chukwudi Okolie, Helen Morgan, Hannah Shaw, Rhiannon Tudor Edwards, Jacob Davies, Ruth Lewis, Alison Cooper, Adrian Edwards

**Affiliations:** Public Health Wales, United Kingdom; Centre for Health Economics and Medicines Evaluation, Bangor University, United Kingdom; Health and Care Research Wales Evidence Centre, Bangor University, United Kingdom; Health and Care Research Wales Evidence Centre, Cardiff University, United Kingdom

## Abstract

Older adults constitute the largest proportion of non-users of the internet. With the increasing digitalisation of services, in particular those provided in Social Care in Wales, it is important to understand how best to support older adults to overcome the challenges they face with accessing or engaging with the digital world (for personal use). This rapid review aimed to assess the effectiveness of interventions to address digital exclusion in older adults (aged 60 years and above). Digital exclusion can occur due to issues with motivation (if people do not see why the internet might be beneficial), accessibility (unable to physically access to the internet), ability (lack of skills to use the internet) or affordability (unable to afford access to the internet) of digital technology.

**Research Implications and Evidence Gaps:** The majority of studies included in this review were of low quality. It is unclear whether study findings would be generalisable to the UK. Outcome measures were heterogeneous across studies making it difficult to compare findings directly. Only one study assessed the cost-effectiveness of a digital education intervention. No study reported on interventions to address language barriers, for example, that may be experienced by people whose first language is not English. No study focused specifically on interventions to improve access to, or affordability of the internet and digital technologies to overcome digital exclusion. Further high-quality UK-based research is needed to better understand the effectiveness and cost-effectiveness of interventions for addressing digital exclusion in older adults.

**Policy and Practice Implications:** This rapid review highlighted the **potential benefits of a range of complex multi-component educational interventions**, particularly with regards to improving digital literacy, and suggests that older adults are accepting of these interventions. To reduce digital exclusion in older adults, evidence suggests it may be important to ensure structural barriers, such as access to the internet and affordability of devices are removed. However, the cost of provision should be considered and assessed. Educational interventions may help to reduce perceptual barriers regarding digital technologies that contribute to digital exclusion including lack of confidence, fear and anxiety, or perceived lack of abilities. It is important to consider that older adults should be equipped with the skills to make an informed choice to interact with essential services physically (offline) or digitally. With the increasing digitalisation of services, it is important that older members of the community who do not wish to use digital technologies, are not left behind or disadvantaged.

**Funding statement:** Public Health Wales were funded for this work by the Health and Care Research Wales Evidence Centre, itself funded by Health and Care Research Wales on behalf of Welsh Government.

## 1. BACKGROUND

### 1.1 Who is this review for?

This Rapid Review was conducted as part of the Health and Care Research Wales Evidence Centre Work Programme. The above question was formulated by Social Care Wales.

### 1.2 Background and purpose of this review

Digital exclusion refers to when sections of the population are unable to exploit the benefits that using digital technologies might make available to them (Honeyman 2020). This occurs due to issues with motivation (those who do not see why the internet might be beneficial) (NIHR 2022), accessibility (those who physically do not have access to the internet), ability (those who do not have the skills to engage with an online environment) or affordability (those who cannot afford access to the internet) of digital technology (Ofcom 2022). Digital divide refers to the gap between those who are excluded and those who are able to benefit from technology (Honeyman 2020). Digital inclusion is an approach for overcoming the barriers to opportunity, access, knowledge and skills for using technology (Gann 2018).

In Wales, 7% of the population (over 16 years) are not online (Welsh Government 2023a). A lack of digital skills and access in terms of device and connectivity can lead to poorer health outcomes, increased loneliness and social isolation, as well as less access to information, learning and essential services (Good Things Foundation 2024). Older adults constitute the largest proportion of non-users of the internet worldwide (Lu et al 2022). While internet use is increasing among older adults, they remain a group at risk of being digitally excluded, with older adults being more likely to lack confidence online (Ofcom 2023). In Wales between 2022-2023 32% of adults aged 75 years and over reported they were not using the internet with 29% not having internet access in their household (Welsh Government 2023b).

Older adults may face a range of barriers when considering or attempting to use digital technologies. Physical barriers include aging-related barriers such as poor eyesight and lack of dexterity, and individual/personal barriers such as living alone, lower income, lack of knowledge around how to use digital technologies and difficulty understanding digital terminology (Vassilakopoulou 2023; Moroney 2020; Schirmer 2023, Yazdani-Darki et al 2020). Perceptual barriers include internalised negative perceptions and stereotypes of aging, fear and anxiety, and lack of confidence of using and/or in digital technologies (Gates 2022; Vassilakopoulou 2023; Schirmer 2023). This highlights the need to support older adults in developing the necessary skills required to navigate the rapidly evolving digital landscape and address the current digital divide.

Preliminary work focused on exploring the effectiveness of interventions to support older adults accessing social care services online. With the increasing digitalisation of services, it is important to understand how best to support older adults to overcome the challenges they face with accessing or engaging with the digital world. However, the initial searches did not identify any studies specifically aimed at supporting older adults to access social care services online. As such, it was agreed with stakeholders that the scope of this rapid review would be broadened to explore the effectiveness of interventions to support older adults to access and engage with digital technologies for personal use. Increasing digital inclusion may help to reduce inequalities and ensure equal access to these services (King’s fund 2023). Our work aims to support this by identifying interventions that are effective at reducing digital exclusion among the older aged population.

This rapid review focuses on primary studies assessing the effectiveness of interventions aimed at addressing digital exclusion in older adults (aged 60 years or over). Interventions aimed at supporting access to health-related services or improving work-related digital skills were excluded from this RR. There are various ways to measure digital literacy that are used in research. For the purposes of this review, digital literacy is defined as the capabilities that allow someone to live, learn, work, participate and thrive in a digital society (NHS Health Education England 2016).

## 2. RESULTS

### 2.1 Overview of the Evidence Base

A total of 21 comparative primary studies were included in the review (see section 5.1, Table 4 for full eligibility criteria). Study designs were either randomised controlled trials (RCT) (n=3) or quasi-experimental (n=18). Of the 18 quasi-experimental studies, eight were uncontrolled before and after (pre-post) studies while 10 were non-randomised controlled studies. Included studies were conducted in a range of countries including USA (n=6), South Korea (n=3), Canada (n=2), Mexico (n=2), Australia (n=1), China (n=1), Netherlands (n=1), Peru (n=1), Portugal (n=1), Singapore (n=1), and Spain (n=1). One study was conducted across multiple countries including the UK, Latvia, Poland and Portugal. Sample sizes were generally small, ranging between 5 and 381 participants in total. Seven of the studies included more than 100 participants.

Data collection methods utilised self-report methods such as surveys and questionnaires, but also included quizzes, focus groups and interviews. The majority of data collection methods included validated tools and measures.

Many of the studies contained complex multi-component interventions, and often had overlapping features making it difficult to categorise the interventions for synthesis. We grouped the studies based on some of the more novel features that could be incorporated or considered when designing interventions to address digital exclusion in the future. This included studies that contained an intergenerational component (n=4), studies that were incorporated into existing services (n=3), studies that created tailored computer software (n=2), one study that incorporated an online game into the intervention, and one study that specifically aimed to teach participants to detect online deception. The remaining 10 studies were classified as more traditional educational interventions. A range of outcomes were reported including digital literacy, technology use and adoption, confidence and self-reported independence, acceptability, and cost-effectiveness. Digital literacy was assessed independently as an outcome, but also included proxy measures for digital literacy including computer proficiency, mobile device proficiency, digital competence, online deception detection and E-health literacy.

The methodological quality of included studies was assessed using the appropriate Joanna Briggs Institute (JBI) critical appraisal tool (for quasi-experimental studies and RCTs). The three RCTs were judged to be of moderate quality while all 18 quasi-experimental studies were determined to be of low quality due to the absence of a control group in eight studies, and uncertainty regarding the reliability of outcome measures used in 13 studies. Of the non-randomised controlled studies, three did not report any between-group analysis and only provided within-group differences (pre-post). The output of the quality assessment can be seen in sections 5.6 and 6.3.

#### Intervention components

The majority of interventions included were diverse, complex and multi-component. All were educational, aimed at training older adults to improve their digital literacy skills. Many incorporated innovative elements such as utilising existing services to deliver the intervention, involving gamification and tailoring the intervention to the specific needs of the participant. Others were single component, focussing on digital skills education more broadly. The various elements and components utilised within each intervention can be seen in the matrix (Table 1). Details of the interventions can be seen in Table 2; a summary of the findings can be found in Table 3 and a summary of the included studies can be seen in section 6.2 (Table 5).

**Table 1.**
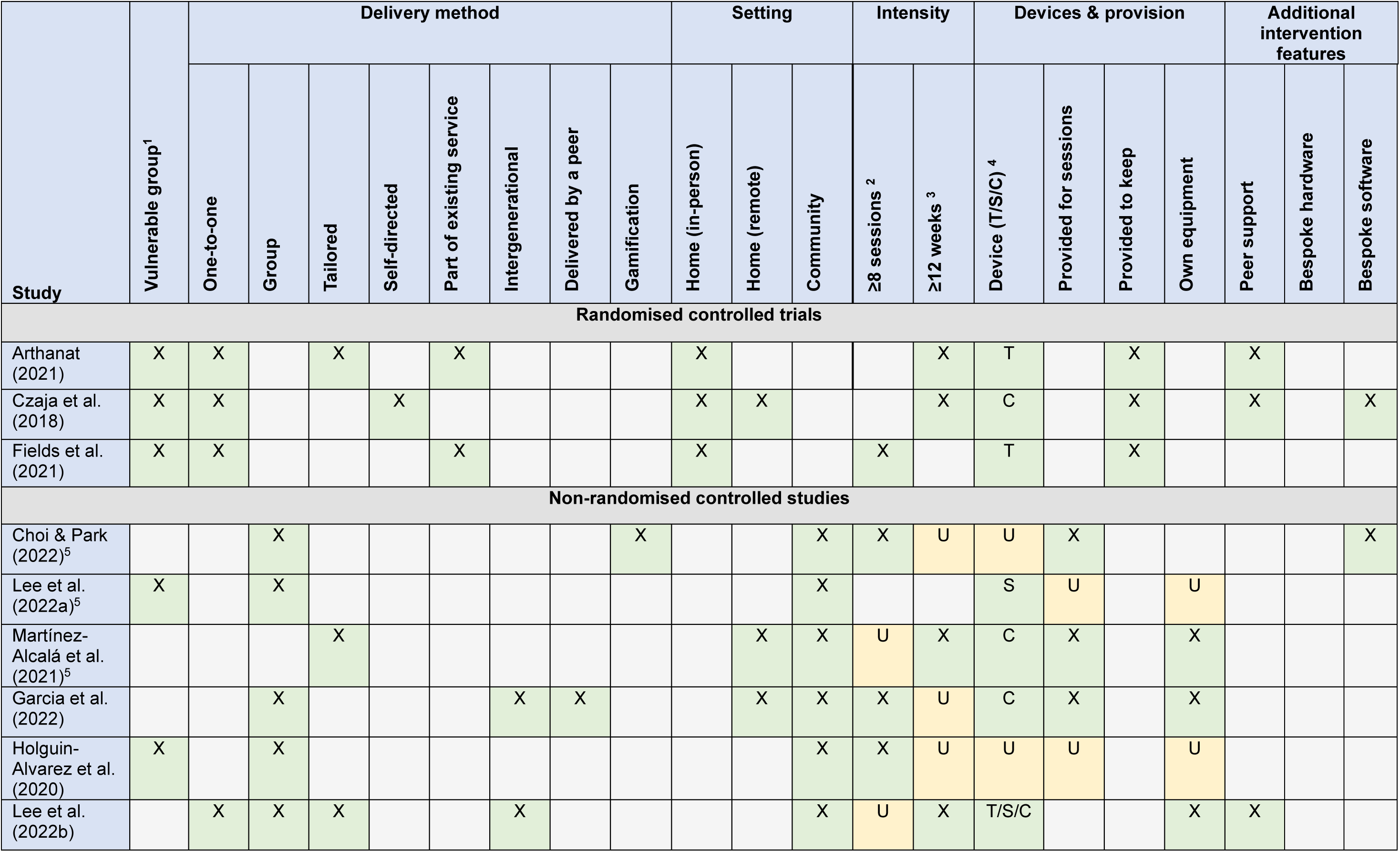
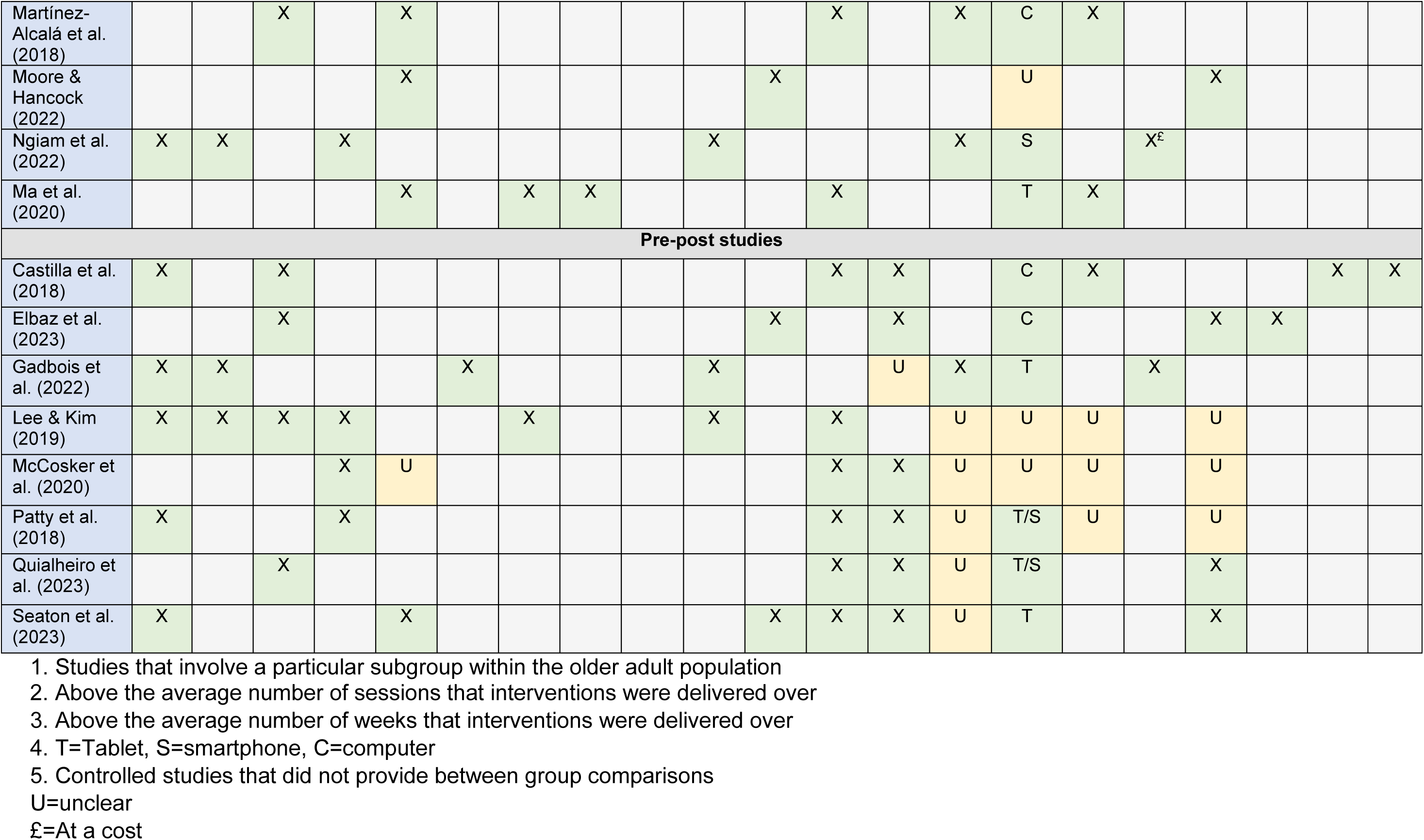
Interventions matrix.

Of the 21 included studies, eleven focused on evaluating interventions among specific subgroups, such as older adults from small or rural towns (n=4), older adults of low socio-economic status (SES) (n=3), older adults at risk of social isolation (n=2), older adults who are homebound (n=1), and visually impaired older adults (n=1).

Interventions were commonly conducted solely within the community (public settings) (n=10), conducted solely in-person at the participants’ homes (n=7) either held virtually (n=2), on a one-to-one in-person (n=4) basis, or a mixture of both (n=1). Three studies included interventions that were conducted across both home and community settings and one home-based intervention involved a mix of in-person and remote teaching. Study authors indicated home-based training may be deemed to be most meaningful and an anxiety free space to learn in. It was also considered convenient in terms of scheduling and transportation (Arthanat et al., 2022). Six interventions were self-directed and the remaining involved a tutor of some sort.

Seven interventions were tailored to participant needs, by using an individualised or tiered curriculum approach, enabling those with varying baseline skills to be taught at an appropriate level to ensure they could gain the most out of the intervention (Ngiam et al., 2022). Patty et al. (2018) also varied the duration of the intervention to ensure all participants completed the programme, whilst accounting for the differing capabilities of participants. Six interventions involved personal computer (PC) equipment, five involved tablet devices, two involved smartphones and three involved the use of both tablet devices and smartphones and five did not specify a device. Five interventions enabled participants to retain digital equipment indefinitely after the intervention had finished, one of which required participants to pay for the device and internet access (Ngiam et al., 2022). Devices provided included tablets in three studies, smartphones in one study and PCs in one study. Seven interventions involved the use of participants own equipment.

### 2.2 Effectiveness of educational interventions on digital technology use

A total of five studies explored the effect of educational interventions on the adoption or use of digital technology (Arthanat., 2021; Fields et al., 2021; Gadbois et al., 2022; Lee et al., 2022a; Seaton., 2023). All studies explored intervention effectiveness among vulnerable population group including those living in rural areas (n=3), those who are socially isolated, (n=1) or homebound (n=1). This outcome was measured using a pre-post study design in two studies (Gadbois et al., 2022; Seaton et al., 2023), a non-randomised controlled study design in one study (Lee et al., 2022a) and a RCT study design in two studies (Arthanat., 2021; Fields et al., 2021). Study findings appeared to show effects in favour of the interventions.

#### Interventions incorporated into existing services

- Gadbois et al. (2022) conducted a pilot study to assess the impact of the Talking Tech intervention, a 14-week, one-to-one, home-based, technology training intervention on technology use. The intervention was incorporated into a home-delivered meals programme for homebound older adults. **A trend towards increased technology use (measuring 10 activities) was reported after the intervention, however the increases were not statistically significant.** Out of 18 participants **who completed baseline and follow-up surveys,** seven (38.89%) reported increased internet use for activities including shopping, prescriptions, social media, and health-related activities, while three (16.67%) reported less use and eight (44.44%) participants reported the same use after the intervention had concluded.
- Arthanat (2021) conducted an RCT to assess the effectiveness of a home-based, individualised inter-generational ICT training programme called the Individualised Community and Home-based Access to Technology Training (i-CHATT) on technology use (measuring 56 activities related to use) for older adults living in rural towns. The intervention formed part of an occupational therapy programme. While no significant change in the frequency of ICT use was reported at the six-month follow-up, **technology use in the intervention group sustained an increasing trend during the remaining follow-up points (12, 18, and 24 months) ending at 36 activities each month compared to about 30 by the control group.**
- Fields et al. (2021) conducted an RCT to assess the impact of a one-to-one, home technology training intervention on technology use for socially isolated older adults. Participants were provided with a tablet and internet access. The intervention was incorporated into a volunteer-based programme that provides home visits for lonely older adults. **Technology use was found to increase statistically significantly in the intervention group between baseline and two months post intervention** (baseline: 33% no internet or email use, two-month: 0% no internet or email use, p=0.004). **There was no change over time within the control group** (baseline: 53% no internet or email use, two-month: 60% no internet or email use, p=0.63). **There were significantly higher rates of technology use within the intervention group compared to the waitlist arm** (OR: 91.20; 95% CI: 11.02 to +Infinity; no p value given).

#### Traditional digital literacy interventions

- Seaton et al. (2023) explored the impact of the Gluu Essentials digital skills training programme on technology use in older adults living in rural communities. **The results showed that the frequency of going online for shopping (p=0.01) and accessing government services (p=0.02) statistically significantly increased after the intervention. Technology use for accessing online banking (p=0.10), and emergency services (p=0.42) also increased after the intervention, although these improvements were not statistically significant. No differences were found in how frequently technology was used to send and receive emails (p=0.47), or searching for information (p=0.96) after the intervention. However, the frequency of going online for COVID-19 related information statistically significantly decreased after the intervention (p=0.01)**. According to the study authors, this may have been due to the easing of COVID-19 restrictions in Canada towards the end of the study period.
- Lee et al. (2022a) assessed the impact of a six-session, community-based, digital literacy education programme to increase the use of smartphones in older adults living in rural areas**. No significant increases were reported in taking photos for either group after the intervention period** (experimental group: p=0.087; control group: p=0.176). **However, the frequency of phone calls made using smartphones increased significantly in the intervention group by 8.5% after the intervention** (t: 1.934, p=0.026). **No changes were observed in the control group. A statistically significant increase in ability to video record using a smartphone was also found in the intervention group** (t/x2: 4.493, p=0.049). **Similarly, no improvement was found in the control group** (t/x2: 0.522, p=0.527). No between group differences were reported.

#### 2.2.1 Bottom line results for effectiveness of educational interventions on digital technology use

There is evidence to suggest that educational interventions are effective at improving digital technology use in older adults from vulnerable contexts including those living in rural areas, those who are socially isolated, or homebound. However, the evidence is mostly of low quality. Evidence suggests that uptake can be increased using traditional digital literacy interventions (n=2) and interventions that are incorporated into existing services (n=3).

While each study reported some outcome improvements for the intervention group, not all were statistically significant when compared to baseline or a comparator. Various forms of technology use were assessed among the studies (such as smartphone use, accessing services such as banking and shopping), but not all were found to improve following the intervention. However, this could be due to the individual digital needs of the participants, rather than the intervention itself.

### 2.3 Effectiveness of educational interventions on digital literacy

Fifteen studies explored the impact of a range of interventions on digital literacy (Choi and Park., 2022; Czaja et al., 2018; Elbaz et al., 2023; Garcia et al., 2022; Holguin-Alvarez et al., 2020; Lee and Kim., 2019; Lee et al., 2022b; Martínez-Alcalá et al., 2018; Martínez-Alcalá et al., 2021; McCosker et al., 2020; Moore and Hancock., 2022; Ngiam et al., 2022; Patty et al., 2018; Quialheiro et al., 2023; Seaton et al., 2023). Three focused on digital literacy as a standalone outcome (Choi and Park., 2022; Garcia et al., 2022; Martínez-Alcalá et al., 2021; Ngiam et al., 2022), two on E-health literacy (Lee and Kim., 2019; Lee et al., 2022b), and ten used proxies for digital literacy (Czaja et al., 2018; Elbaz et al., 2023; Holguin-Alvarez et al., 2020; Martínez-Alcalá et al., 2018; McCosker et al., 2020; Moore and Hancock., 2022; Patty et al., 2018; Quialheiro et al., 2023; Seaton et al., 2023). Study findings appeared to show positive effects in favour of the interventions.

#### Effectiveness of educational interventions on digital literacy as a standalone outcome

Digital literacy was assessed as a standalone outcome in three studies using a non-randomised controlled study design (Choi and Park., 2022; Martínez-Alcalá et al., 2021; Ngiam et al., 2022). A broad range of questionnaires and tools were used to measure digital literacy. See data extraction in section 6.2 (Table 5) for full details of the tools used.

#### Gamification

- Choi and Park. (2022) assessed the impact of a 10-session, community-based, educational IT programme using an educational game called ‘Save the Titanic using the decision tree’, compared with general IT educational sessions. No between group differences were reported. However, within group differences identified **digital literacy statistically significantly improved in the intervention group (p<0.05) after receiving the intervention and while digital literacy also improved in the control group** (after receiving general internet and digital use education)**, this improvement was not statistically significant (p>0.05).** More specifically, when looking at recognition and behaviour domains the intervention group statistically significantly improved after the intervention (mean recognition score 2.45, SD:0.55 vs 3.02, SD:0.64; p=0.012; mean behaviour score 3.16, SD:0.66 vs 3.67 SD:0.59; p=0.001). Improvements were also made in the control group however these were not statistically significant (mean recognition score 2.34, SD:0.47 vs 2.51, SD:0.35; p=0.102; mean behaviour score 3.10, SD:0.81 vs 3.32, SD:0.34; p=0.84).

#### Traditional digital literacy interventions

- Martínez-Alcalá et al. (2021) assessed the impact of digital literacy workshops delivered in different formats in response to the COVID-19 pandemic. This study contained three different delivery methods as the intervention moved from blended learning (pre-pandemic), to transition treatment, and then a fully digital intervention. The study analysed data from two groups of older adults with varying levels of baseline digital literacy (basic and intermediate) as they progressed through the workshops and workshop formats. **Both groups, regardless of initial skill level, exhibited statistically significant improvements in digital literacy scores (p<0.001) across all delivery formats**. No between group differences were reported.
- Ngiam et al. (2022) explored the impact of a home-based, volunteer-led, one-to-one digital literacy programme, compared with no intervention, in older adults of lower SES. Older adults were equipped with smartphones and internet connection. The study findings showed **statistically significant improvements in mean digital literacy scores of the intervention group after the programme, compared to the control group** (2.42, 95% CI: 1.71 to 3.11 vs 0.13 95% CI: −0.48 to 0.75 mean difference: 2.28, 95% CI: 1.37 to 3.20; p<0.001). This difference remained after adjusting for baseline digital literacy scores and differences in age, gender, education, living arrangement, housing type and baseline social connectivity and loneliness status (*β:* 1.90, 95% CI: 0.91 to 2.90, p<0.001).

#### Effectiveness of educational interventions on proxy outcomes for digital literacy

Ten studies assessed the impact of interventions on a number of proxy outcomes for digital literacy (computer/mobile device proficiency, digital competence, digital/ICT skills and online deception detection) (Czaja et al., 2018; Elbaz et al., 2023; Garcia et al., 2022; Holguin-Alvarez et al., 2020; Martínez-Alcalá et al., 2018; McCosker et al., 2020; Moore and Hancock., 2022; Patty et al., 2018; Quialheiro et al., 2023; Seaton et al., 2023). This outcome was measured using a pre-post study design in five studies (Elbaz et al., 2023; McCosker et al., 2020; Patty et al., 2018; Quialheiro et al., 2023; Seaton et al., 2023), a non-randomised controlled study design in four studies (Garcia et al., 2022; Holguin-Alvarez et al., 2020; Martínez-Alcalá et al., 2018; Moore and Hancock., 2022) and a RCT study design in one study (Czaja et al., 2018). Digital literacy proxy outcomes were measured using a range of self-reporting tools including the Computer Proficiency Questionnaire (CPQ) and the Mobile Device Proficiency Questionnaire (MDPQ). See section 6.2 for full details.

#### Interventions that incorporated tailored computer software

- Czaja et al. (2018) conducted an RCT and assessed the impact of a specially designed computer system, the Personal Reminder Information and Social Management (PRISM) system for older adults at risk of social isolation on computer proficiency. PRISM included: Internet access (with vetted links to sites such as NIHSeniorHealth.Gov), an annotated resource guide, a dynamic classroom feature, a calendar, a photo feature, E-mail, games, and online help. Users can also be listed as a “PRISM Buddy” to enable them to have contact with people who had similar interests. Participants in the intervention group (PRISM) were provided with a Personal Computer (PC), monitor, mouse, keyboard, printer and access to the internet to complete the intervention at home. Participants in the comparator group were provided with a notebook with printed content similar to that within the PRISM system. **Participants in the intervention group were found to show a greater increase in CPQ scores at six months** (*b* −6.37; effect size 1.11; 95% CI: −7.39 to −5.35; p<0.001) and twelve months (*b* −7.06; effect size 1.23; 95% CI: −8.08 to −6.03; p<0.001) **compared to the comparator group.**

#### Interventions with an intergenerational component

- Garcia et al. (2022) analysed the impact of three different educational approaches (intergenerational, peer-to-peer and online) in improving older adults’ digital skills. The author’s assessed digital literacy by looking at the effectiveness on a range of outcomes including information and data literacy (I&DL), digital communication and collaboration (C&C) and safety, all of which statistically significantly improved compared to the baseline findings for all groups showing all delivery methods to be effective (p≤0.01). However, motivation was not reported to have improved for any of the groups. **Statistically significant differences in I&DL, C&C, and safety were identified in the peer-to-peer delivery method when compared to the online approach (p=0.001). Statistically significant differences in I&DL, C&C, and safety were identified in the intergenerational delivery method compared to the online approach (p=0.002).** No significant differences were reported between the peer-to-peer and intergenerational methods.

#### Interventions to improve online deception detection

- Moore and Hancock (2022) assessed the impact of a self-directed online course to improve deception detection online. Participants who received the intervention statistically significantly improved their ability to determine fake news after the intervention while participants in the control group also improved their ability after the intervention. However the improvement was not statistically significant (probability of accurately judging the veracity of a headline) 64% (95% CI: 61 to 67%) to 85% (95% CI: 82 to 88%) vs 55% (95% CI: 53 to 58%) to 57% (95% CI: 55 to 60%); respectively; p<0.001). **Participants in the intervention group were reported to have a statistically significantly greater improvement in deception detection compared to the control group** (B: 1.073, SE: 0.159, p < 0.001).

#### Traditional digital literacy interventions

- Elbaz et al. (2023) conducted a pilot study to assessed the impact of a four-week, online, digital literacy programme on computer proficiency, and found that the **mean Computer Proficiency Questionnaire (CPQ) scores were statistically significantly higher after the intervention compared to before the intervention** (CPQ score: 17.72 ±1.94 vs. 13.24 ±2.40; t(4)−8.910; p<0.001). More specifically, significant improvements were reported for computer basics (CPQ score 3.97 ±0.45 vs. 3.23 ±0.60; t(4)−5.880; p=0.004), communication (CPQ score 3.36 ±0.38 vs. 2.40 ±0.48; t(4)−8.353; p=0.001), and using the Internet (CPQ score 3.63 ±0.16 vs. 2.71 ±0.48; t(4)−4.257; p=0.013). No significant differences were reported for printing, scheduling online, and using multimedia (p>0.05).
- Seaton et al. (2023) explored the impact of the Gluu Essentials digital skills training programme on mobile device proficiency and confidence in going online in older adults living in rural communities. This included 12 lessons delivered either in person or over the phone. The study findings reported a statistically significant improvement in **mobile device proficiency after the intervention** (mean mobile device proficiency questionnaire (MDPQ) score 3.93, SD:0.91 vs 4.13, SD:0.79; p<0.001).
- Quialheiro et al. (2023) measured the impact of an eight-workshop digital inclusion programme - the OITO (Oficinas de Inclusão Tecnológica Online, “Workshops for Online Technological Inclusion”) project on digital literacy of older adults at three time points – before, immediately after, and at one month after completing the programme. **The results showed statistically significant improvements in MDPQ score after the intervention and at one month follow-up compared to before the intervention** (mean MDPQ increase 2.49; 95% CI: 1.80 to 3.18; p<0.001), but without a statistically significant difference between the post-intervention times.
- McCosker et al. (2020) assessed the impact of an online, community-based national digital inclusion programme (Be Connected) on digital skills, which consisted of 12 modules. **Participants improved in all 11 operational and strategic digital skills measured after the intervention, but this improvement was only statistically significant for 10 of the skills reported.** This included statistically significant changes to their ability to: make basic changes to others content (p=0.007), bookmark a website (p<0.001), create something new from existing online images music or video (p=0.001), install apps (p<0.001), understand which licenses apply to online content (p=0.005), to open a tab in a browser (p=0.031), to open downloaded files (p=0.048), to download/save a photo (p=0.036), use shortcut keys (p<0.001), keep track of mobile app costs (p<0.001). No significant change was found in participants abilities to create a website (p=0.016).
- Patty et al. (2018) explored the impact of a community-based Information and communication technology (ICT) training intervention (provided as part of standard rehabilitative eye care) on a range of ICT skills for visually impaired older adults. Using an adapted version of the Dutch Activity Inventory (D-AI) to evaluate the effects of training, authors found **ICT skills statistically significantly improved (perceptions of difficulty decreased) after the intervention and continued to improve three months after the intervention** (D-AI sum score: 22.98 (at baseline) vs 13.13 (after the intervention) vs 12.97 (three-month follow-up); p=0.01). The most notable improvements were in computer skills, using the internet and using hotkeys. This study also reported cost-effectiveness outcomes (summarised in section 2.6).
- Holguin-Alvarez et al. (2020) assessed the impact of a 50-session, community-based, social media programme to increase digital competence for older adults of lower SES. The programme supported participants to engage with social media sites and to communicate online. **The experimental group statistically significantly improved their digital competence compared to the control group** (average digital competency score M:119.1; SD:0.24 vs M:45.1; SD:1.06; p<0.001).
- Martínez-Alcalá et al. (2018) assessed the impact of digital literacy workshops for older adults on digital competence. Two different delivery methods were assessed; a face-to-face approach and a blended approach (which included multimedia learning activities and materials as well as face-to-face). **The workshops statistically significantly improved digital competence in both groups** (face-to-face approach: *z*:−6.79, p<0.0001 and blended approach: *z*:−5.30, p<0.0001). The findings show that **participants in the blended workshop group reported a statistically significant greater improvement in digital competence compared to the face-to-face group** (difference in competence *U*:810.5, p<0.01).

#### Effectiveness of educational interventions on E-health literacy

Two studies (Lee and Kim., 2019; Lee et al., 2022b) assessed the effectiveness of interventions for improving E-health literacy in older adults. Interventions in both studies incorporated both educational and mentorship approaches. A pre-post study design was used to measure E-health literacy in Lee and Kim 2019, while a non-randomised controlled study design was used in Lee et al 2022b. Study findings appeared to show effects in favour of the interventions.

#### Interventions with an intergenerational component

- Lee and Kim (2019) explored the impact of a community-based group, intergenerational programme on E-health literacy. The programme included six group sessions in the community where older adults were paired with student mentors. E-Health literacy was measured before the intervention and found to be neutral (defined as undecided), however, **after the intervention participants showed a statistically significant improvement** (t:−5.89, d:−0.79; p<0.001).
- Lee et al. (2022b) assessed the impact of a 12-week, community-based, intergenerational, group, educational programme where older adults were paired with student mentors and found that E-health literacy **statistically significantly improved for individuals who received the intervention compared to the control group** (who received no intervention) (t(49):−4.23, d:−0.60; p<0.001).

#### 2.3.1 Bottom line results for the effectiveness of educational interventions on digital literacy

There is evidence to suggest that educational interventions are effective at improving digital literacy in older adults. The results suggest that interventions that incorporate gamification, tailored computer software, intergenerational approaches, or aim to teach specific digital literacy skills such as deception detection are effective at improving digital literacy as were the traditional digital literacy interventions.

Evidence of improvements were also seen for older adults in vulnerable contexts including those at risk of social isolation, lower SES groups, those living in rural areas and visually impaired older adults. However, the evidence in support of these interventions is of low quality, so we cannot be certain of the true effect.

Improvements were reported regardless of whether the intervention was delivered in the community, at home or online. While all delivery methods identified within the included studies improved participants digital literacy, there is limited evidence to suggest peer-to-peer or intergenerational training may be more effective than online training, and that blended training involving multimedia activities and materials may be more effective than face-to-face training.

### 2.4 Effectiveness of educational interventions on participants’ perceptions of technology use

A total of nine studies reported participant perceptions of technology use (Arthanat., 2021; Castilla et al., 2018; Czaja et al., 2018; Fields et al., 2021; Lee and Kim., 2019; Lee et al., 2022a; Lee et al., 2022b; Ma et al., 2020; Quialheiro et al., 2023). This outcome included self-perceptions of abilities when using digital technologies (Castilla et al., 2018; Czaja et al., 2018; Fields et al., 2021; Lee and Kim., 2019; Lee et al., 2022a; Ma et al., 2020; Quialheiro et al., 2023), technophobia (Lee and Kim., 2019; Lee et al., 2022b) and acceptability of technology (Arthanat., 2021; Castilla et al., 2018; Czaja et al., 2018; Lee and Kim., 2019). Study findings appeared to show effects in favour of the interventions.

#### Effectiveness of educational interventions on participants’ self-perceptions of abilities

Seven studies explored the impact of interventions on participants’ perceptions of their own abilities to use new technologies, this included perceptions of capabilities, self-efficacy, self-autonomy and confidence as a standalone outcome (Castilla et al., 2018; Czaja et al., 2018; Fields et al., 2021; Lee and Kim., 2019; Lee et al., 2022a; Ma et al., 2020; Quialheiro et al., 2023). This outcome was measured using a pre-post study design in three studies (Castilla et al., 2018; Lee and Kim., 2019; Quialheiro et al., 2023), a non-randomised controlled study design in two studies (Lee et al., 2022a; Ma et al., 2020) and a RCT study design in two studies (Czaja et al., 2018; Fields et al., 2021).

#### Interventions that incorporated tailored computer software

- Castilla et al. (2018) assessed the effectiveness of an eight-session, community-based, group, social network (Butler 2.0) training programme on attitudes to using new technologies in older adults living in rural areas. **Statistically significant improvements were reported in participant perceptions of their capability to use new technologies after receiving the intervention** (score from 1-4; mean: 2.3; SD:0.92; to mean 2.74; SD:0.80; p=0.012).
- Czaja et al. (2018) assessed the impact of the PRISM system (home-based) on attitudes towards computer self-efficacy for older adults at risk of social isolation. Participants in the intervention group reported a greater **increase in computer self-efficacy at 6 months** (*b*:−1.29; effect size: 0.41; 95% CI: −2.01 to −0.57; p<0.001) **and 12 months** (*b*:−0.94; effect size: 0.30; 95% CI: −1.67 to −0.22; p<0.02) follow-up **compared to the comparative group** and these increases were statistically significant.

#### Interventions with an intergenerational component

- Lee and Kim (2019) assessed the impact of a community-based, group, intergenerational programme on a range of attitudes including perceived self-efficacy and found that participants perceptions of **self-efficacy statistically significantly improved after receiving the intervention** (t:−8.36, d:−1.13; p<0.001).
- Ma et al. (2020) explored the impact of a three-session video tutorial-based intervention in the community on perceived digital self-efficacy. The intervention was delivered to three separate intervention groups that differed according to the model used in the videos (a child model, young adult model, or older adult model). The results found that digital **self-efficacy increased significantly (p<0.001) after training regardless of the delivery method. However, the older adult model group contributed to the highest improvement in self-efficacy** (F:3.878; p<0.05). **No significant difference was found between the child model group and the young adult model group.**

#### Interventions incorporated into existing services

- Fields et al. (2021) assessed the impact of an at home technology training intervention on participants self-reported confidence for socially isolated older adults. The intervention was incorporated into a volunteer-based programme that provides home visits for lonely older adults. Although not statistically significant, **the intervention group reported higher levels of self-confidence in their digital skills** (baseline: 52% little to no confidence searching for information online and using email vs two-months: 35% little to no confidence, p=0.13). No change was reported for the control group (baseline: 76% little to no confidence vs 77% little to no confidence at two months, p=1.0). **Those in the intervention group reported higher levels of confidence** (OR: 8.99; 95% CI: 1.55 to 96.57, at two months follow-up) **compared with those in the waitlist group.**

#### Traditional digital literacy interventions

- Quialheiro et al. (2023) assessed the impact of an in-person digital inclusion programme (OITO project) on perceptions **of self-reported autonomy** when it comes to using a mobile phone or tablet with a touchscreen and found a **statistically significant improvement one month after the intervention**, increasing from 4.5 at baseline to 6.7 points, with a possible score ranging from 0 to 10 (t(40):–7.3; p<0.001).
- Lee et al. (2022a) assessed the impact of a community-based, digital literacy education programme on perceived self-efficacy and found that those in the **intervention group reported improved self-efficacy after the intervention, however this was not statistically significant** (57.1-58.1 p=0.530), **whereas the self-efficacy of the control group was reported to significantly increase after the intervention** (55.6-60.1 p=0.025). **However, when compared, no significant difference was reported between groups** (t:-1.382; 95% CI: −8.499 to 1.494; p=0.169).

#### Effectiveness of educational interventions on technophobia

Two studies assessed the impact of interventions incorporating both education and mentorship on technophobia (using measurements of confidence and anxiety) (Lee and Kim., 2019; Lee et al., 2022b). This outcome was measured using a pre-post study design in one study (Lee and Kim 2019) and a non-randomised controlled study design in one study (Lee et al 2022b).

#### Interventions with an intergenerational component

- Lee and Kim (2019) found that participants who took part in the community-based, group intergenerational programme reported **a statistically significant increase in their perceived confidence about their skills when using computers/Internet** (t:−3.69, d:−0.50; p<0.001) **and that anxiety toward technology statistically significantly decreased after receiving the intervention** (t:2.65, d:0.36; p<0.01).
- Lee et al. (2022b) found that participants who took part in a 12-week, community-based, group, intergenerational educational programme **reported a statistically significant increase in confidence in using technology** (mean confidence score 2.59 vs 2.87; t(49):−5.05, d:−.71; p<0.001) **and a statistically significant decrease in anxiety** (mean anxiety score 2.88 vs 3.07; t(49):−2.77, d:−.39; p<0.01) **after receiving the intervention.** However in the control group, anxiety was also significantly reduced (mean anxiety score 2.82 vs 3.00; t(53):−2.04, d:0.28; p<0.05), and an insignificant increase in confidence was reported (mean confidence score 2.50 vs 2.58; t(53):-1.48, p=0.15). **Confidence was the only outcome reported to have differed among the two groups with a statistically significant increase being reported for the intervention group compared to the control** (mean confidence score M:2.87, SD:0.47, M_adj_:2.84 vs M:2.58, SD:0.56, M_adj_:2.61; F(1,101):9.99; η2p:0.09; p=0.002).

#### Effectiveness of educational interventions on acceptability of technology

Four studies explored the impact of interventions on participants’ acceptability of technology (Arthanat., 2021; Castilla et al., 2018; Czaja et al., 2018; Lee and Kim., 2019). This outcome was measured using a pre-post study design in two studies (Castilla et al., 2018; Lee and Kim., 2019) and a RCT study design in two studies (Arthanat., 2021; Czaja et al., 2018).

#### Interventions that incorporated tailored computer software

- Castilla et al. (2018) assessed the effectiveness of an eight-session, community-based, group, social network (Butler 2.0) training programme on attitudes towards new technologies for older adults living in rural areas. **Participants’ attitudes toward new technologies were found to improve after receiving the intervention.** Statistically significant improvements were reported in participants level of interest in using new technologies (p=0.003). Improvements were also reported in how participants felt when using new technologies however this difference was not statistically significant (p>0.500).
- Czaja et al. (2018) found that **compared to the comparative group, participants at risk of social isolation who received the home-based PRISM intervention reported statistically significant increases in computer comfort at six months** (*b*:−1.68; effect size: 0.39; 95% CI: −2.57 to −0.78; p<0.001) **and 12 months** (*b*:−2.32; effect size: 0.53; 95% CI: −3.22 to −1.41; p<0.001); **and in computer interest at six months** (b:−1.52; effect size: 0.46; 95% CI: −2.26 to −0.79; p<0.001) **and 12 months** (b:−0.99; effect size: 0.30; 95% CI: −1.74 to −0.25; p<0.01).

#### Interventions with an intergenerational component

- Lee and Kim (2019) found that participants who participated in the community-based, group, intergenerational programme **showed a statistically significant improvement in their interest towards computers after the intervention** (t:−9.24, d:−.25; p<0.001).

#### Interventions incorporated into existing services

- Arthanat (2021) found that participants living in rural areas who received the ICT training programme (i-CHATT) at home as part of an occupational therapy programme**, reported more positive responses about technology when compared to the control group, across the six, 12, 18, and 24-month follow-up.** Statistically significant differences in favour of the intervention were reported for perceiving technology experiences as satisfying (F(4,1):3.5, ηp2: 0.04; p=0.007), seeing technologies as encouraging (F(4,1):2.4, ηp2: 0.01; p=0.05), comfort with technology (F(4,1):3.6, ηp2: 0.04; p=0.009), and feeling good around technology (F(4,1):2.3, ηp2: 0.02; p=0.05).

#### 2.4.1 Bottom line results for effectiveness of educational interventions on participant perceptions of technology use

There is evidence to suggest that educational interventions can improve a range of perceptions among older adults including their own perceived abilities, increased acceptability of technology, and reduced technophobia. However, the evidence in support of these interventions is mostly of low quality. Perceived abilities improved in all studies reporting outcomes included in this category, irrespective of the setting or mode of delivery of the interventions. Various methods were used to assess participant perceptions making it difficult to compare studies directly.

A range of intervention approaches were found to be effective in improving participants’ perceptions of own abilities including those using tailored computer software, those with intergenerational components, those incorporated into existing service and traditional digital literacy interventions. Technophobia was found to decrease in the two studies reporting this outcome, both of which utilised an intergenerational approach. Acceptability of technology was also seen to improve regardless of the intervention method, including interventions that incorporated tailored computer software, that had intergenerational components or interventions that were incorporated into existing services.

When delivery methods were compared among an older population, one study suggested that using an older adult model in video tutorials improved participants’ self-efficacy more than when using the child or young adult model, suggesting a peer-to-peer approach was preferred over an intergenerational approach. However, as only one study reported this finding firm conclusions cannot be made.

While all studies had methodological limitations, there is evidence to suggest that educational interventions can improve perceptions of technology and perceived ability to engage with the digital world with two studies showing improvements for older adults living in rural areas.

### 2.5 Acceptability of educational interventions

A total of nine studies reported participants’ acceptability of the interventions (Castilla et al., 2018; Choi and Park., 2022; Czaja et al., 2018; Elbaz et al., 2023; Fields et al., 2021; Gadbois et al., 2022; Lee and Kim., 2019; Martínez-Alcalá et al., 2018; Seaton et al., 2023). This outcome was measured using a pre-post study design in five studies (Castilla et al., 2018; Elbaz et al., 2023; Gadbois et al., 2022; Lee and Kim., 2019; Seaton et al., 2023), a controlled study design in two studies (Choi and Park., 2022; Martínez-Alcalá et al., 2018), and a RCT study design in two studies (Fields et al., 2021; Czaja et al., 2018). Study findings appeared to show effects in favour of the interventions. Acceptability of the intervention was obtained through focus groups or questionnaires.

#### Interventions that incorporated tailored computer software

- Castilla et al. (2018) found that 96% of older adults living in rural areas who participated in the eight-session, community-based, group, social network (Butler 2.0) training programme **expressed the intention to continue to use the Butler 2.0 social network** in future, compared to 2% who expressed no intention to use it, and 2% who were unsure.
- Czaja et al. (2018) found that most participants at risk for social isolation who received the PRISM system at home, **found the software useful in their daily life (82%),** indicated that it made their life easier (80%), improved their daily life (84%), and enabled them to accomplish tasks more quickly (73%). They also found PRISM easy to use (88%) and easy to become skilled at using PRISM (80%). Participants found it easier to communicate with family and friends compared to controls (82% vs. 47%) and engage in hobbies and play games (82% vs. 52%). Participants in both conditions reported that it was easier to look up community information (78% vs. 73%) and health information (82% vs. 80%). **However, those in the comparative group were more satisfied with the in-home training they received (90% vs. 82%).**

#### Interventions incorporated into existing services

- Fields et al. (2021) found that participants who received the at home technology training (incorporated into a volunteer-based programme providing home visits for lonely older adults), **would have liked to receive more sessions, specifically with the volunteer instructors and that some participants appreciated the personalised approach**.
- Gadbois et al. (2022) found that almost all of the homebound participants who received the pilot talking tech intervention incorporated within a home-delivered meals programme **described having a positive experience.** A few participants suggested including more training sessions to accommodate participants with a slower learning pace or for those starting with limited technology knowledge. Alternatively, others with more computer experience felt aspects of the module content were too basic.

#### Interventions with an intergenerational component

- Lee and Kim (2019) found that participants who participated in the community-based, group, intergenerational programme **reported multiple benefits after receiving the intervention.** Benefits emerged into four major themes: communication tools, independent living, leisure activities, and intergenerational learning.

#### Gamification

- Choi and Park (2022) found that **participants who received the community-based, educational IT programme with the educational game ‘Save the Titanic using the decision tree’, reported high levels of satisfaction** (mean satisfaction score (on a 5-point likert scale) 4.13 (±0.65). More specifically, satisfaction with the educational content, educational activity and educational material was all reported to be high (mean satisfaction score; 4.22 (±0.64); 3.94 (±0.76); 4.05 (±0.69); respecitvely).

#### Traditional digital literacy interventions

- Elbaz et al. (2023) found that some participants who received the pilot four-week, online, digital literacy programme described being confused as to why certain applications appeared different (i.e., had a different name compared to what the trainer was showing, different icons, or the user interface was not the same). **However, overall, all participants appreciated the digital literacy training as well as the accessibility, and patience of the trainer and facilitators.**
- Seaton et al. (2023) found that participants’ (living in rural areas) **acceptability of the Gluu Essentials digital skills training programme was high after the intervention**. Participants’ recommendations included the need for providing ongoing programmes for support and training because technology constantly changes, reducing costs for technology and internet access, and keeping learning resources simple and easy to access.
- Martínez-Alcalá et al. (2018) found that of those who received the blended workshops (37 participants), **13 participants indicated a positive agreement stating that the interaction with the system was clear and understandable and even the menu was easy to use.** A total of 16 participants stated that it was useful to implement this type of workshop to improve the populations digital literacy skills, 15 older adults were enthusiastic about using the platform, 15 older adults indicated a positive agreement stating that they will use the system to reinforce their knowledge during and after the workshop.

#### 2.5.1 Bottom line results for the acceptability of digital education interventions

There is considerable evidence to show that participants had positive perceptions of the educational interventions. This included interventions that incorporated tailored computer software, interventions that were incorporated into existing services, intergenerational interventions, interventions that incorporate gamification and traditional digital literacy interventions.

The evidence in support of this is generally of low quality. Positive perceptions of the interventions were also found for some sub-populations including participants living in rural areas, being at risk of social isolation or being homebound, and the findings do not appear to be dependent on the delivery method used (at home, online, in groups).

### 2.6 Cost-effectiveness of educational interventions

One study assessed the cost-effectiveness of an educational intervention (Patty et al., 2018). This outcome was measured using a pre-post study design (at 3-months follow-up). The study findings appeared to show effects in favour of the intervention.

#### Traditional digital literacy interventions

- Patty et al. (2018) assessed the cost-effectiveness of a community-based ICT training intervention for visually impaired older adults in the Netherlands and found that the intervention was cost-effective but under the assumption that the effects of the training remain consistent for 10 years. This would result in an incremental +-effectiveness ratio (ICER) of € 11,000 per quality-adjusted life-year (QALY) and € 8000 per year of well-being gained, when only the costs of ICT training are considered. Furthermore, when the willingness-to-pay threshold is € 20,000 per year of well-being, the probability that ICT training will be cost-effective is 91% when including only the costs of ICT training).

#### 2.5.2 Bottom line results for cost-effectiveness of educational interventions

There is very limited, low-quality evidence to suggest that educational interventions for visually impaired older adults are cost-effective. However, as this outcome was reported by only one study firm conclusions cannot be made.

**Table 2:**
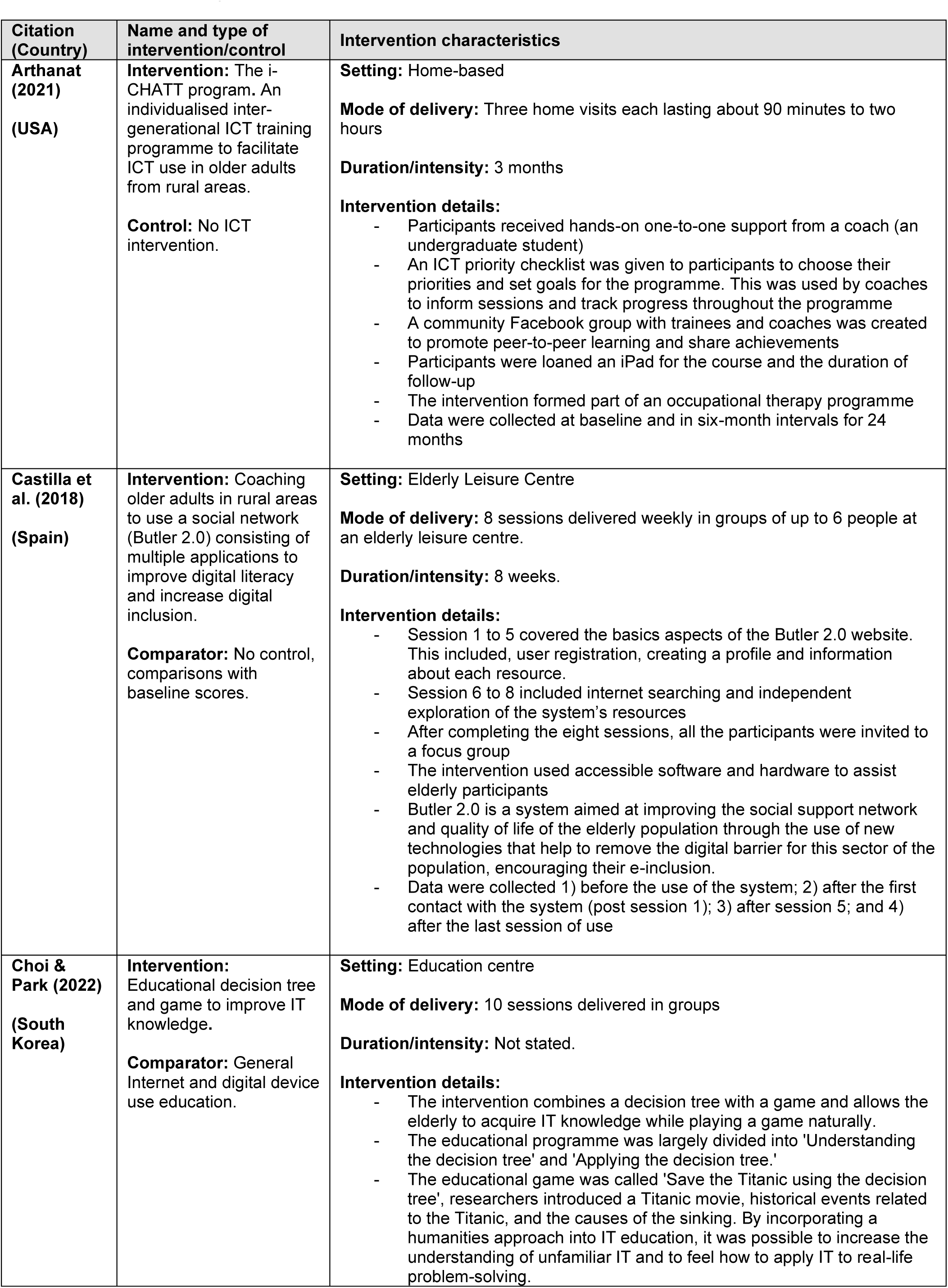
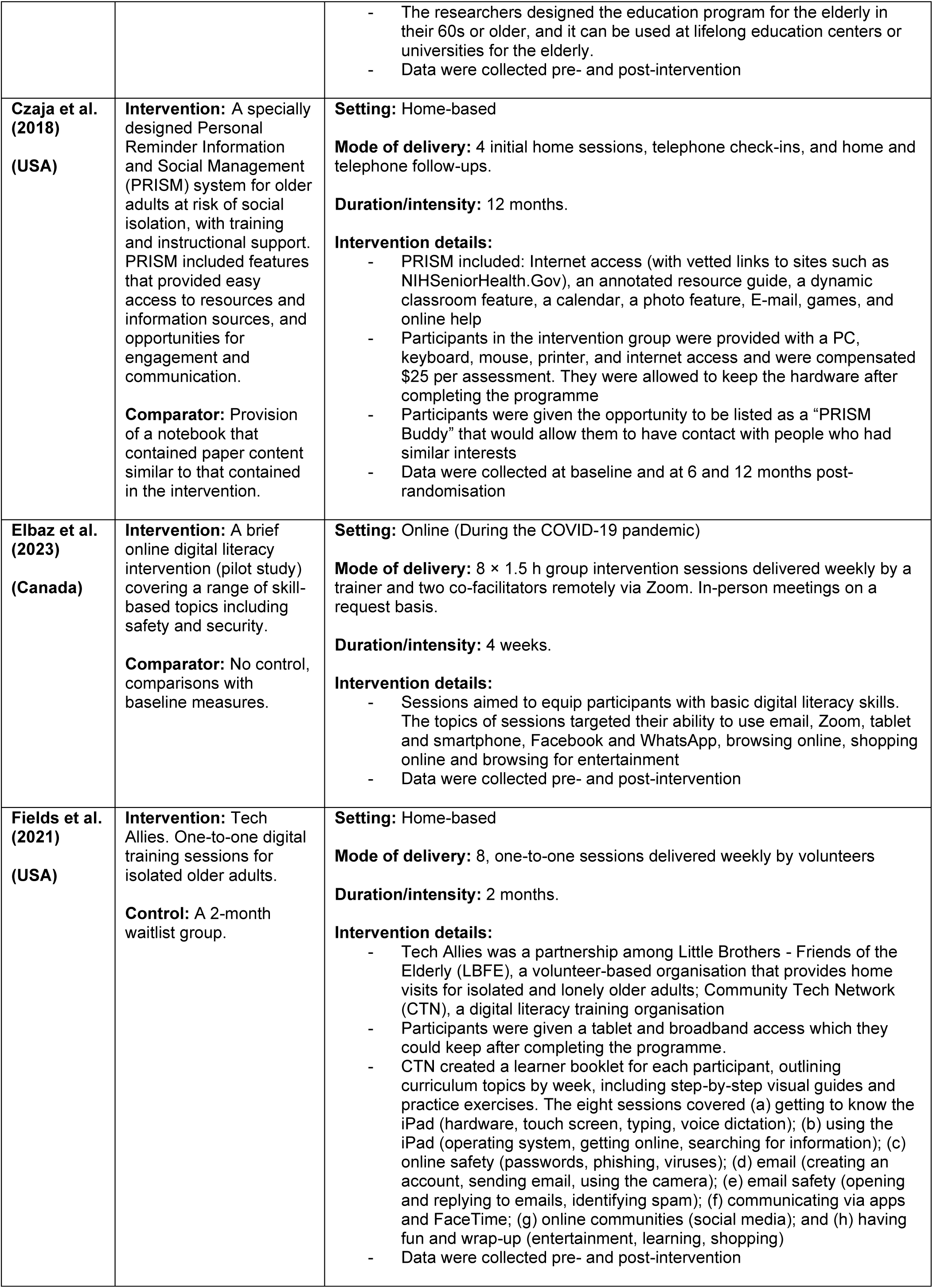
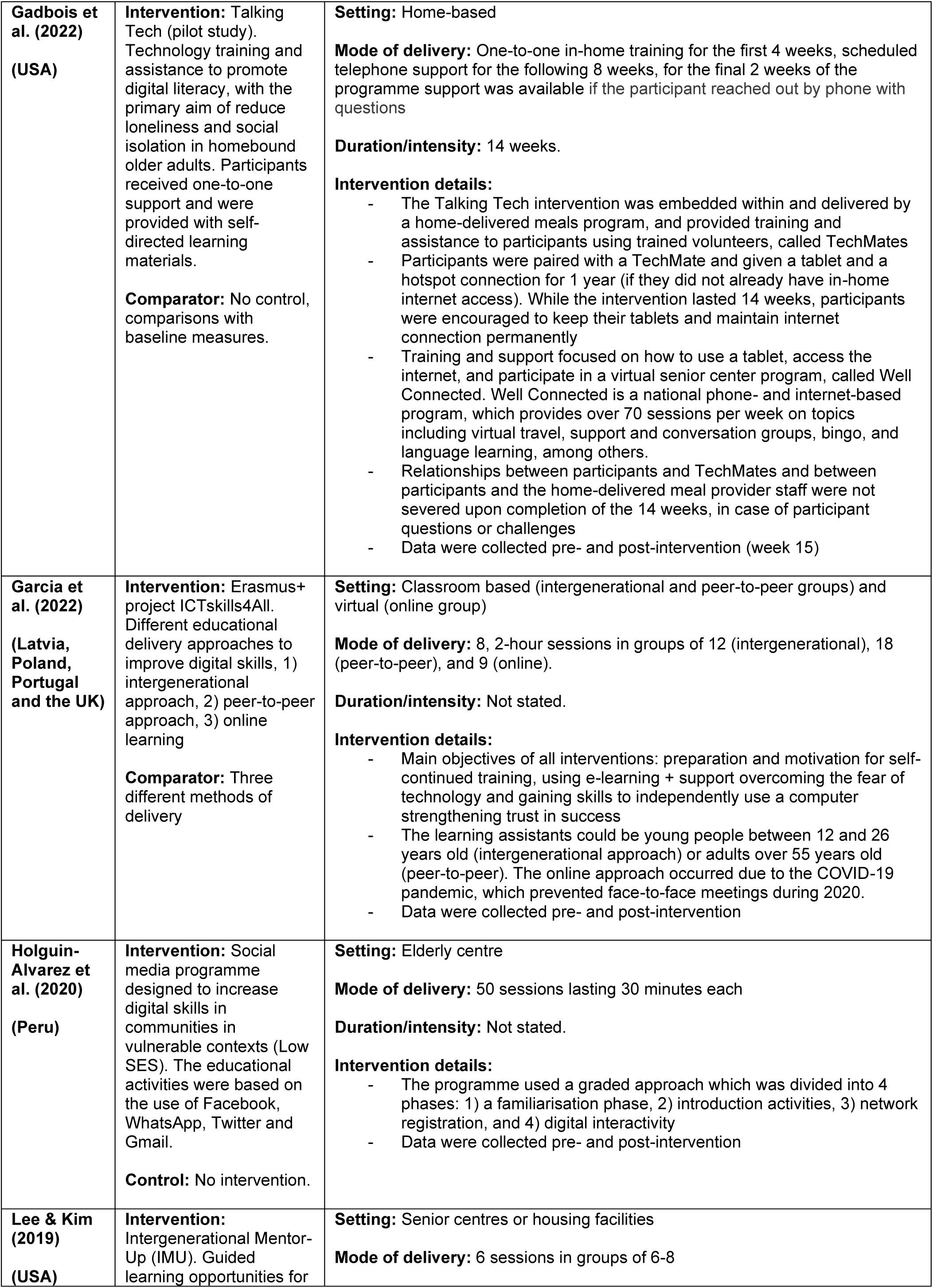
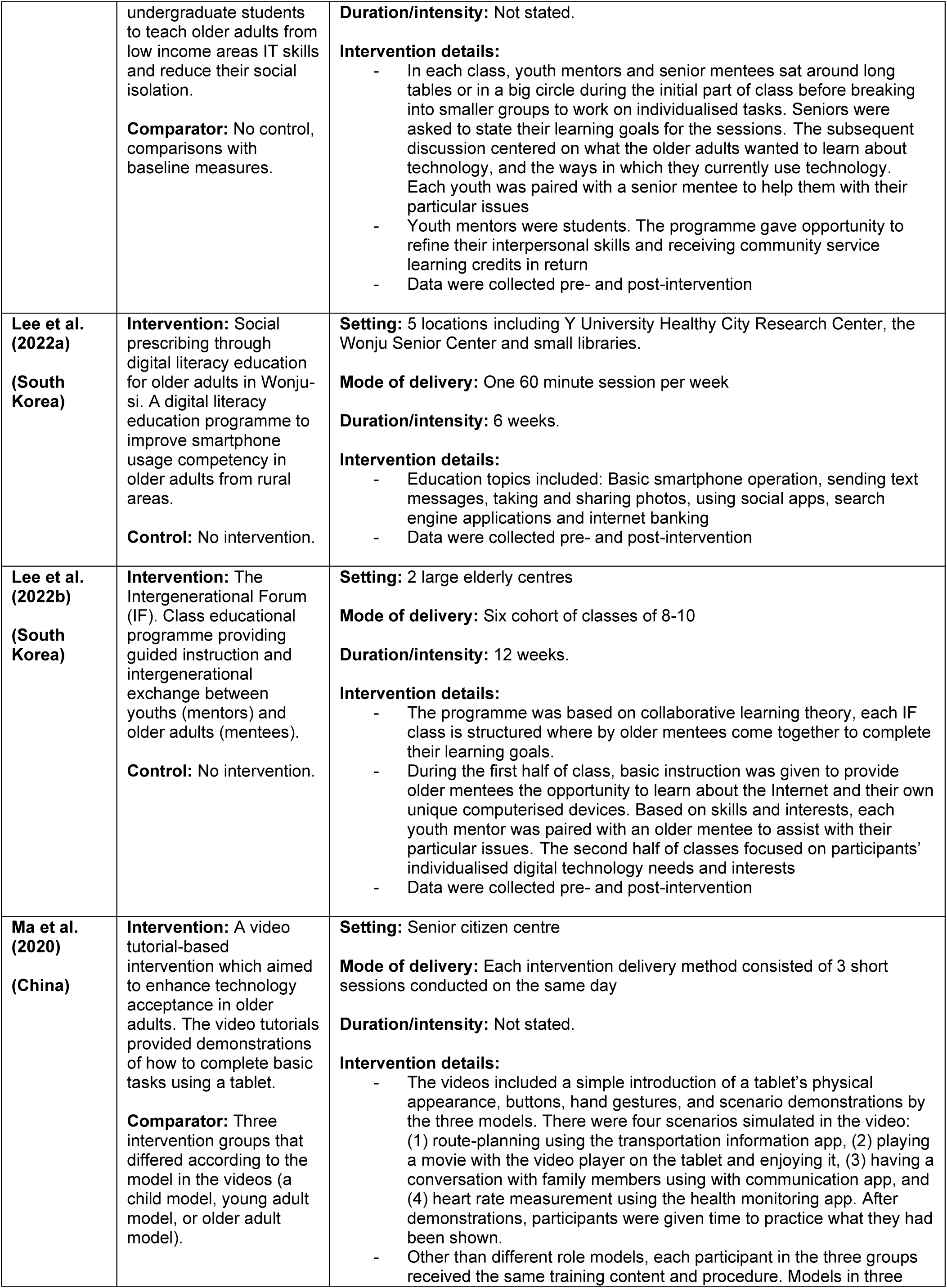
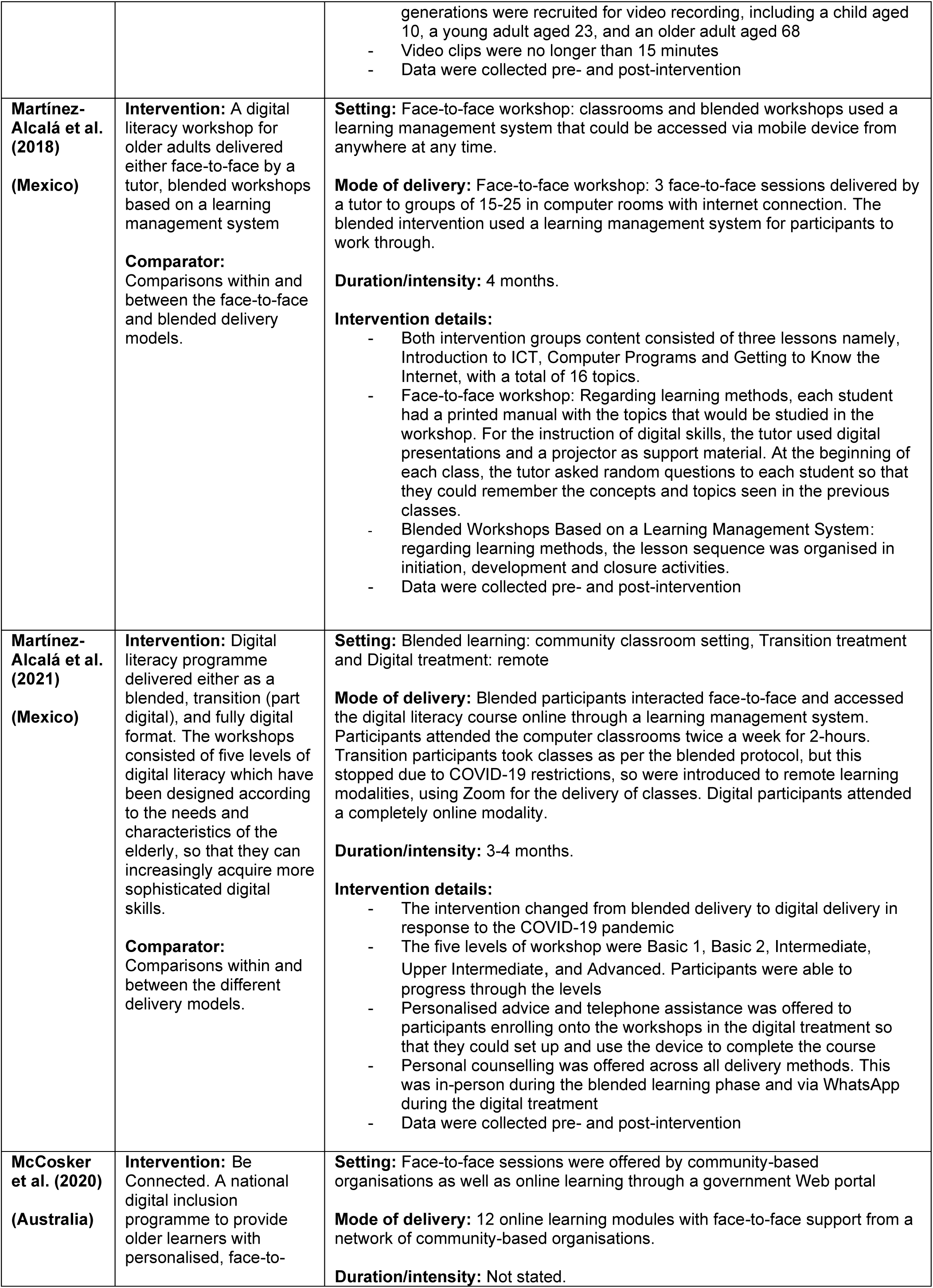
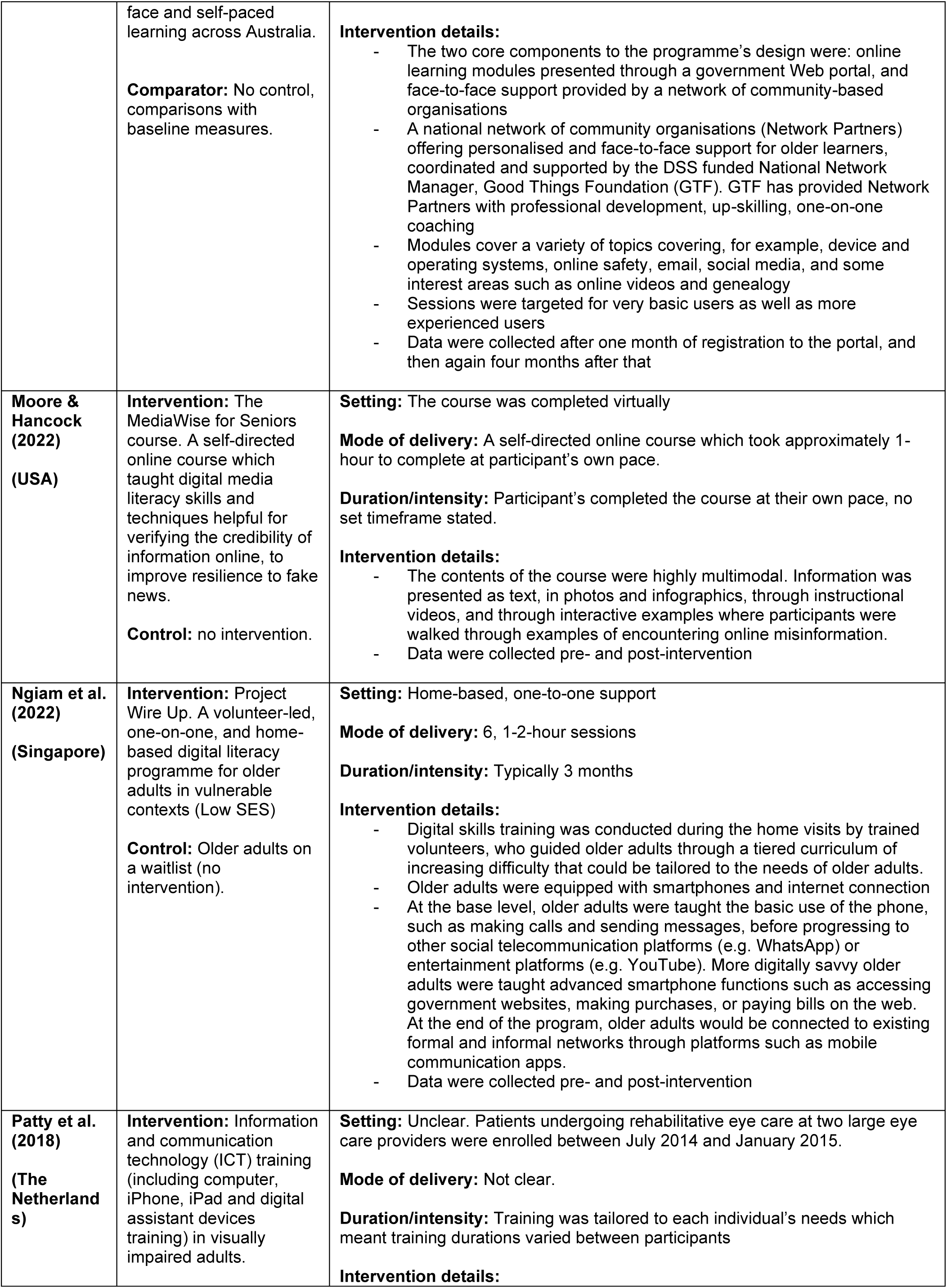
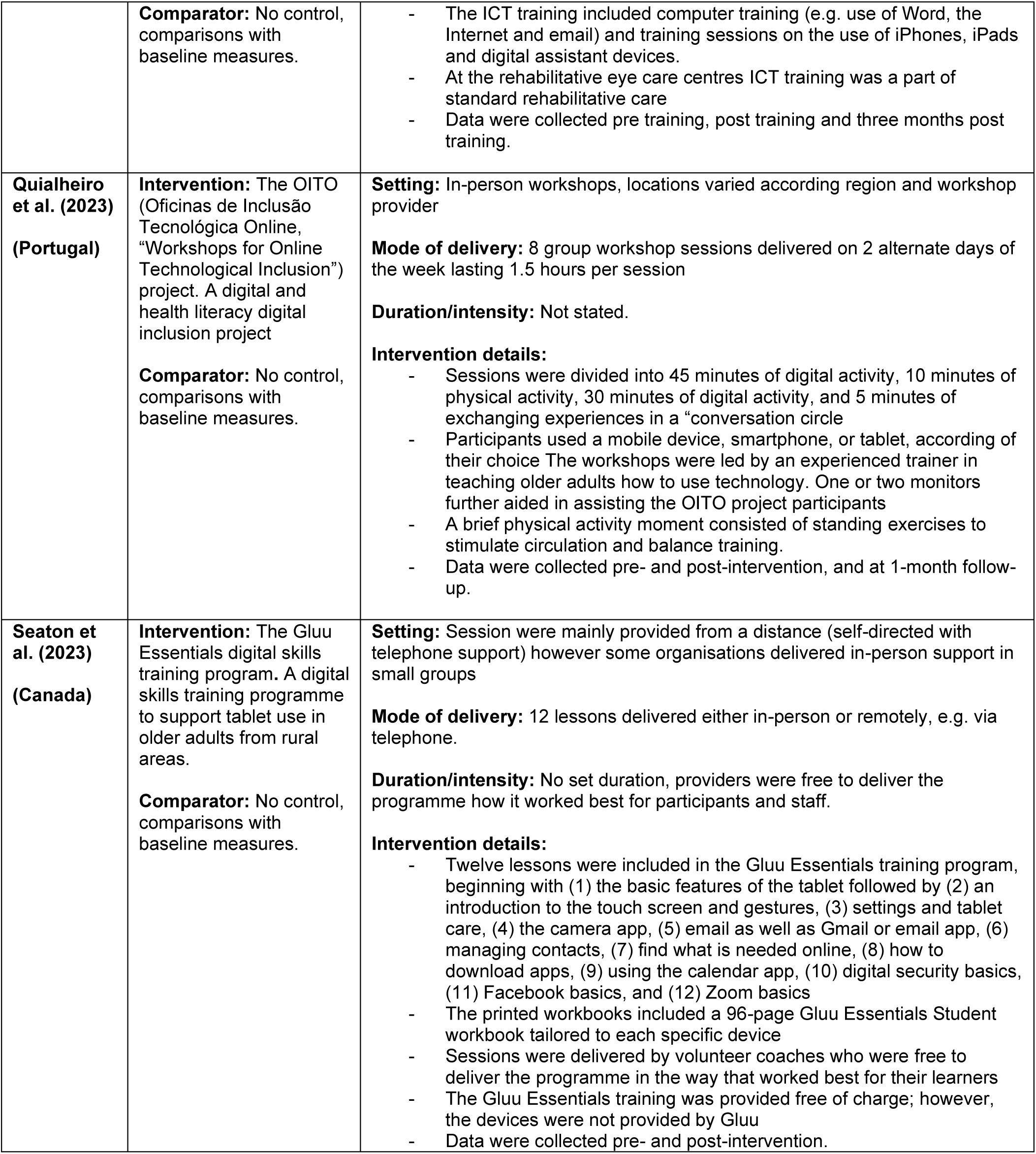
Summary of interventions.

**Table 3.** Summary of findings.

| Intervention Outcome<br>[No. of studies] | Direction of effect<sup>1</sup> | Study design | Limitations | Inconsistency | Certainty |
| --- | --- | --- | --- | --- | --- |
|  |  |  | Risk of bias from critical appraisal checklist | Variation in findings across studies (eg CIs don't overlap) |  |
| **Technology use/adoption** |  |  |  |  |  |
| **Use/Uptake/Adoption of technology**<br>[5] | **Favours intervention** (Arthanat., 2021; Fields et al., 2021; Gadbois et al., 2022; Lee et al., 2022a; Seaton et al., 2023) | **3 Quasi-experimental**<br>**2 RCT** | **3 Low Quality**<br>**2 Moderate Quality** | Results appear to be positive however two studies reported no effect for some individual outcomes, however this could be a results of differing personal needs. Outcome measures also varied across studies. | Low certainty |
| **Digital literacy/skills** |  |  |  |  |  |
| **Digital literacy**<br>[3] | **Favours intervention** (Choi and Park., 2022; Martínez-Alcalá et al., 2021; Ngiam et al., 2022) | **3 Quasi-experimental** | **3 Low quality** | Results appear to be consistently positive however outcome measures varied | Low certainty |
| **Proxies for digital literacy**<br>(includes computer/mobile device proficiency, digital competence, digital/ICT skills and online deception detection)<br>[10] | **Favours intervention** (Czaja et al., 2018; Elbaz et al., 2023; Garcia et al., 2022; Holguin-Alvarez et al., 2020; Martínez-Alcalá et al., 2018; McCosker et al., 2020; Moore and Hancock., 2022; Patty et al., 2018; Quialheiro et al., 2023; Seaton et al., 2023) | **9 Quasi-experimental**<br>**1 RCT** | **9 Low quality**<br>**1 Moderate quality** | Results appear to be consistently positive but may depend on specific competences measured. | Low certainty |
| **E-health literacy**<br>[2] | **Favours intervention** (Lee & Kim., 2019; Lee et al., 2022b). | **2 Quasi-experimental** | **2 Low Quality** | Results appear to be positive however the outcome was only reported by two studies. | Very low certainty |
| **Participant perceptions of technology use** |  |  |  |  |  |
| **Self-perceptions of abilities**<br>[7] | **Favours intervention** (Castilla et al., 2018; Czaja et al., 2018; Fields et al., 2021; Lee & Kim., 2019; Ma et al., 2020; Quialheiro et al., 2023)<br>**Neutral** (Lee et al., 2022a) | **5 Quasi-experimental**<br>**2 RCT** | **5 Low Quality**<br>**2 Moderate Quality** | Results appear to be mostly positive however one study reported no significant improvement in self-efficacy following the intervention. | Low certainty |
| **Technophobia**<br>[2] | **Favours intervention** (Lee & Kim., 2019; Lee et al., 2022b) | **2 Quasi-experimental** | **2 Low Quality** | Results appear to be positive but limited by number of studies. | Very low certainty |
| **Acceptability of technology**<br>[4] | **Favours intervention** (Arthanat., 2021; Castilla et al., 2018; Czaja et al., 2018; Lee & Kim., 2019). | **2 Quasi-experimental**<br>**2 RCT** | **2 Low Quality**<br>**2 Moderate Quality** | Results appear to be consistently positive. | Low certainty |
| **Acceptability of interventions** |  |  |  |  |  |
| **Acceptability of intervention(s)**<br>[9] | **Favours intervention** (Choi & Park., 2022; Castilla et al., 2018; Czaja et al., 2018; Elbaz et al., 2023; Fields et al., 2021; Gadbois et al., 2022; Lee & Kim., 2019; Martínez-Alcalá et al., (2018); Seaton et al., 2023) | **7 Quasi-experimental**<br>**2 RCT** | **7 Low Quality**<br>**2 Moderate Quality** | Results appear to be consistently positive even when constructive criticism was provided. | Low certainty |
| **Cost-effectiveness** |  |  |  |  |  |
| **Cost effectiveness**<br>[1] | **Favours intervention** (Patty et al., 2018) | **1 Quasi-experimental** | **1 Low Quality** | Results appear to be positive however only one study reported this outcome. | Very low certainty |

1. Positive findings indicate where the results are in favour of the intervention(s), however this is not dependent of statistical significance.

## 3. DISCUSSION

### 3.1 Summary of the findings

The aim of this rapid review was to identify primary research addressing digital exclusion in older adults. While none of the included studies specifically aimed to address digital exclusion, 21 primary studies were identified that indicate educational interventions have the potential to improve digital literacy, digital use, perceptions of technology and participant acceptability among older adults. This included interventions that were incorporated into existing services, intergenerational interventions, interventions that incorporated tailored computer software or gamification, those teaching specific skills such as deception detection and more traditional digital literacy interventions.

The review findings indicate that educational interventions have the potential to improve digital literacy among this population. While most of the evidence demonstrated positive results in the general older adult population, improvements were also observed in various sub-populations, particularly older adults in vulnerable contexts such as individuals living in rural areas, those at risk of social isolation, homebound, visually impaired, or belonging to lower SES groups. Improvements in digital literacy were reported in both community and home-based settings. Findings also suggest ICT training interventions may be cost-effective, however this was only from one study, which was conducted in a population of visually impaired older adults.

The review findings indicate that educational interventions may improve digital use among older adults. Positive outcomes were reported in studies involving rural and homebound population groups, as well as the general older adult population. Furthermore, this rapid review shows that educational interventions may enhance positive perceptions of technology and the perceived ability to engage with the digital world. This is an important aspect in addressing established barriers to digital inclusion, such as the fear and anxiety associated with the use of technologies highlighted previously. Improvements in perceived abilities were consistently reported across all studies that reported this outcome, irrespective of the intervention delivery format or location.

Regarding participant acceptability, positive perceptions were reported by participants irrespective of country, population group or intervention delivery method/approach. Despite some participant feedback suggesting areas to improve interventions, participants largely reported the education and training provided were valued and beneficial.

A small number of the included studies evaluated an intervention that included providing participants with digital devices and internet access. Providing devices and access to the internet may increase online engagement and address the affordability and accessibility issues related to digital exclusion. Qualitative feedback also highlighted the need for continued support with technology and internet access after the intervention, to facilitate sustained use of digital devices and services.

### 3.2 Strengths and limitations of the available evidence

The available evidence included a range of vulnerable population groups. This included rural, visually impaired, lower SES and homebound older adults. However, the majority of studies were focused on older adults in general. Twenty-one studies published within the previous 5-years were identified, indicating the recency of internationally published evidence available to inform future policy and practice.

An additional strength of the evidence is the use of a range of intervention delivery methods, such as online, in-person, community- and home-based approaches. Furthermore, several different aspects of digital inclusion, such as proficiency, competence, technology use and uptake, and several others, were investigated across the included studies, providing a range of outcomes. This enables a more comprehensive and wide-ranging assessment of intervention effectiveness.

Several evidence gaps were identified in the evidence base, notably an absence of studies conducted within the UK in the past five years. There was also a lack of research focused specifically on interventions to improve access to, or affordability of the internet and digital technologies to overcome digital exclusion. None of the included studies specifically aimed to address digital exclusion as such. Due to the paucity of UK-based research on this topic area, and the absence of qualitative data to assess participant acceptability, we cannot be certain that the interventions, irrespective of location or delivery method, would be applicable to the UK context.

The stakeholders involved in this rapid review initially expressed a desire for the review team to identify interventions to improve access to digital services in social care. However, no research in this area was identified, which highlights a further evidence gap.

Although the findings suggest that ICT training and education interventions can improve digital inclusion in a range of older adult population groups, only a small number of studies performed longer-term follow up to assess whether the intervention effects were sustained over time. Furthermore, no studies specifically addressed language barriers, for example, that may be experienced by people whose first language is Welsh, revealing a further evidence gap.

In addition, there is gap in the evidence due to the paucity of cost-effectiveness analyses. Only one study included in this rapid review analysed intervention cost-effectiveness, which investigated an ICT training intervention in the Netherlands. Despite reporting positive outcomes however, we cannot comprehensively assess the cost-effectiveness of such interventions due to insufficient evidence.

The evidence is limited by low quality research with a high risk of bias. Of the 21 included studies, most (n=18) were quasi-experimental, employing either control or comparator groups, or pre-post measures to assess intervention effectiveness. The majority of these studies evaluated the effectiveness of interventions using a pre-post design. Of those that utilised more robust methodology, those with an intervention and control group, few undertook between group comparisons and only investigated within group differences. Only three studies were RCTs, highlighting a paucity of studies utilising robust experimental methods to determine the effectiveness of interventions address digital exclusion in older adults. In addition, follow-up data was rarely collected, so sustainability of the intervention effectiveness is largely unknown.

Critical appraisal of included studies identified the quasi-experimental studies were limited by the absence of a control in several studies (n=8), and by uncertainty regarding the reliability of outcome measurement (n=13). Study quality of RCTs was limited by insufficient reporting on concealed allocation of participants and loss to follow up as well as uncertainty regarding the reliability of outcome measures used. Therefore, given the quality of the evidence base, it is not possible to draw definitive conclusions regarding the effectiveness of the educational interventions included in this rapid review.

### 3.3 Strengths and limitations of this Rapid Review

The studies included in this rapid review were systematically identified through an extensive search of electronic databases and grey literature. We also used a variation of the systematic review approach, abbreviating or omitting some components following established guidelines in an attempt to capture all relevant publications with minimal risk of bias in a timely manner. However, it is possible that additional eligible publications may have been missed, which may bias the rapid review findings. Digital Communities Wales have conducted a range of case studies to support digital inclusion (Welsh Government 2022), for example providing digital devices to care homes across Wales (Welsh Government 2020). However, as they were not published within peer-reviewed journals and often lacked full evaluations, our ability to quality appraise the methodology and findings was limited and made them ineligible for inclusion in this rapid review.

Every effort was made to conduct a robust synthesis of study findings; however, the evidence is limited by poor reporting of intervention methods and results in several studies, which made interpretations challenging. However, each stage of the review was consistency checked for accuracy, and issues were discussed within the team.

We used a broad definition of digital literacy to inform our inclusion eligibility, a strength as it enabled the identification and inclusion of a wide range of research addressing various facets of digital literacy. However, this broad approach limited the comparability of findings across studies due to the varied terminology, definitions and measurements used to assess different outcomes. Study findings were also commonly self-reported and were not obtained using objective measures, limiting our ability to make firm inferences relating to intervention effectiveness.

### 3.4 Implications for policy and practice

Overall, the evidence identified suggests educational interventions broadly support improved digital inclusion among the older population. Findings identified here could inform the development and delivery of future interventions. However, it is important to consider the context in which the included interventions were used and the lack of certainty of the findings.

Lower income and SES, as well as costs of acquiring and using technology are established barriers to digital inclusion (Chen et al, 2022 & Moroney et al, 2020). To achieve improved and sustained digital inclusion in older adults, evidence suggests it may be important to ensure structural barriers, such as access to the internet and affordability of devices are removed. However, it is unclear what the cost implications may be to deliver this, or if these barriers could be reduced by raising awareness of social tariffs available in the UK for those receiving pension credit (Ofcom 2024). In practice, providing ICT education and training to older adults without considering these barriers may exacerbate existing inequalities. In addition, it is also important to consider that digital devices and services are continually evolving, which suggests education and training may need to be ongoing to ensure sustained digital literacy and technology use.

Although study participants generally reported that they appreciated and benefitted from the educational interventions, it is important to consider that older adults retain the right to choose whether or not to interact with essential services physically (offline) or digitally. The Digital Strategy for Wales highlights the need to equip individuals of all ages with the motivation, access, skills and confidence to engage with digital technologies when they want to (Welsh Government 2021). There are also several barriers to engaging with the digital world that are particularly pertinent within the older adult population, including fear, anxiety, stereotypes, and stigmas, as well as costs or disabilities. The different age groups within the older adult population should also be considered for example working older adults may have different need to older adults who have retired. Services undergoing digitisation may need to find ways to encourage and support older adults to engage. However, alternative methods of accessing these services should remain available where possible or it may exacerbate exclusion by leaving some people behind.

### 3.5 Implications for future research

Our rapid review identified a paucity of UK based research in this area, indicating a need for future investigations within the UK to comprehensively assess the effectiveness and generalisability of interventions, particularly within the Welsh context. Most studies identified were quasi-experimental in design, and the paucity of RCTs shows more, higher-quality research is possible and also needed to further advance this topic area. The review identified a number of multi-component, complex interventions developed to address the many inter-related aspects of supporting elderly people (or vulnerable subgroups) and increase digital inclusion. However, these were mainly evaluated using pre-post study design and further well designed research is needed to rigorously evaluate their effectiveness and identify which components are most impactful.

It may be beneficial for researchers to adopt standardised definitions and measurement tools, promoting greater consistency and comparability in study findings in relation to digital inclusion. If future research is conducted within the Welsh context, consideration should be given to the Welsh Minimum Digital Living Standard set out by the Welsh Government (MDLS) (Welsh government 2023c). This standard highlights the need to support individuals not only to access the internet or digital devices and develop digital skills, but also to improve their ability to engage online safely and confidently. Future research should consider how improving older adults’ digital skills such as learning how to use email may not necessarily improve their ability to identify risks such as phishing emails potentially putting them at increased risk.

Further research is needed to strengthen the evidence base of digital inclusion in vulnerable older adults to better understand the effectiveness of educational interventions in these population groups. This is important because certain population groups are likely to be at greater risk of digital exclusion than others. Furthermore, there is a need for greater research which focuses on overcoming language barriers when addressing digital exclusion in older adults; a key challenge highlighted by stakeholders in the Welsh context.

There is also a need for more research specifically aimed at improving access to digital devices and services. Additionally, UK based research focussing on digital inclusion in older adults accessing social care services is needed, as no such research was identified. Without this, the digital transition of social care services may be hindered and could potentially exacerbate digital exclusion among those who need to access it without careful consideration of it’s users.

Lastly, further evidence is needed to determine if digital exclusion can be addressed in a cost-effective way and to determine potential cost savings in both the general older adult population as well as the vulnerable sub-populations within it.

#### 3.6 Economic considerations*

- It is important that the relative cost-effectiveness and acceptability of digital versus traditional in-person social care solutions continue to be investigated. With a particular focus given to older people in which digital exclusion is more prevalent than in other age groups.
- There is a larger existing evidence base concerning reducing digital exclusion in healthcare solutions than social care solutions.
- There is some systematic review evidence since 2020 that simple, text-based interventions enabling communication with social care services for older adults is cost-effective (Ghani et al., 2020).
- Further research is required that considers the role of family/informal carers assisting older individuals accessing social care through digital means.
- Technological solutions to improve patient data flows between health and social care in England have been found to provide monetary benefits over and above the initial investment. Benefits to patients through the introduction of improved data linkages and comprehensive digital health records included promotion of self-care/independent living, improved management of long-term conditions and supporting the reduction of long-term care needs (NHS Digital, 2021). There exist significant barriers to older people regarding the use of digital solutions at a whole system level (Basis Research, 2022).
- £2.4 billion** is spent on adult social care each year in Wales (Social Care Wales, 2018). Demographic changes including greater life expectancies and increases in individuals presenting with complex care needs have increased pressures on adult social care sector resources (UK Parliament, 2024).

*\*This section has been completed by the Centre for Health Economics & Medicines Evaluation (CHEME), Bangor University*

*\*\*Figure inflated from 2016 prices to 2024 prices using the Bank of England Inflation Calculator (https://www.bankofengland.co.uk/monetary-policy/inflation/inflation-calculator)*

## Data Availability

All data produced in the present study are available upon reasonable request to the authors

## Abbreviations

Acronym: Full Description
CAS: Computer Anxiety Scale
C&C: Digital Communication and Collaboration
CI: Confidence Interval
COVID-19: Coronavirus disease 2019
CPQ: Computer Proficiency Questionnaire
CTN: Community Tech Network
D-AI: Dutch Activity Inventory
DILE: Digital Literacy Evaluation Tool
eHEALS: The 8-item eHealth Literacy Scale
GSE: General Self-Efficacy Scale
GTF: Good Things Foundation
HSL-12: 12-question Health Literacy Scale
ICER: Incremental Cost-Effectiveness Ratio
I-CHATT: Individualized Community and Home-based Access to Technology Training
ICT: Information and Communication Technology
I&DL: Information and Data Literacy
IF: Intergenerational Forum
IMU: Intergenerational Mentor-up
JBI: Joanna Briggs Institute
LBFE: Little Brothers – Friends of the Elderly
LMS: Learning Management System
Madj: Adjusted Mean Confidence Score
MDPQ-16: 16-question Mobile Device Proficiency Questionnaire
ND: Motivation
OITO: Oficinas de Inclusão Tecnolόgica Online, “Workshops for Online Technological Inclusion”
OR: Odds Ratio
PC: Personal computer
PRISM: Personal Reminder Information and Social Management system
QALY: Quality-Adjusted Life-Year
RCT: Randomised Controlled Trial
RR: Rapid Review
SD: Standard Deviation
SDLE: Senior Digital Literacy Evaluation
SES: Socio-Economic Status
SOTU: Survey Of Technology Use
UK: United Kingdom
USA: United States of America
WRAT: Wide Range Achievement Test

## 5. RAPID REVIEW METHODS

We searched for primary sources to answer the review questions: What evidence exists on the effectiveness of interventions to address digital exclusion in older adults?

### 5.1 Eligibility criteria

**Table 4:** Eligibility criteria.

|  | **Inclusion criteria** | **Exclusion criteria** |
| --- | --- | --- |
| **Participants** | Older adults aged 60 years or over as defined by the Older People's Commissioner for Wales (Older People's Commissioner for Wales 2023)<br>Carers of older adults aged 60 years or over | Adults less than 60 years of age<br>Children |
| **Settings** | Any setting except health interventions | Hospital settings |
| **Intervention / exposure** | Interventions to address digital exclusion | Interventions to assess the use of telemedicine or to improve health literacy |
| **Comparison** | If applicable, no intervention or alternative interventions |  |
| **Outcomes** | Outcomes(s):<br>• Effectiveness of interventions to address digital exclusion (in relation to access, ability or affordability)<br>• Digital literacy skills<br>• Technology use<br>• ICT acceptance<br>• Views and perceptions of the intervention<br>• Technological knowledge<br>• Self-efficacy |  |
| **Study design** | Any primary studies that:<br>• Assess the effectiveness of interventions to address digital exclusion (comparative) | Secondary/Tertiary research |
| **Countries** | All countries |  |
| **Language of publication** | English |  |
| **Publication date** | 2018-2023 |  |
| **Publication type** | Published | Commentaries, editorials, letters, conference abstracts, preprints |

### 5.2 Literature search

A search of electronic bibliographic databases including Social Policy and Practice (Ovid), Scopus and Sociology Collection was conducted, as well as supplementary searches performed in Google Scholar, SCIE and The King’s Fund. Citation tracking from secondary sources identified during the preliminary stages was also undertaken. All searches were conducted between 21^st^ and 23^rd^ November 2023. Search concepts and keywords included digital inclusion, digital exclusion, digital skills, elderly, older adults.

### 5.3 Study selection process

All studies were uploaded to the systematic reviewing platform Rayyan and were screened by two independent reviewers. Any conflicts were resolved within the review team.

### 5.4 Data extraction

Data extracted was conducted by a single reviewer and was consistency checked by a second reviewer. Information extracted includes:

- Citation
- Study design
- Intervention
- Comparator
- Study aim
- Data collection methods and dates
- Outcomes reported
- Sample size
- Participants
- Setting
- Key findings
- Observations/Notes

### 5.5 Study design classification

The included studies were classified as quasi-experimental studies or RCTs.

### 5.6 Quality appraisal

The JBI critical appraisal checklists for quasi-experimental studies and RCTs were used to assess the methodological quality of each included study. These checklists are not designed to assign an overall score to each study. For the purposes of this review, a pragmatic system devised and used in a previous rapid review (Weightman et al 2022), was used to assess each study as being of high, moderate, or low quality.

#### High quality

RCT plus a score of 8 or more YES on the JBI RCT check list, plus, no concerns suggesting a medium or high risk of bias in any aspect of the study design.

#### Moderate quality

RCT plus a score of 6 or more YES on the JBI RCT check list OR a score of 8 or more YES on the JBI quasi experimental study checklist, pus, no concerns suggesting a very high risk of bias in any aspect of the study design

#### Low quality

Others

Quality assessment was undertaken in duplicate by two independent reviewers. Any discrepancies were discussed and resolved between reviewers. The quality assessment of individual studies can be seen in section 6.3.

### 5.7 Synthesis

Data was synthesised narratively to provide a collective interpretation of the evidence.

### 5.8 Assessment of body of evidence

An assessment of the overall body of evidence was made based on the relevance of the available evidence in addressing the review question, the amount and quality of the evidence, the magnitude and direction of effects and consistency in the findings. This information is provided for the differing outcomes in Table 3, and was used to classify the body of evidence for each outcomes as: Very low quality; Low quality; Moderate quality; and High quality.

## 6. EVIDENCE

### 6.1 Search results and study selection

A total of 1,540 records were retrieved which were managed in Endnote 20. Following deduplication, 1,307 records remained. The search strategy used to search Social Policy and Practice is available in Appendix 1. A total of 68 articles were screened at full text by two independent reviewers, and any conflicts were discussed and resolved within the team. A visual representation of the flow of studies throughout the review can be found in Figure 6.1.

**Figure 1.**
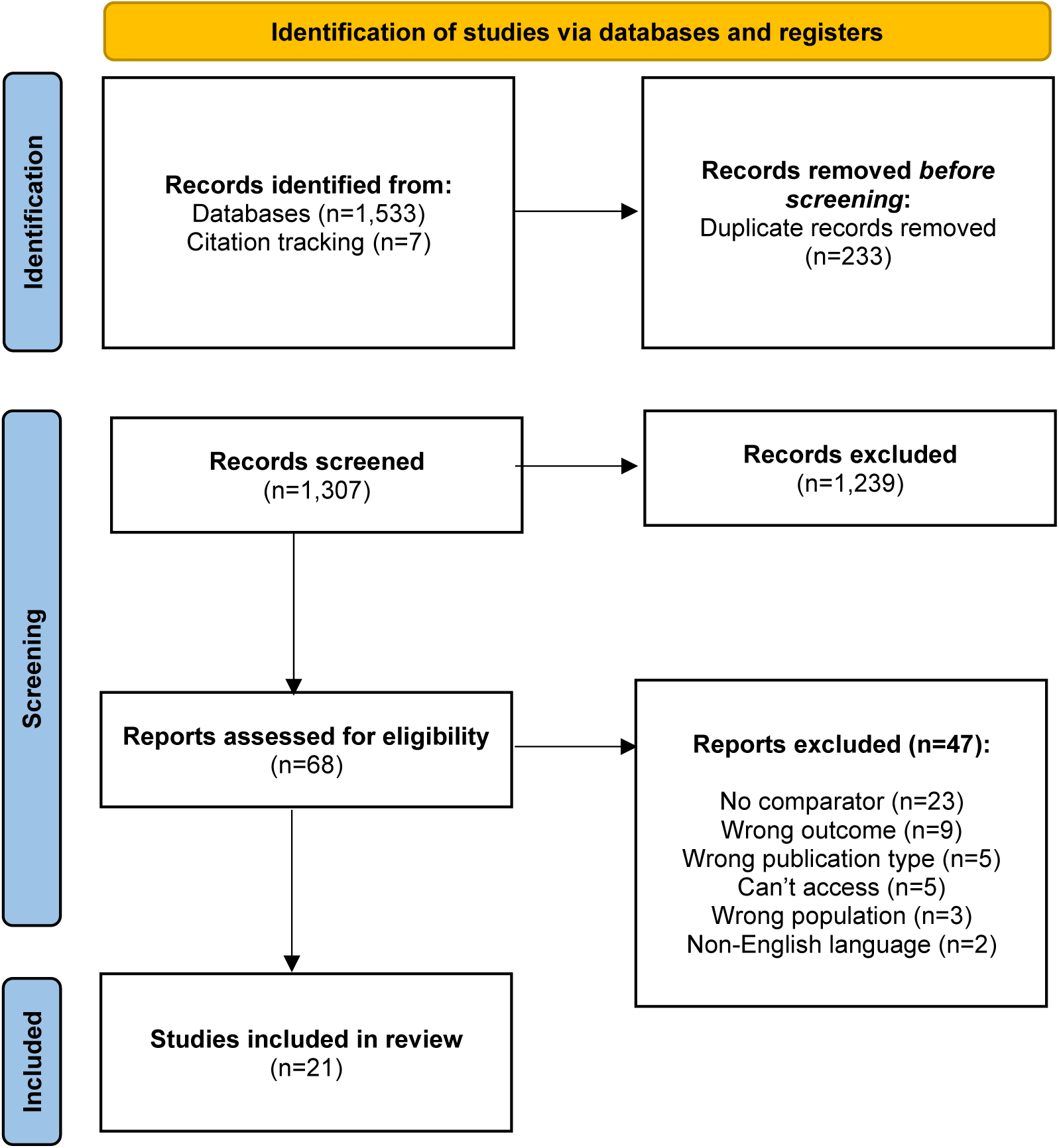
PRISMA flow diagram.

### 6.2 Data extraction

**Table 5:**
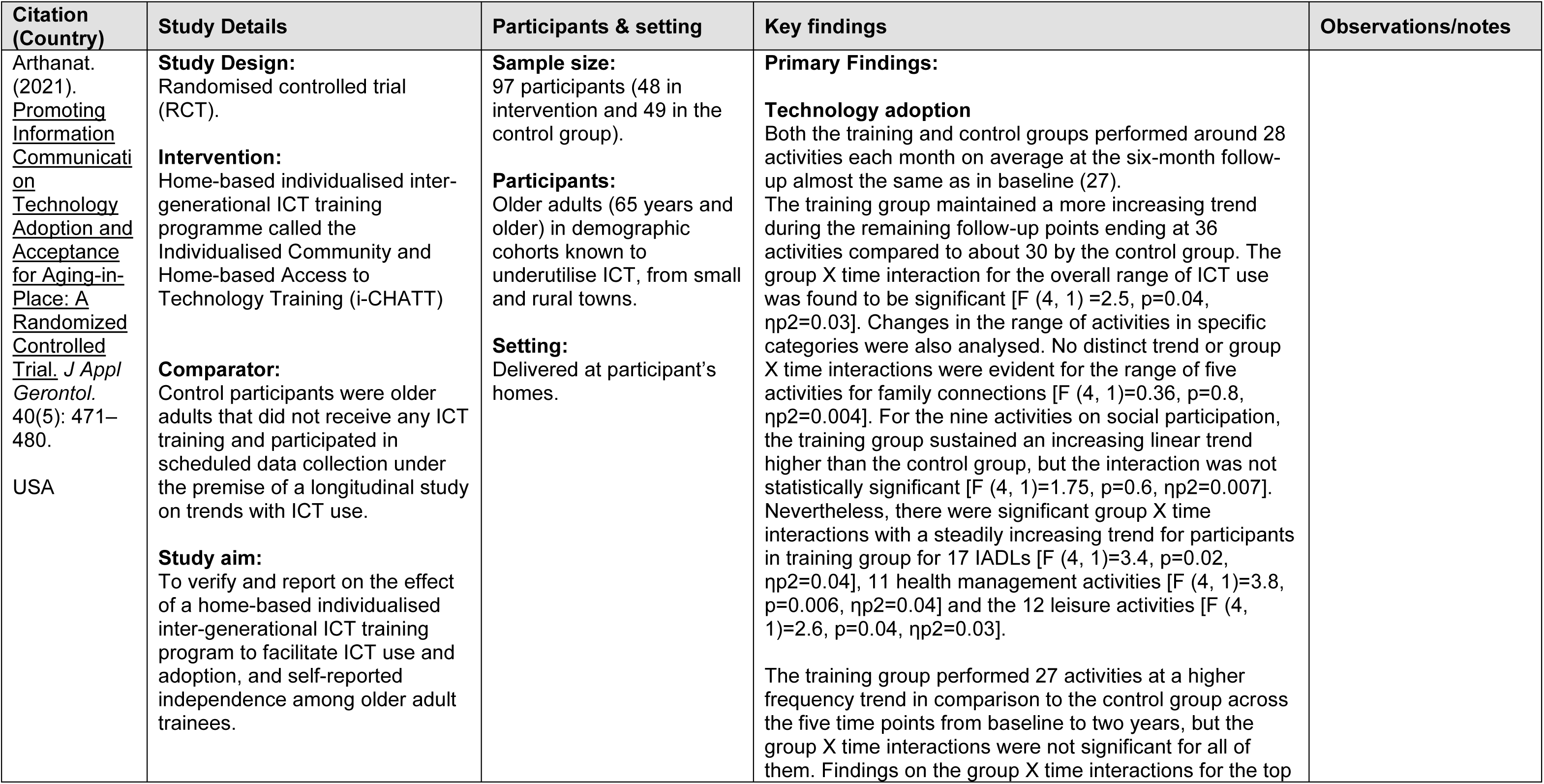
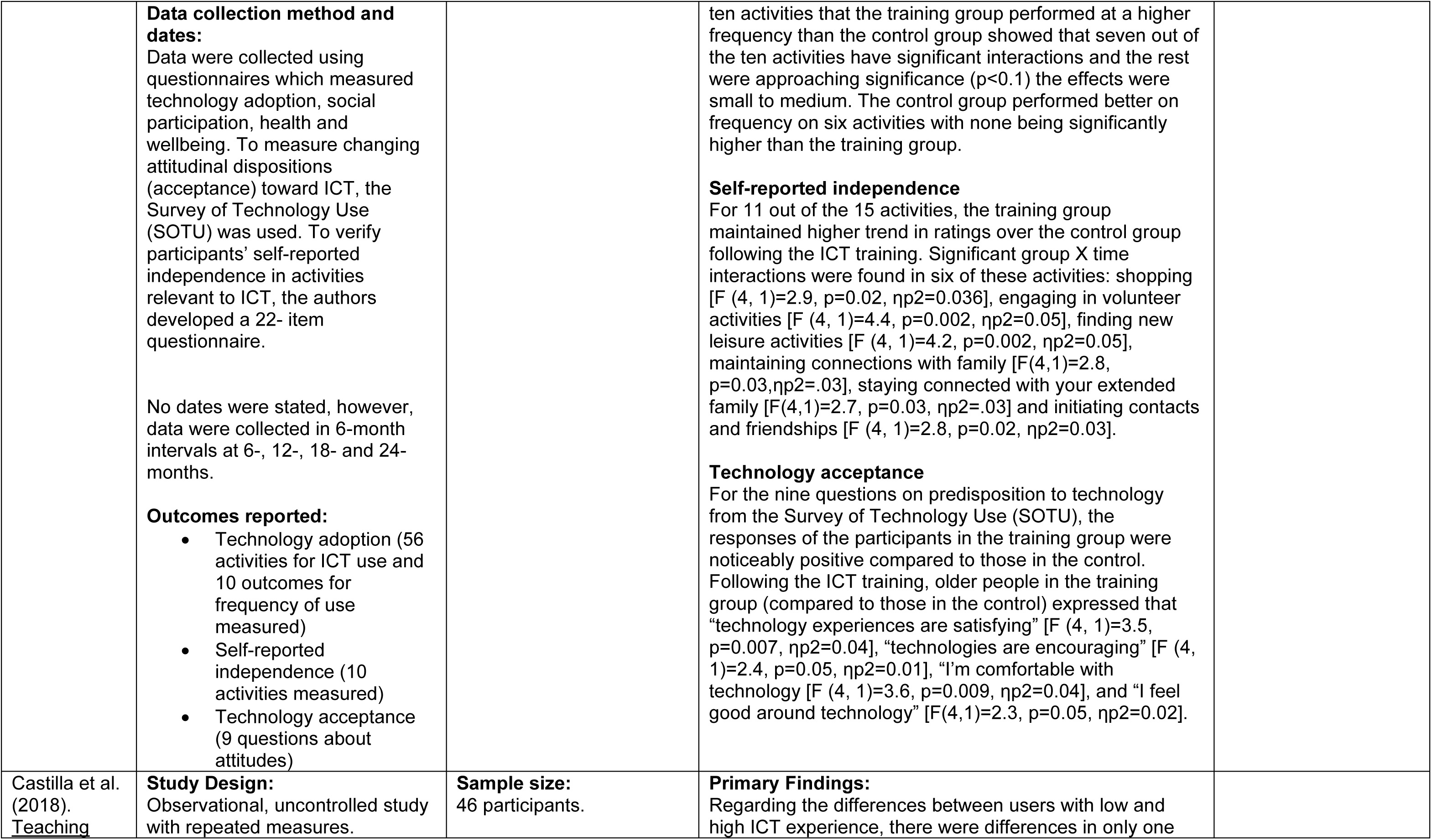
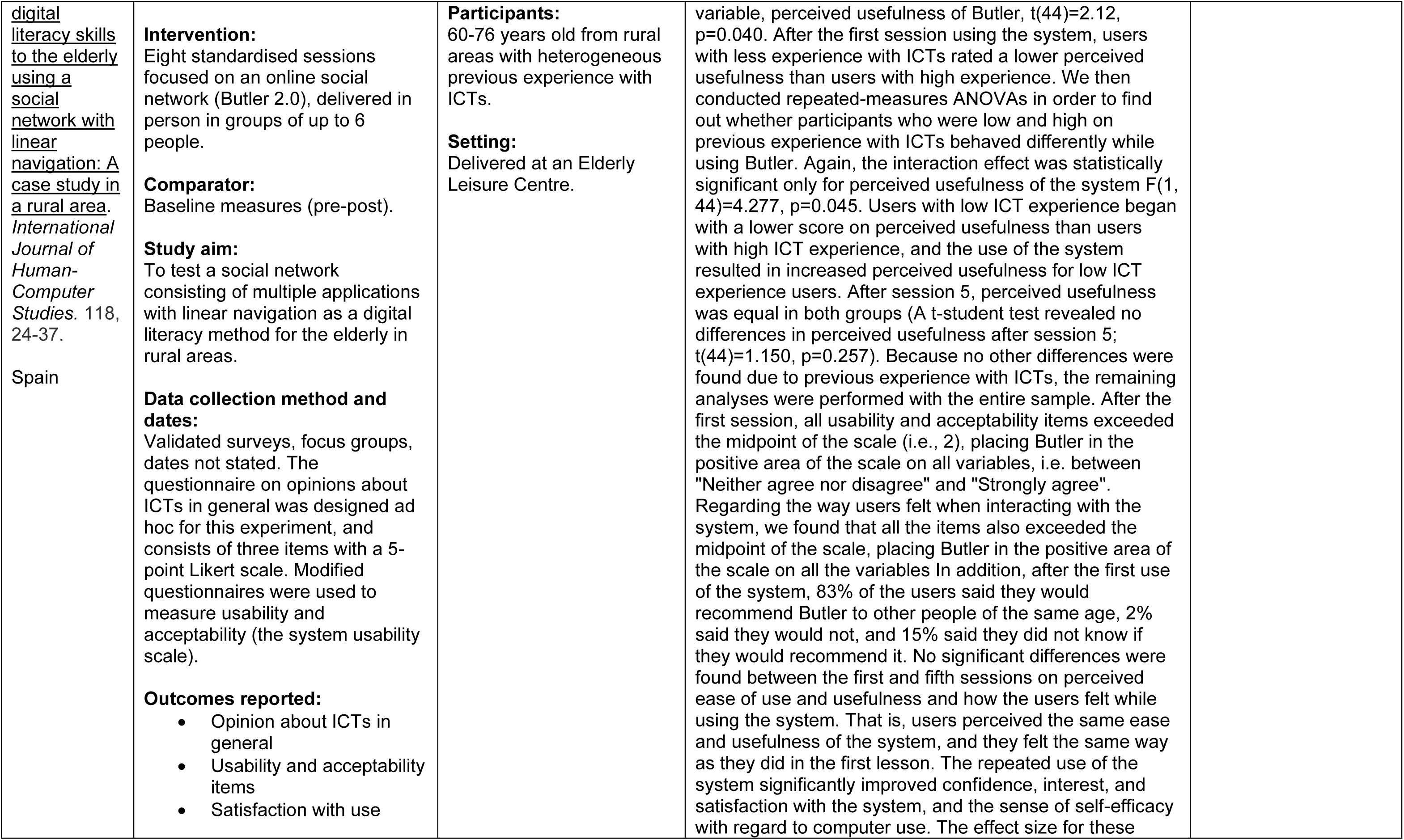
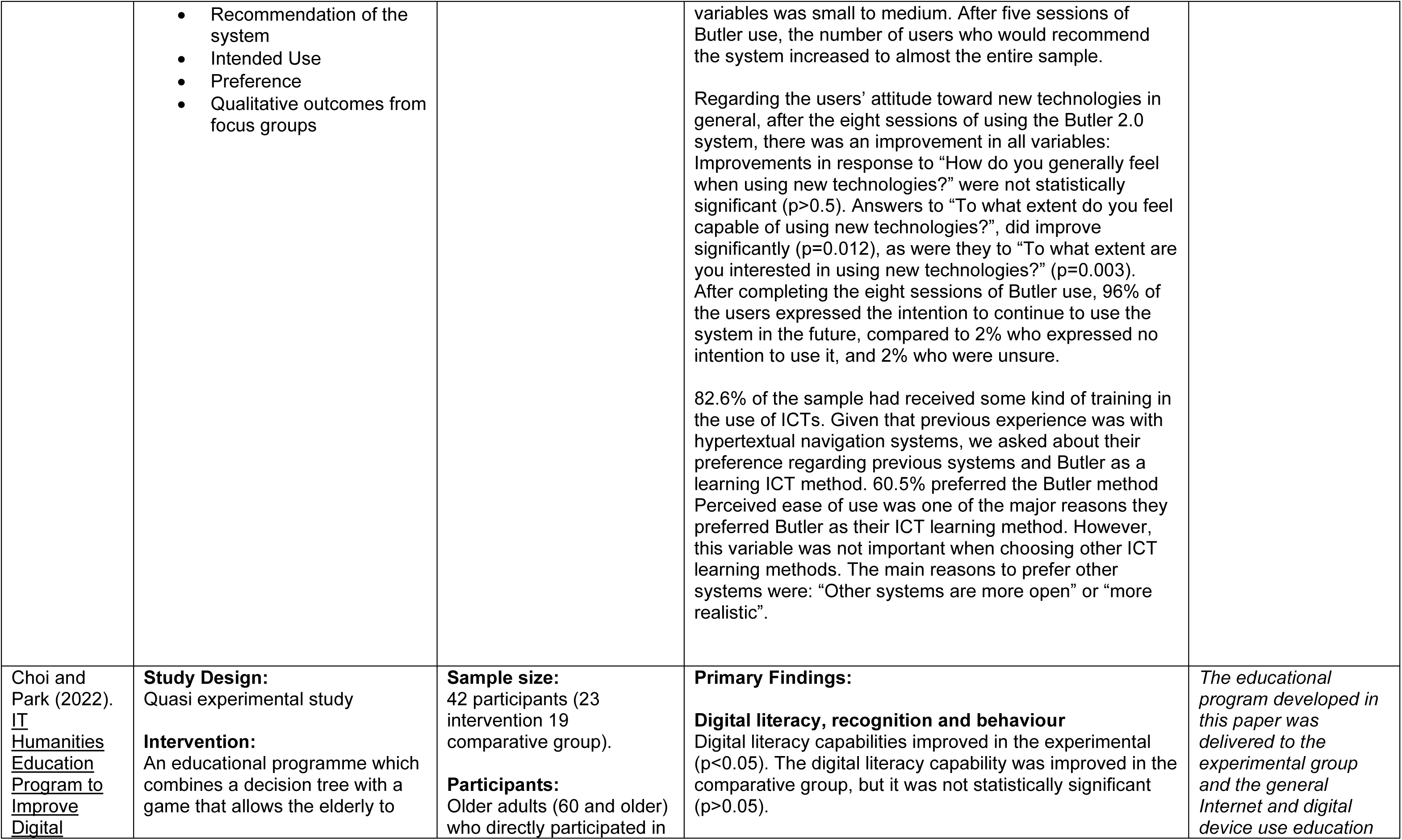
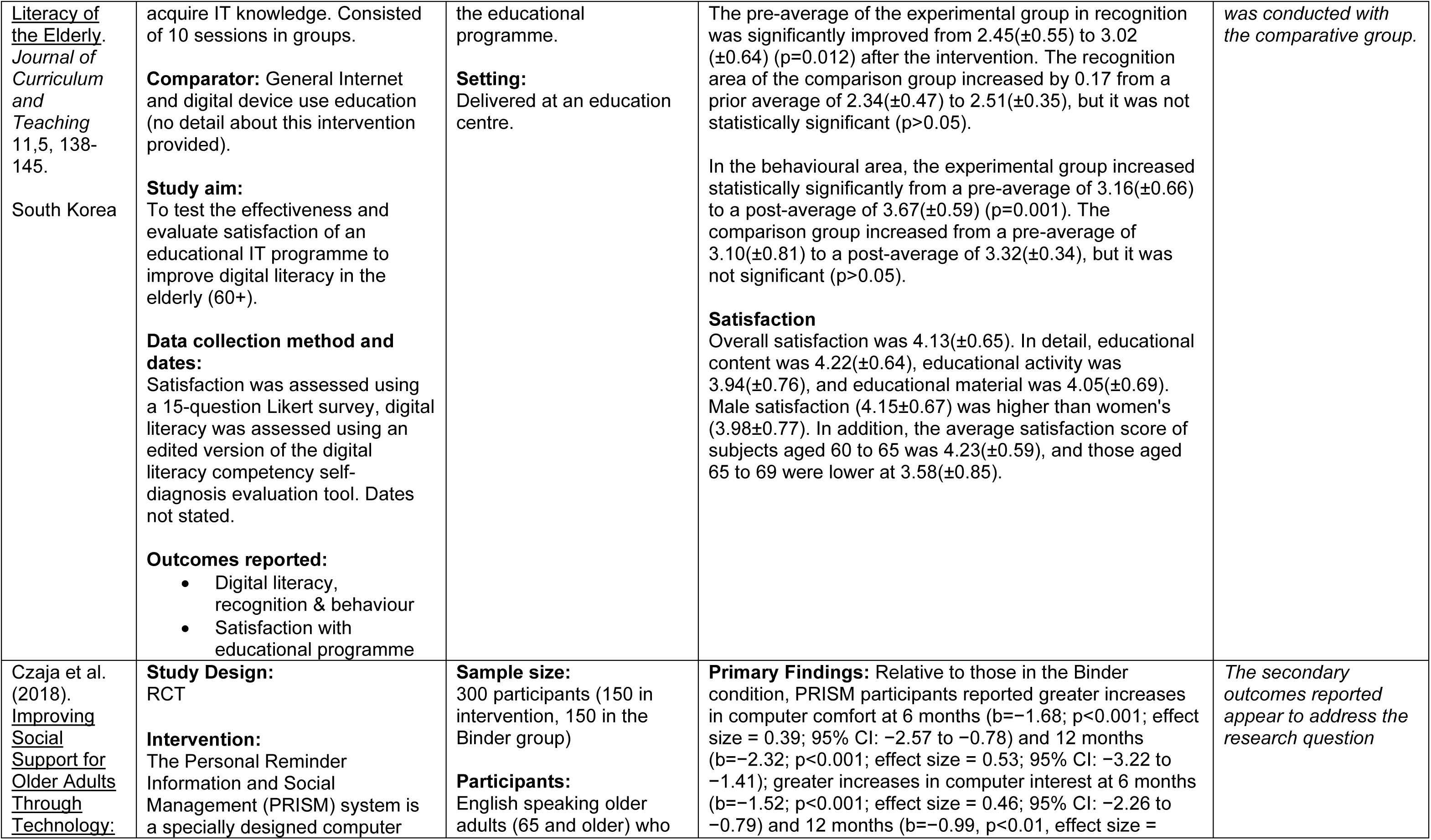
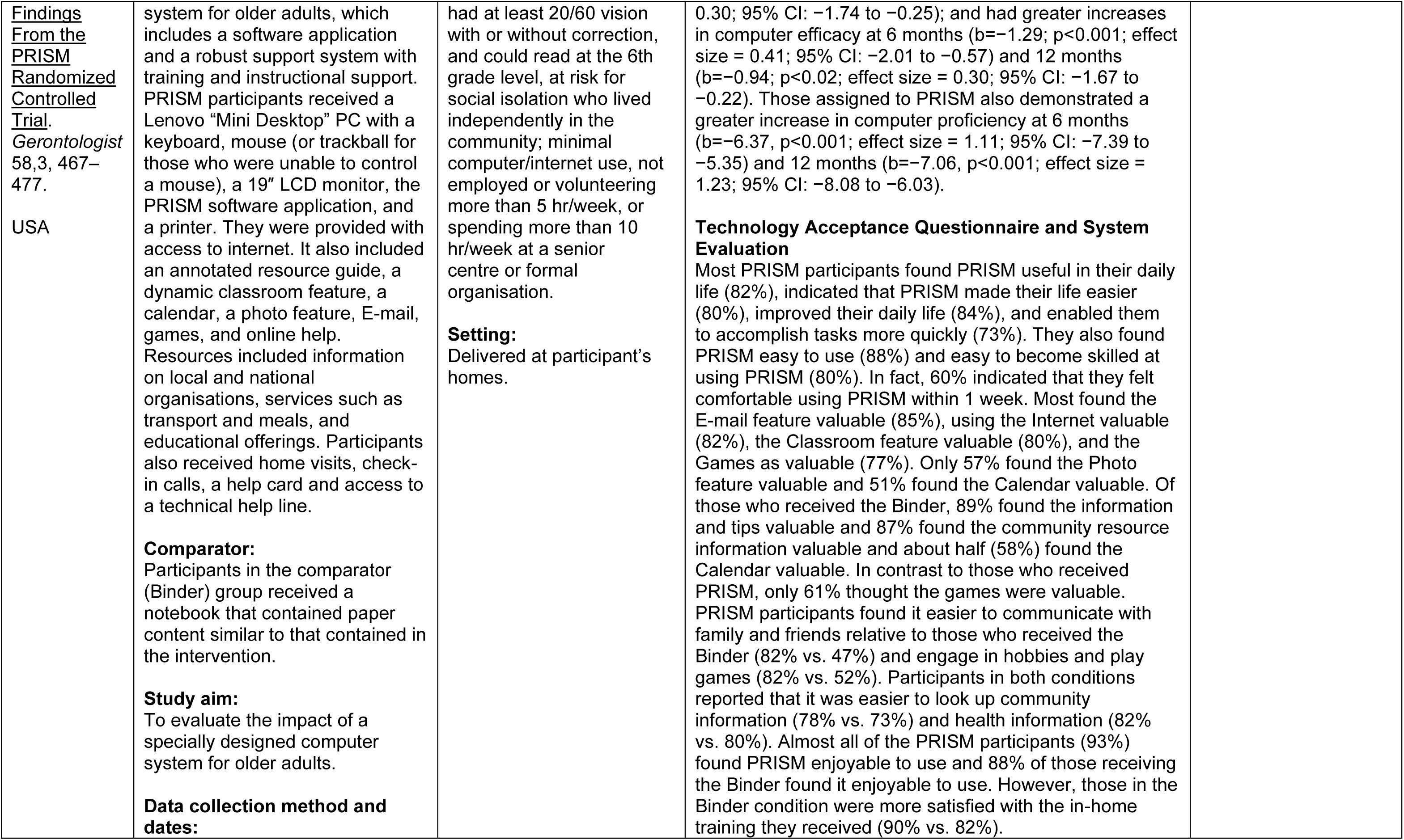
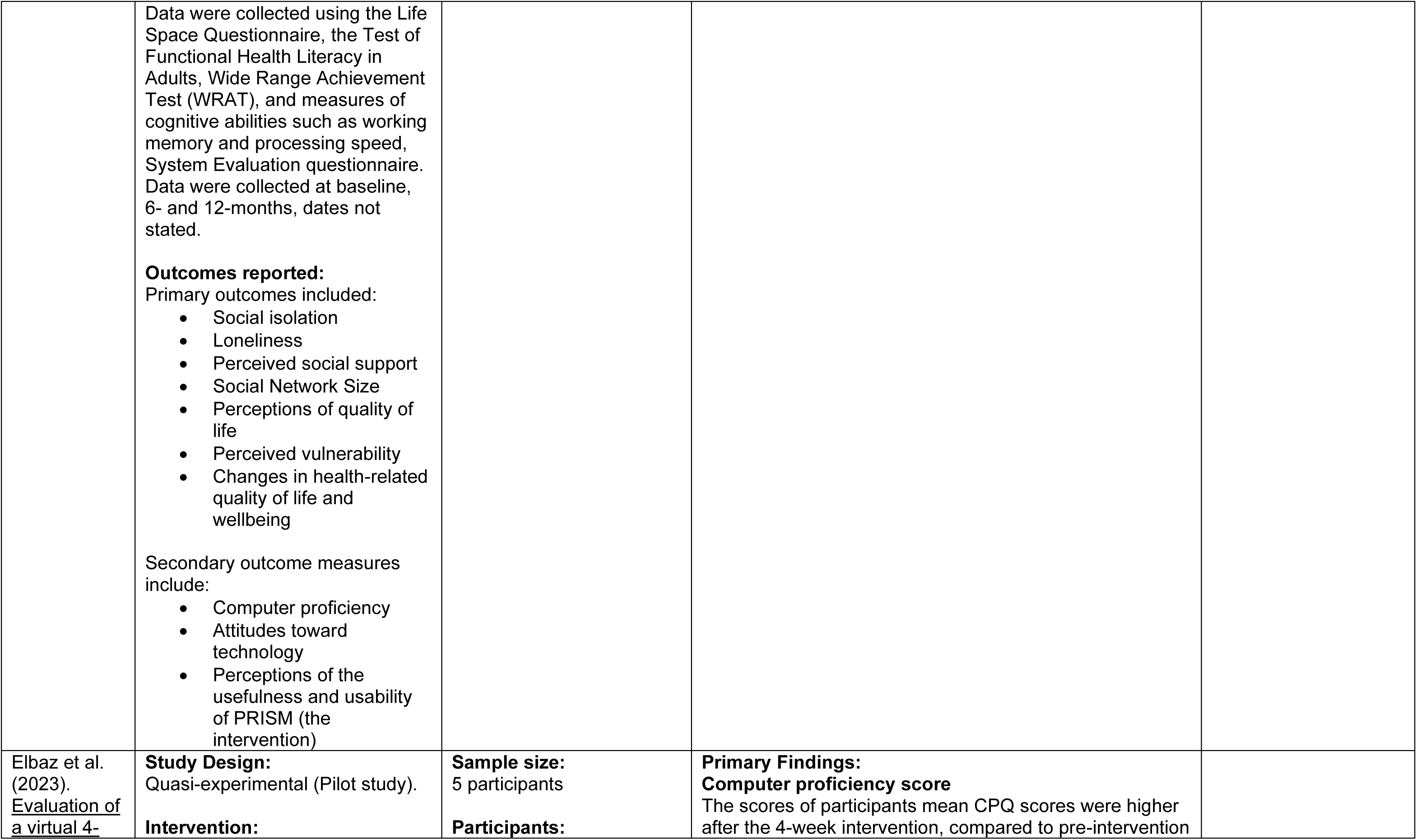
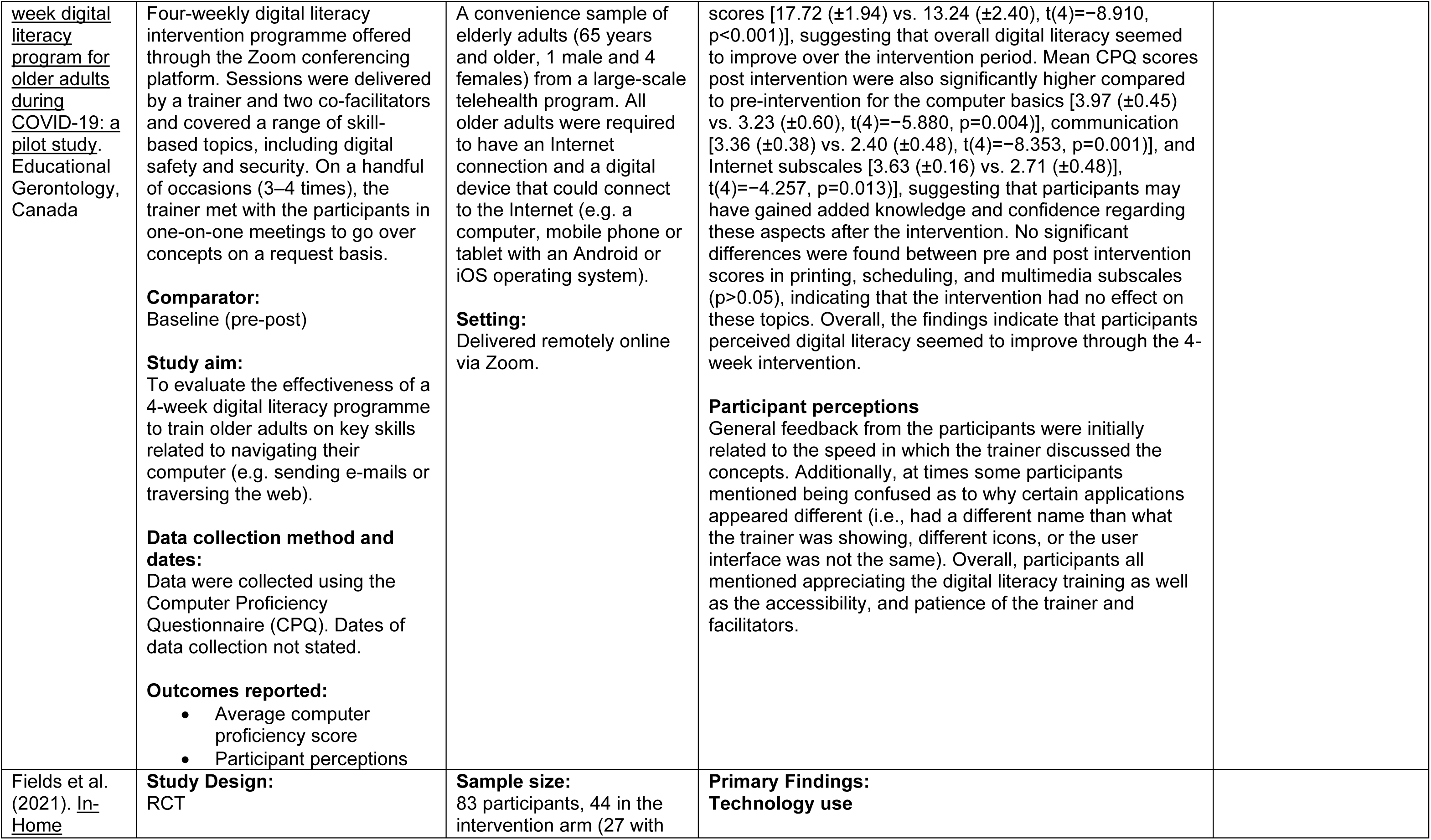
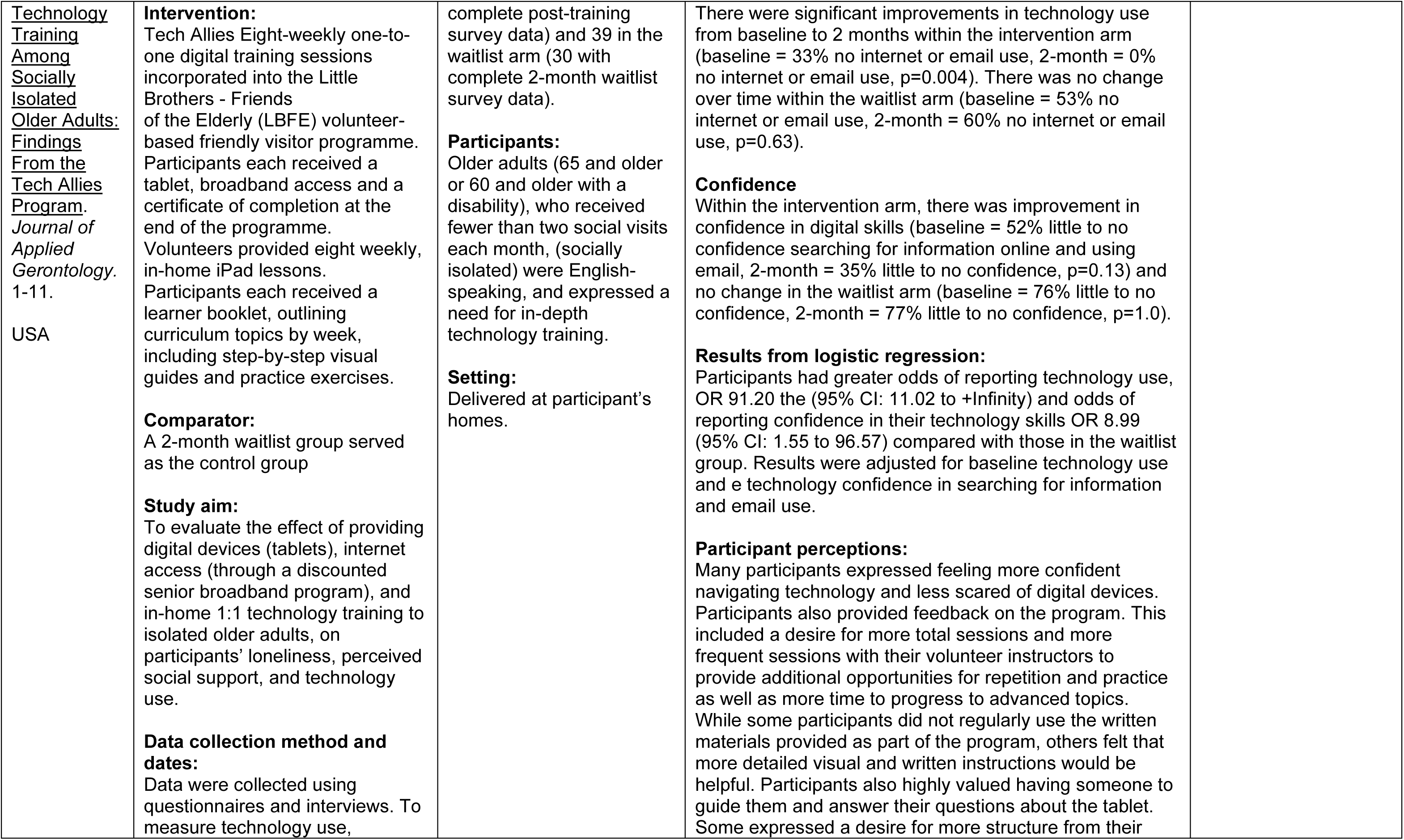
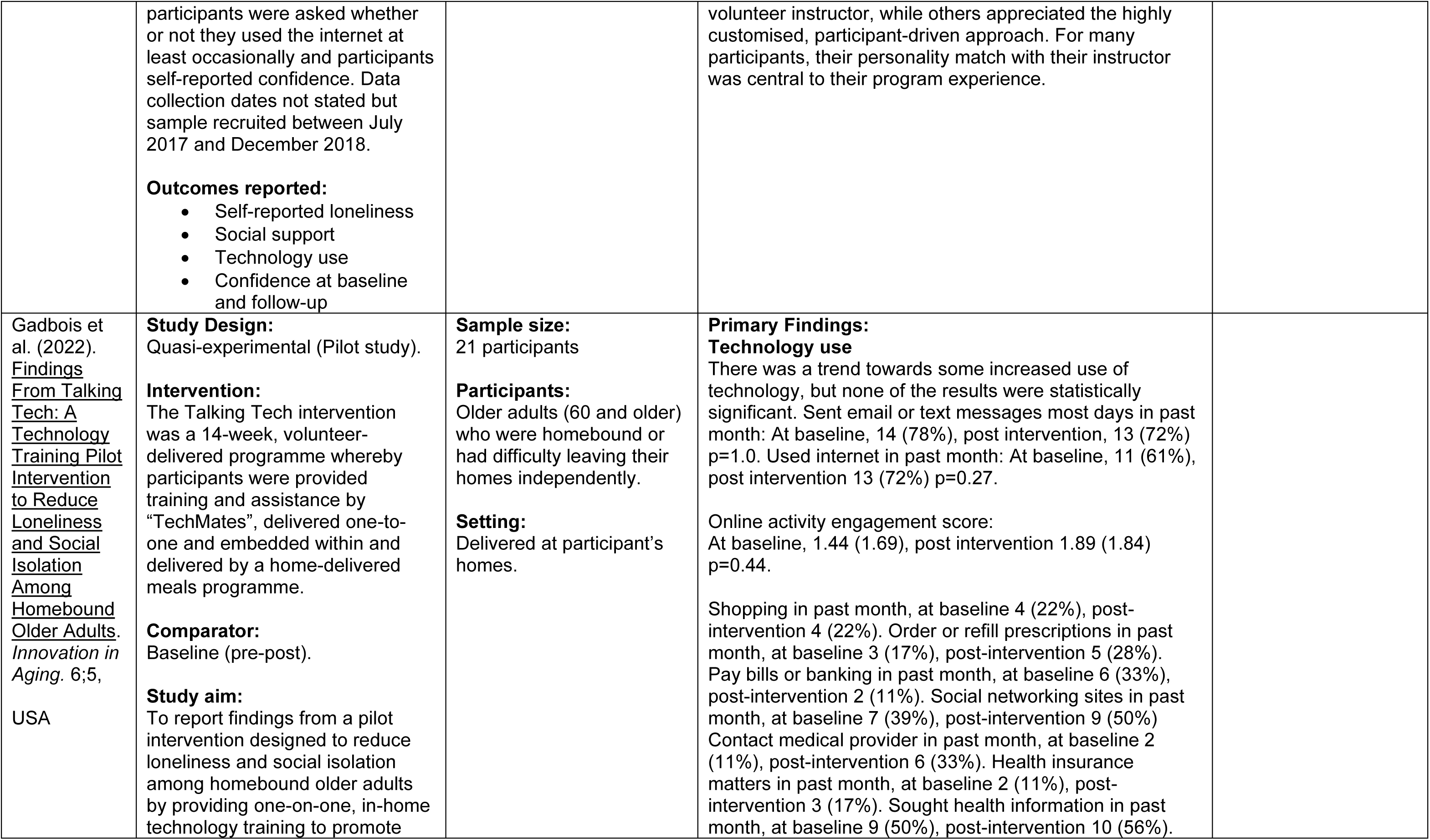
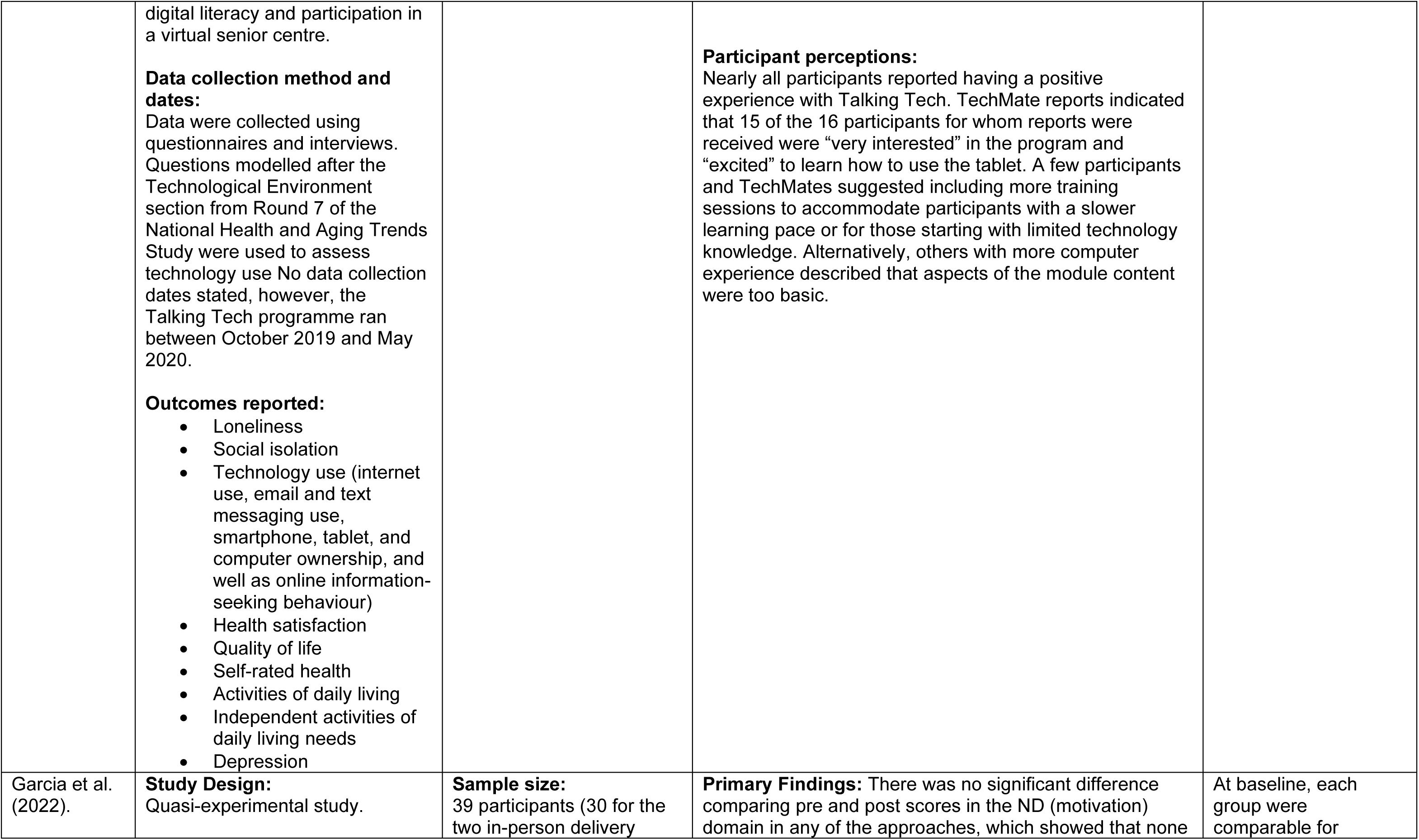
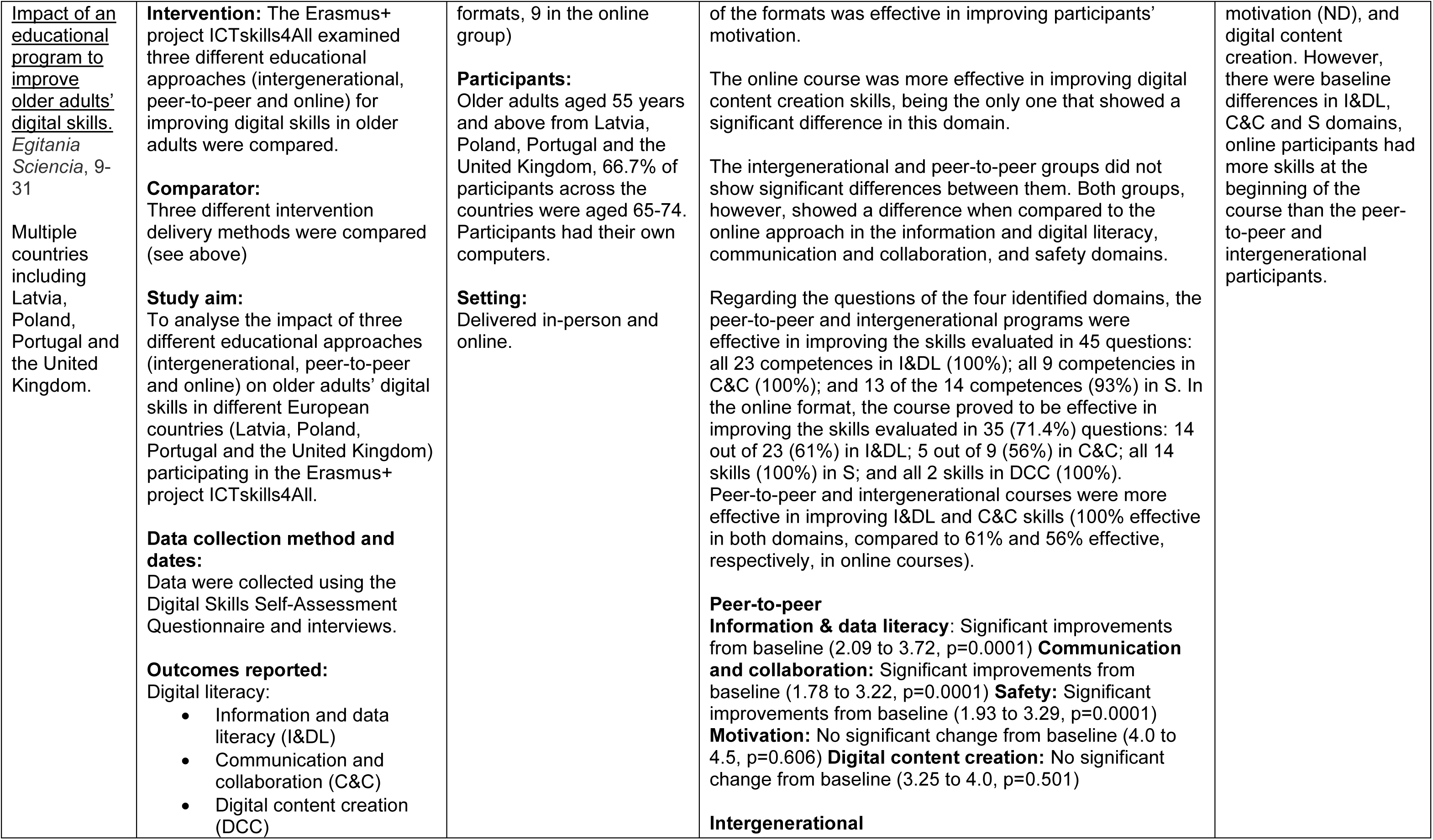
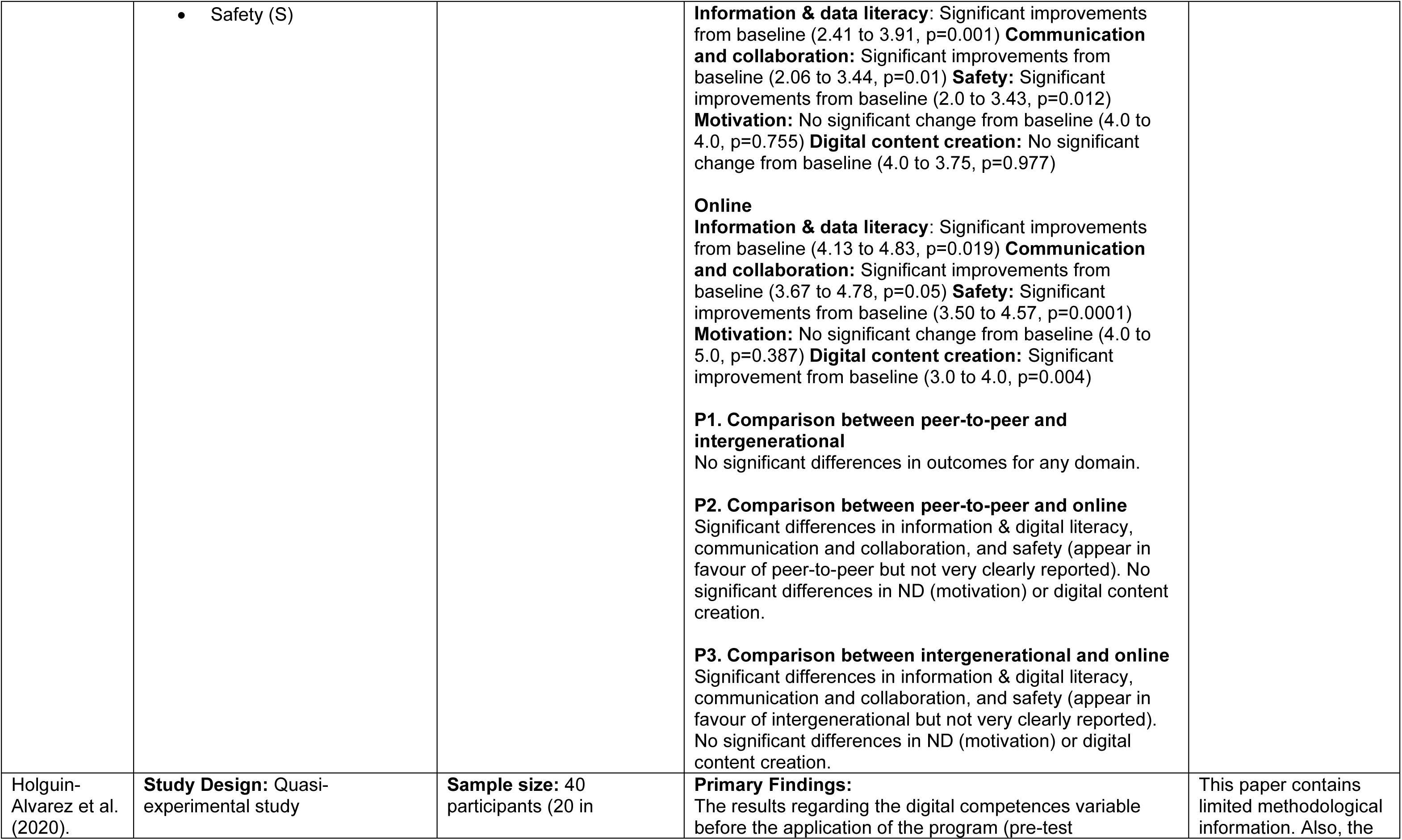
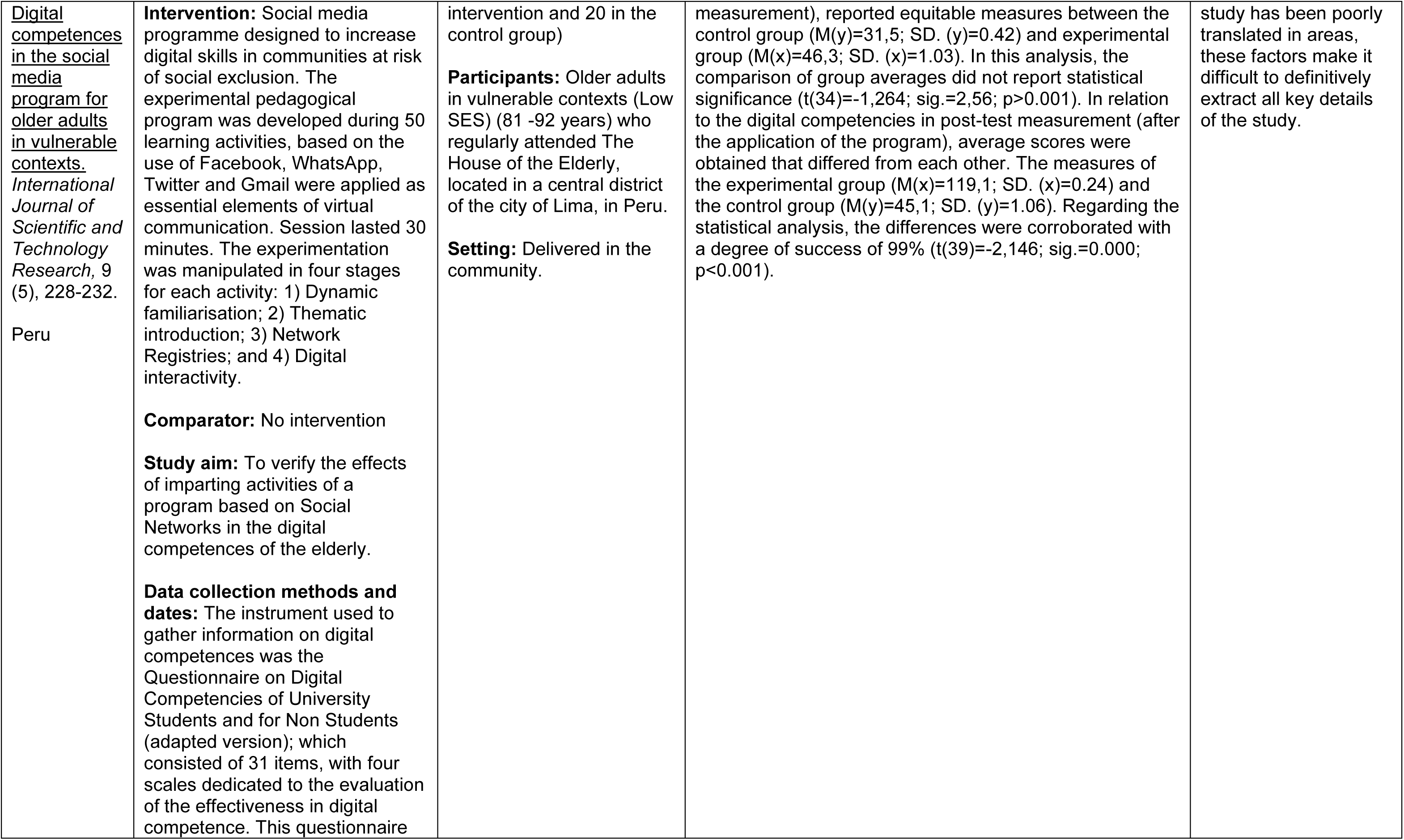
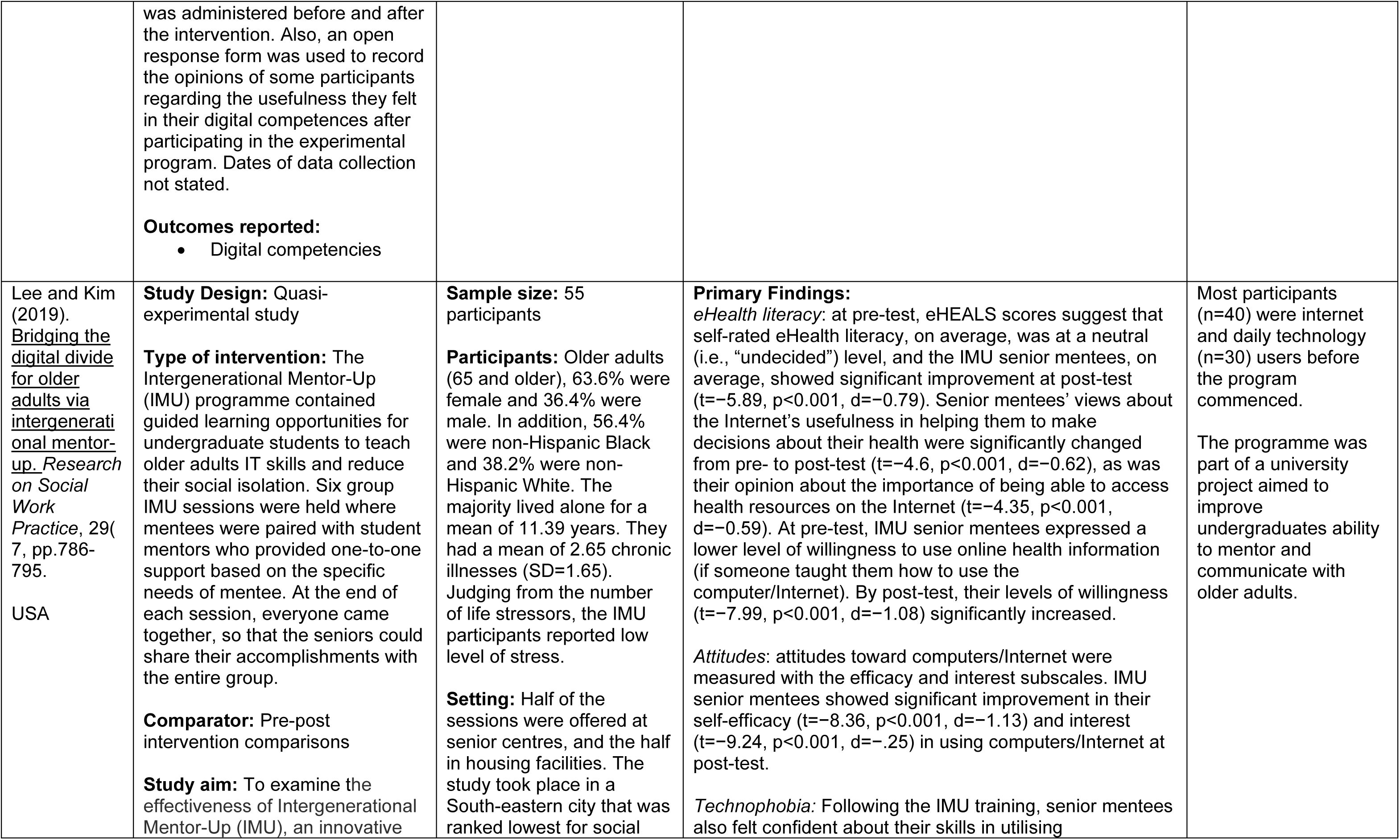
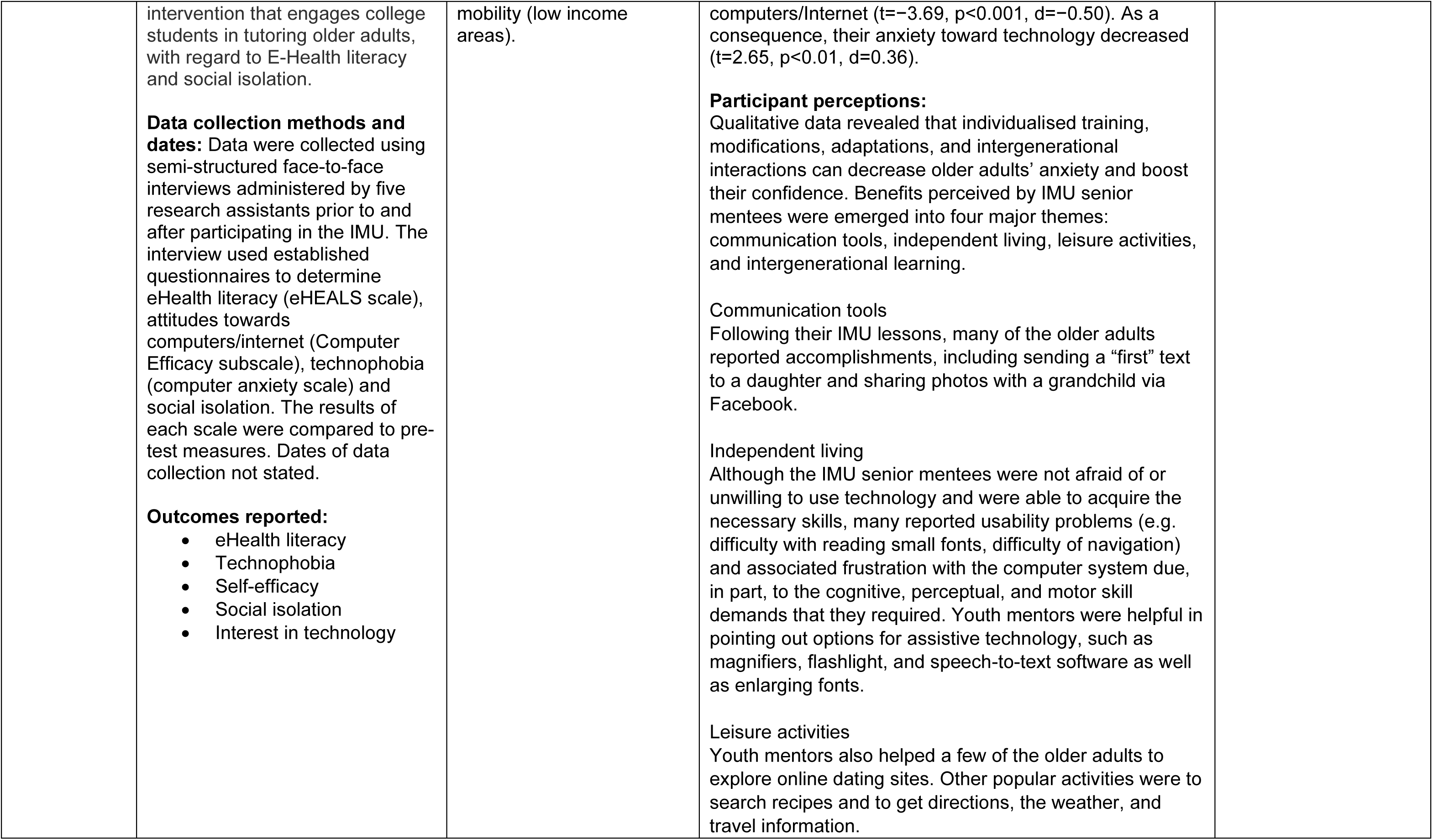
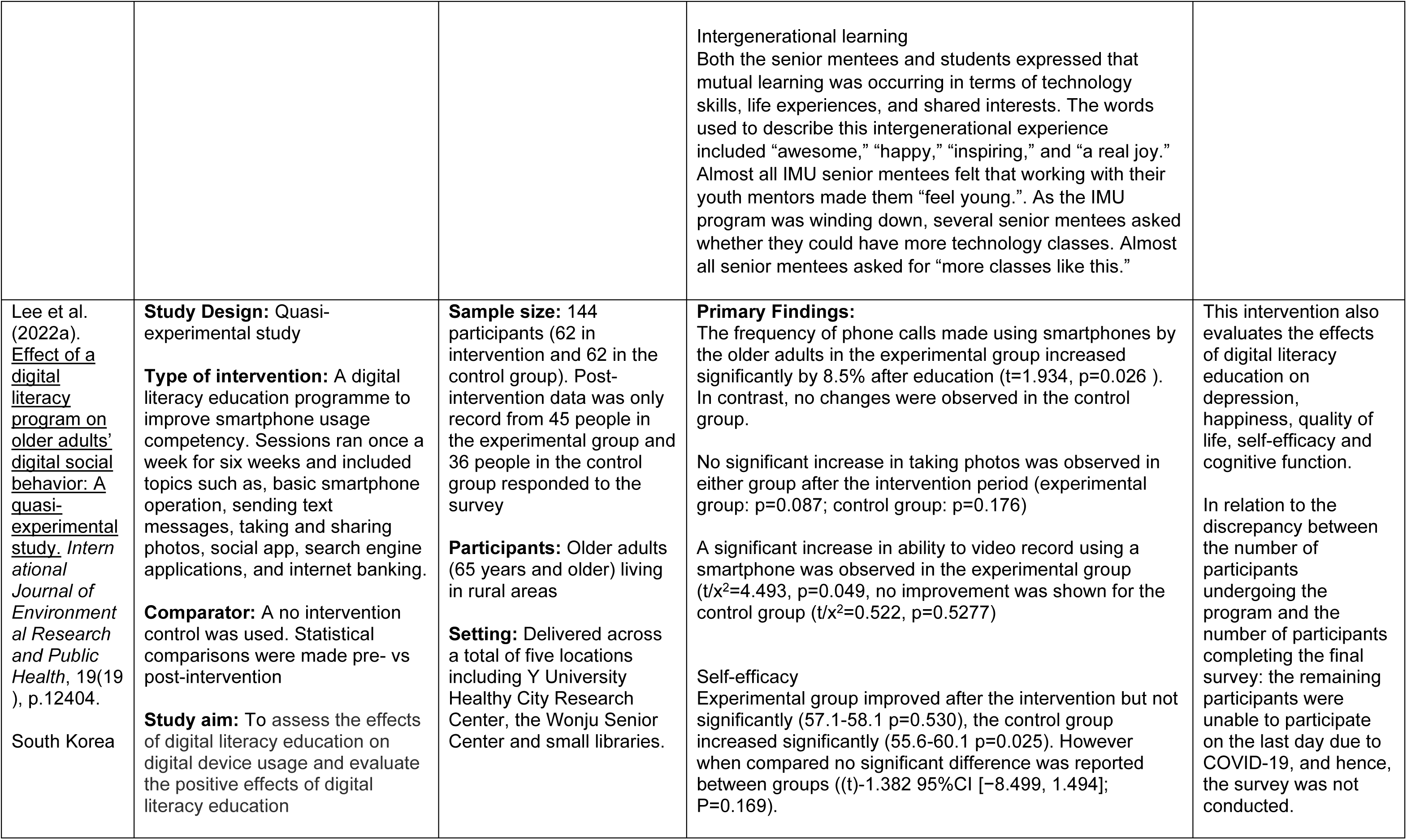
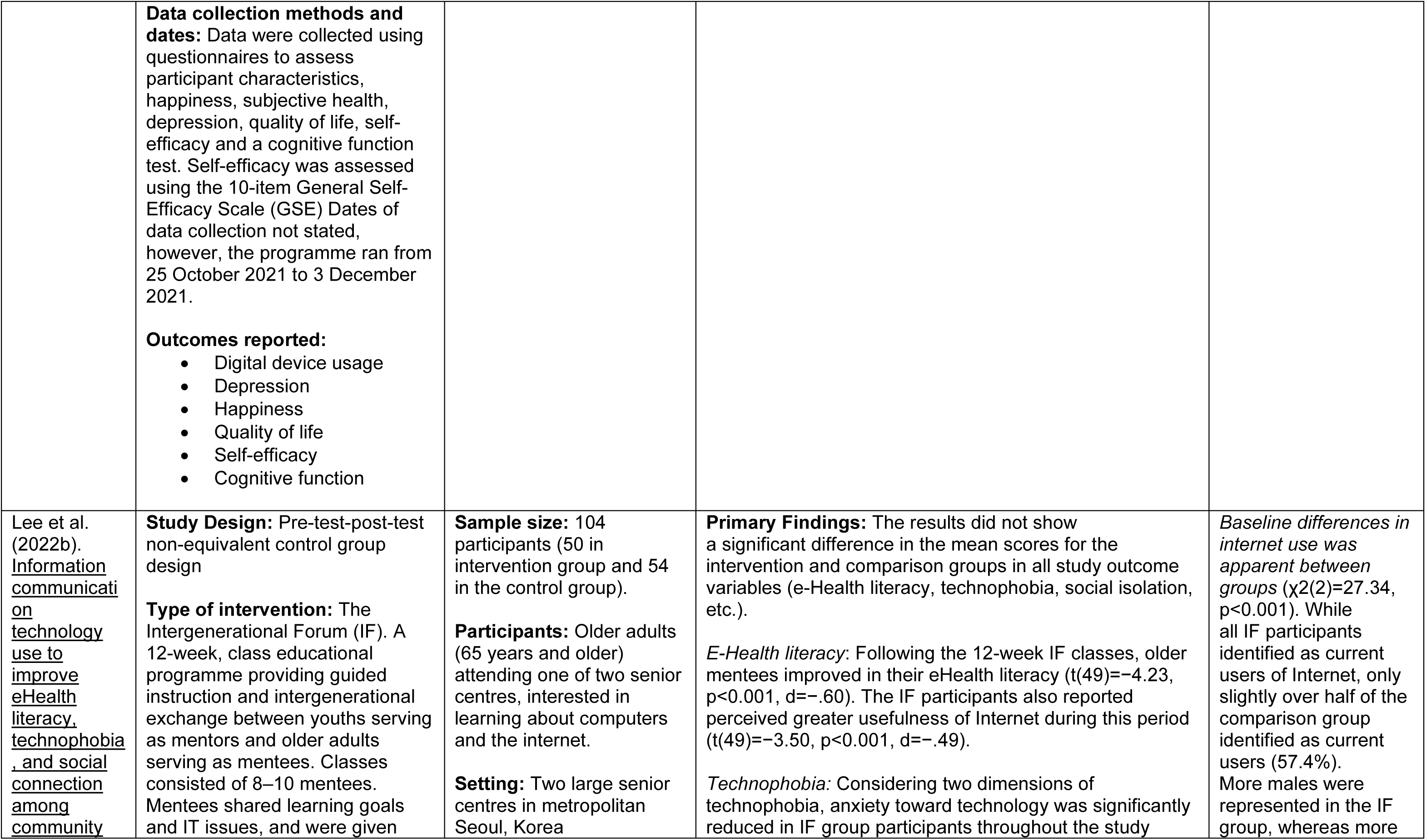
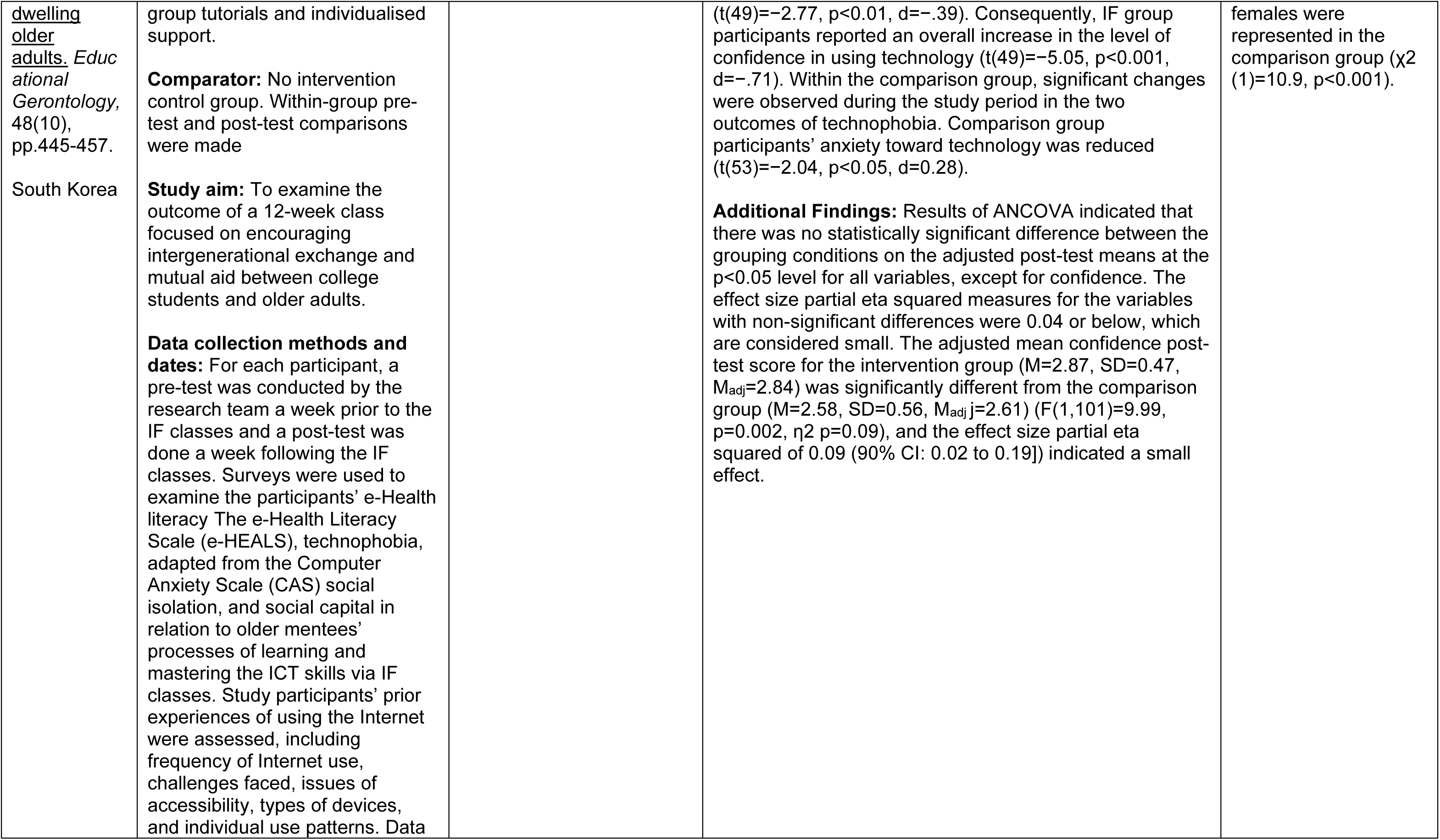
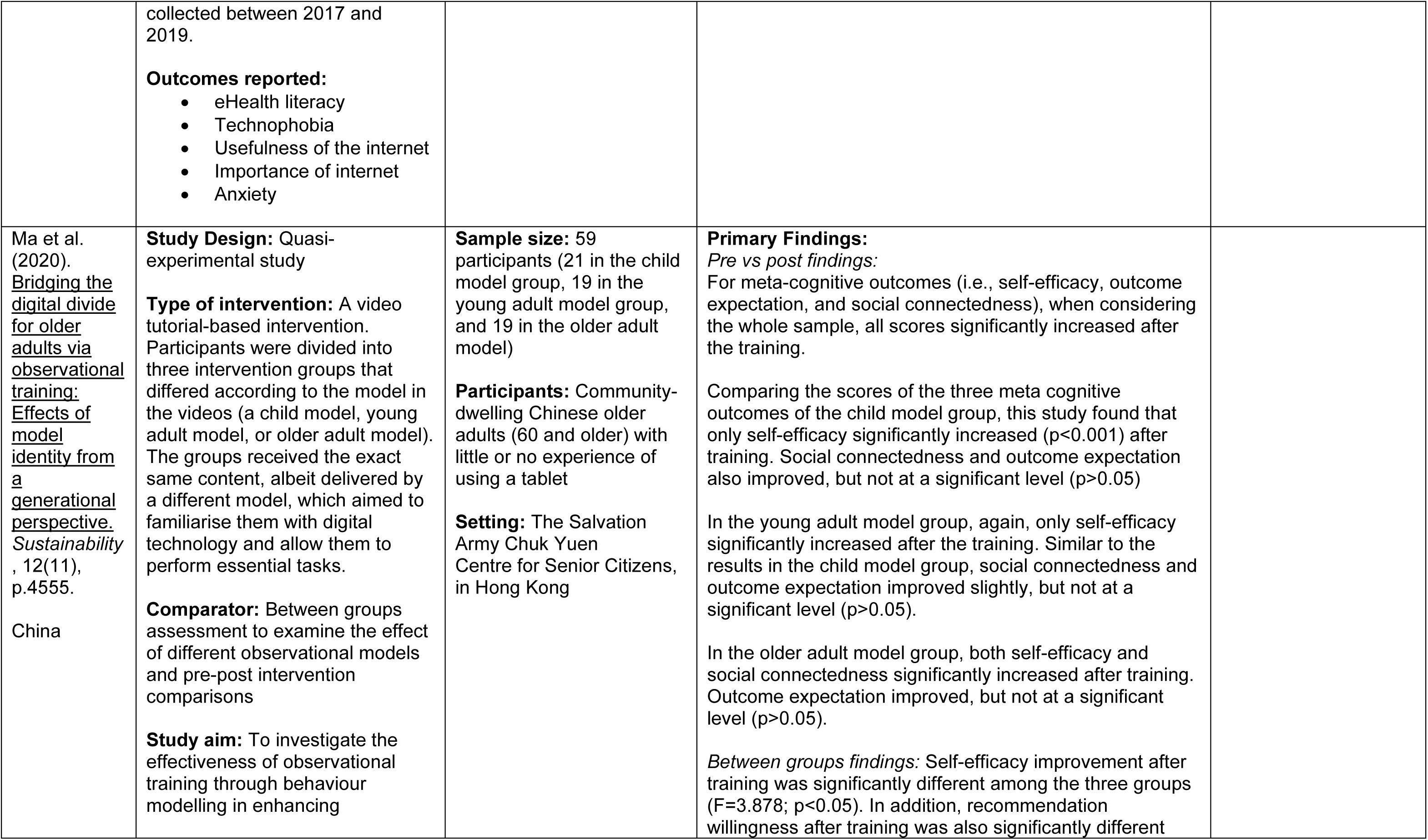
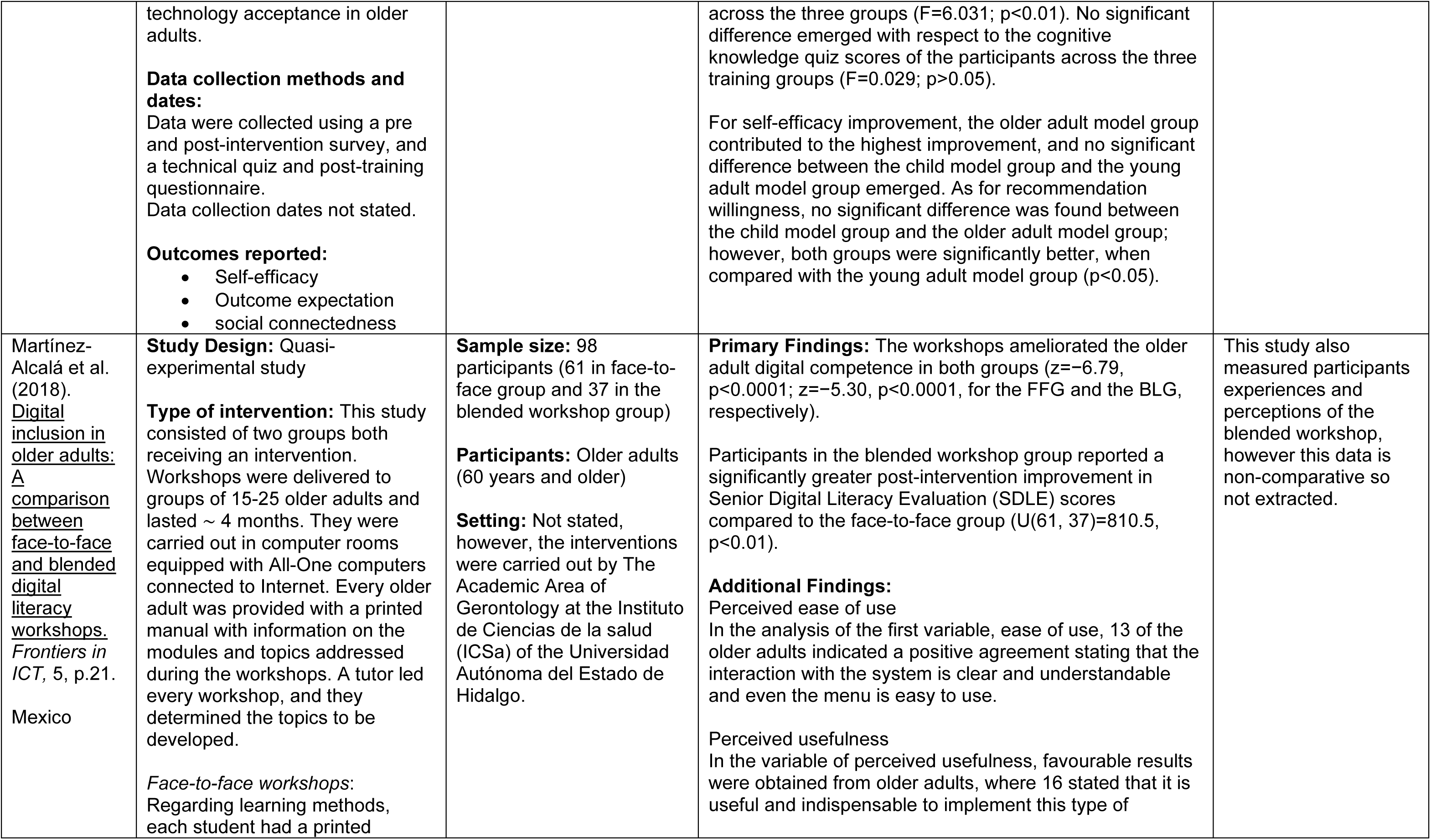
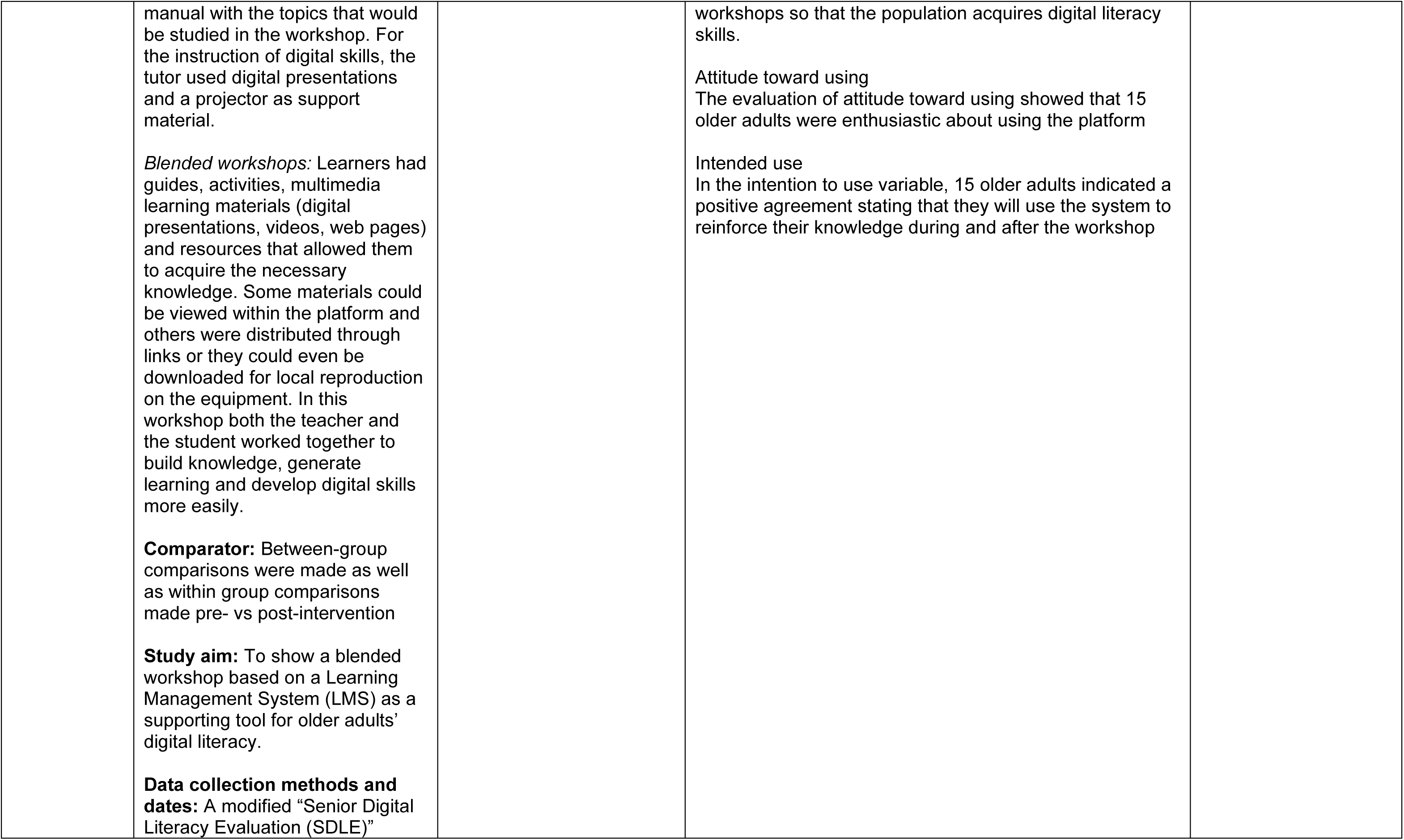
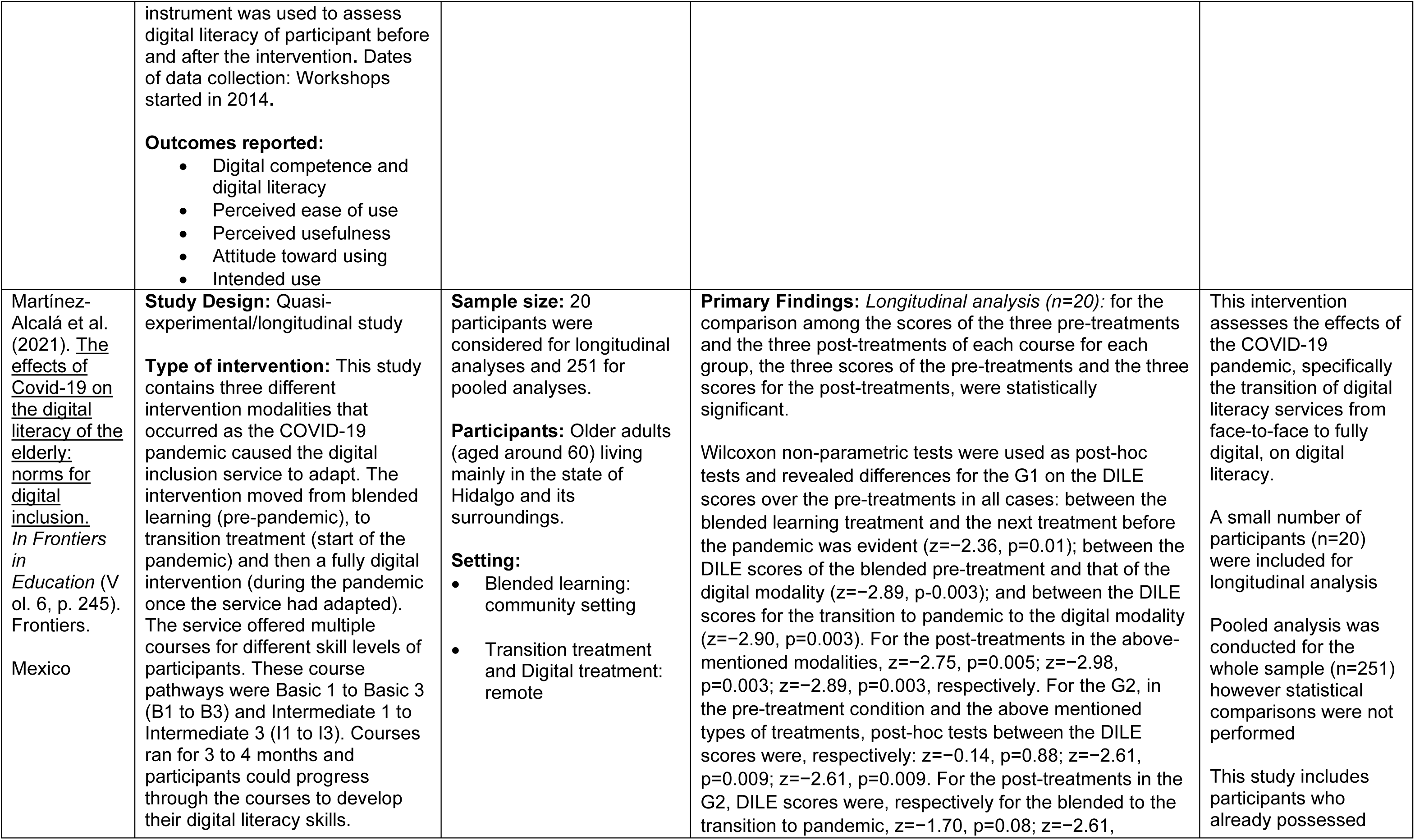
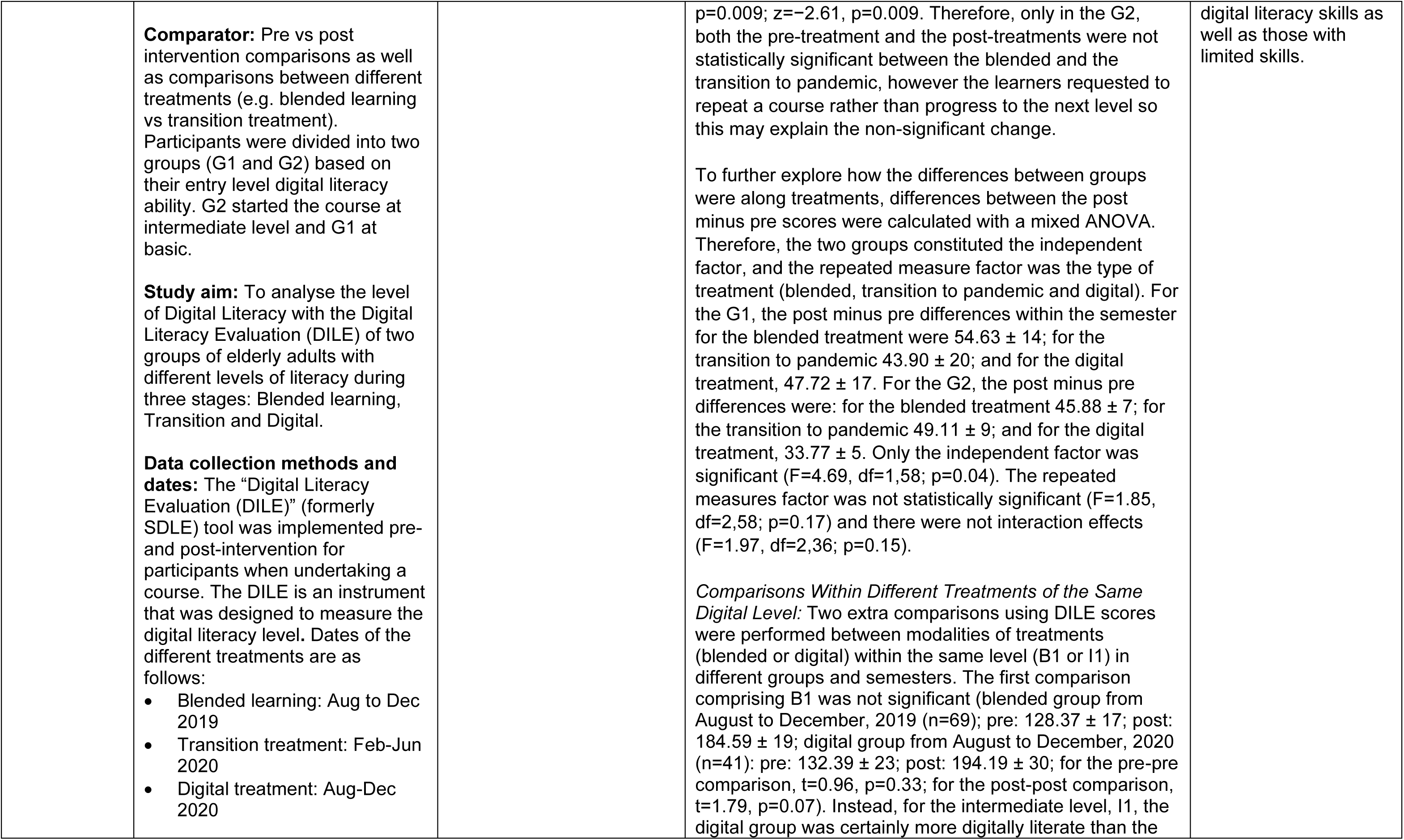
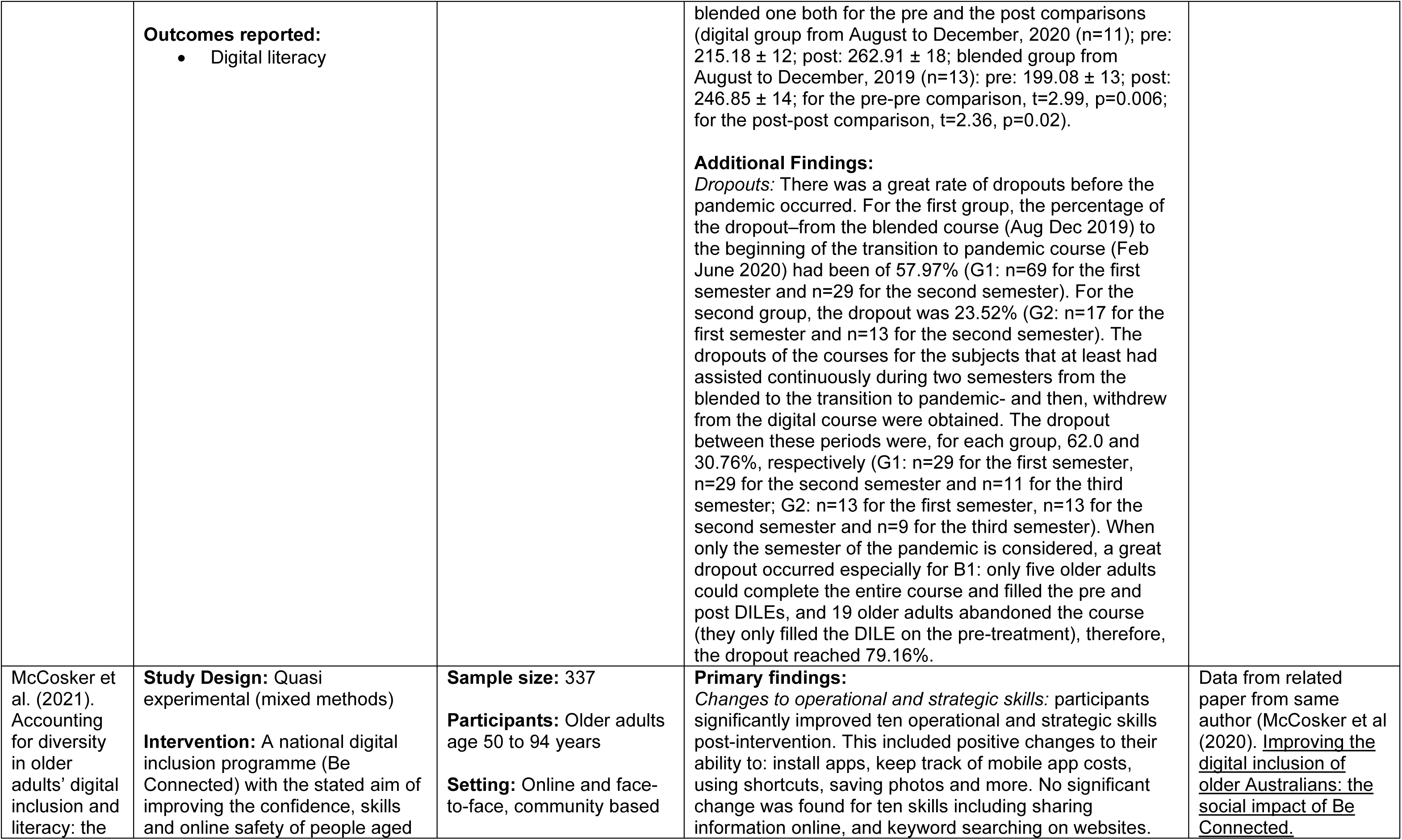
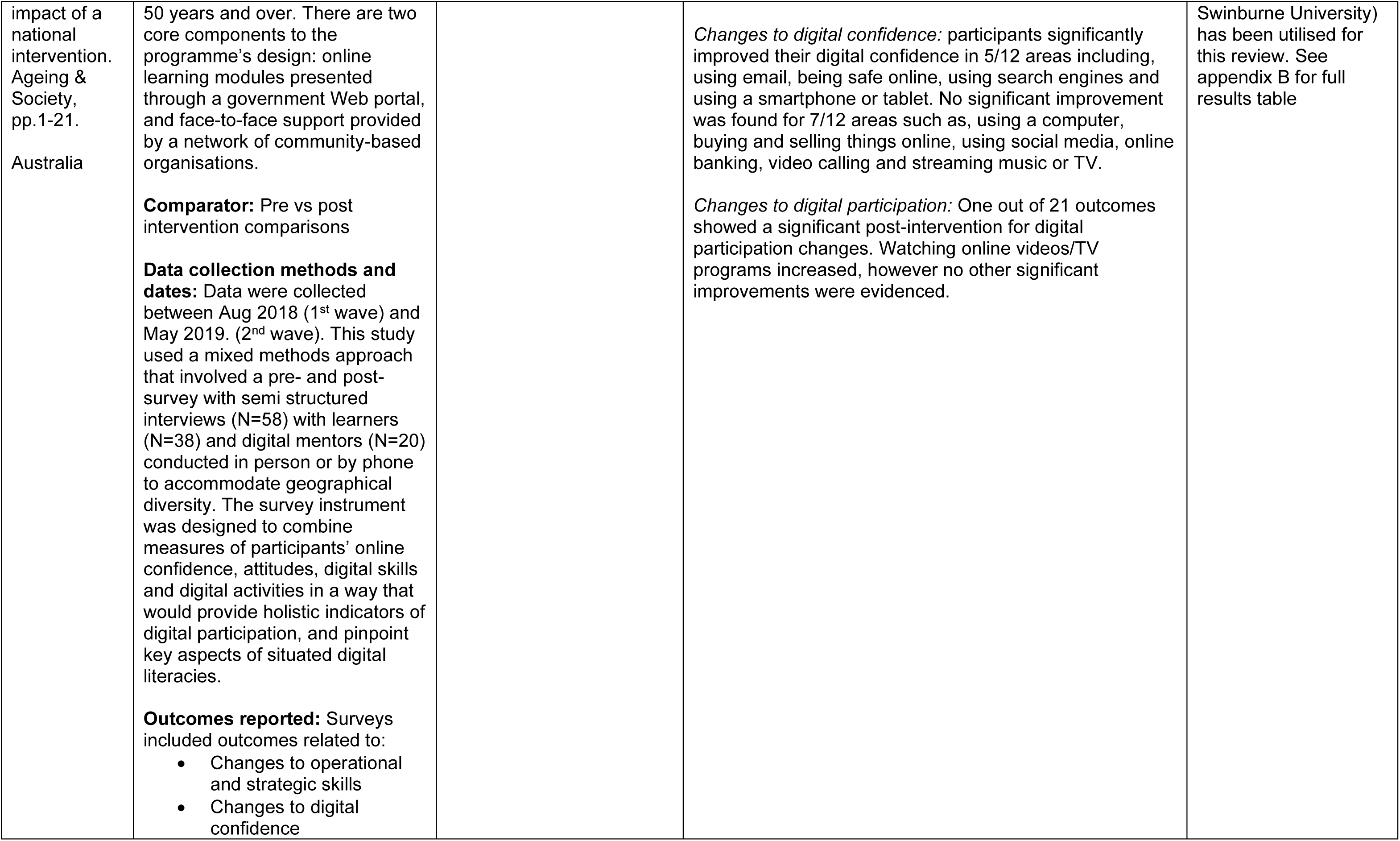
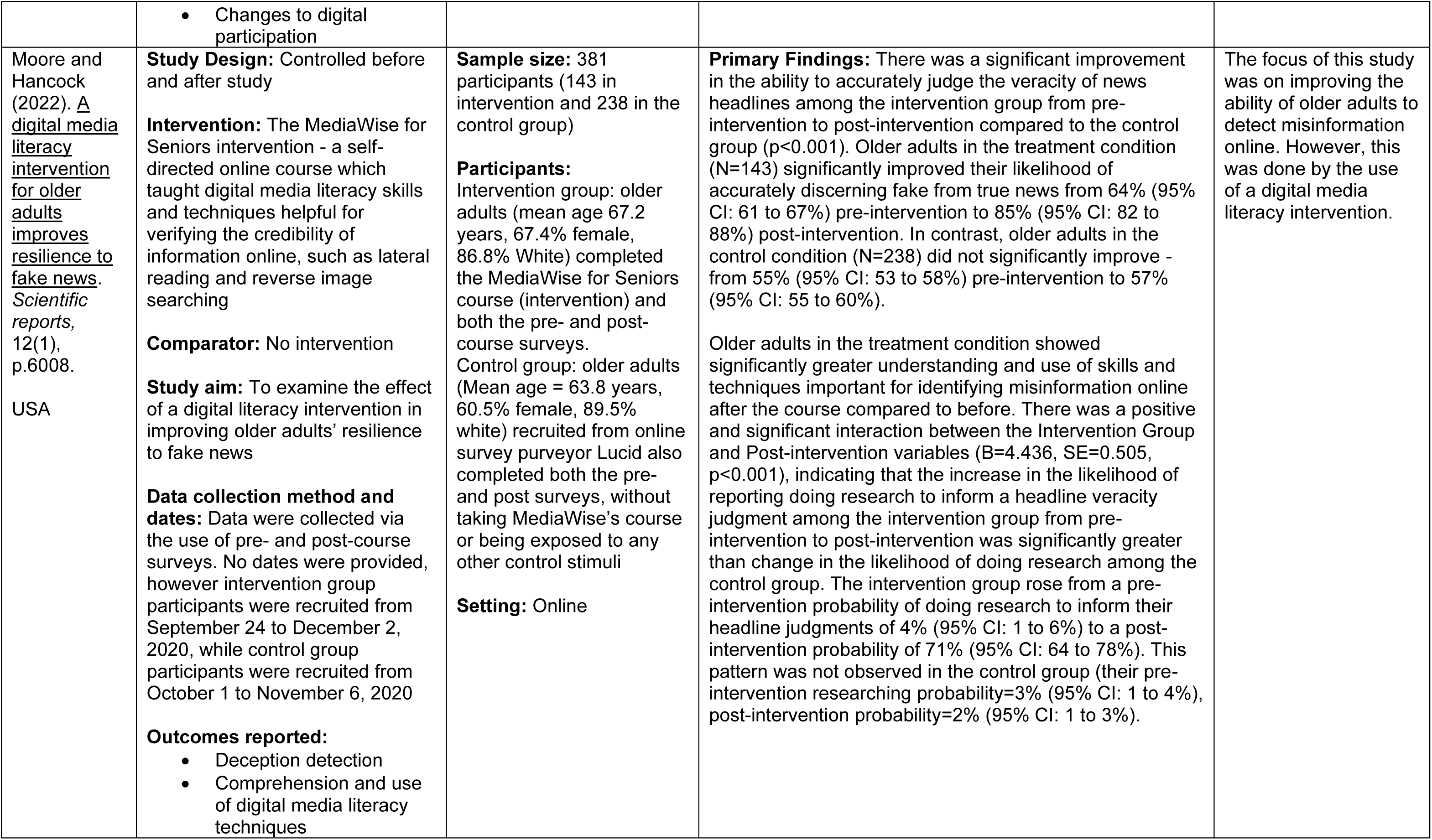
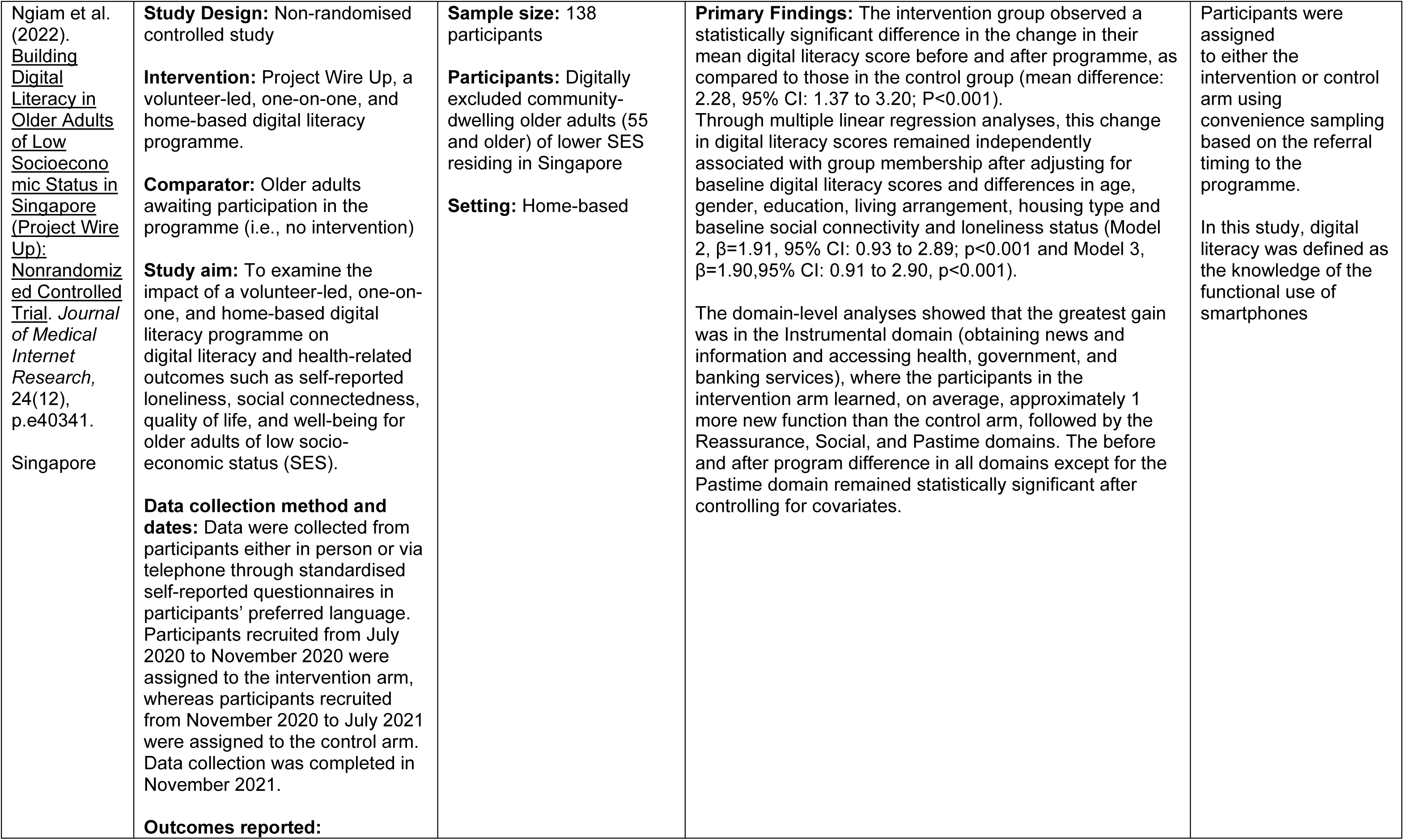
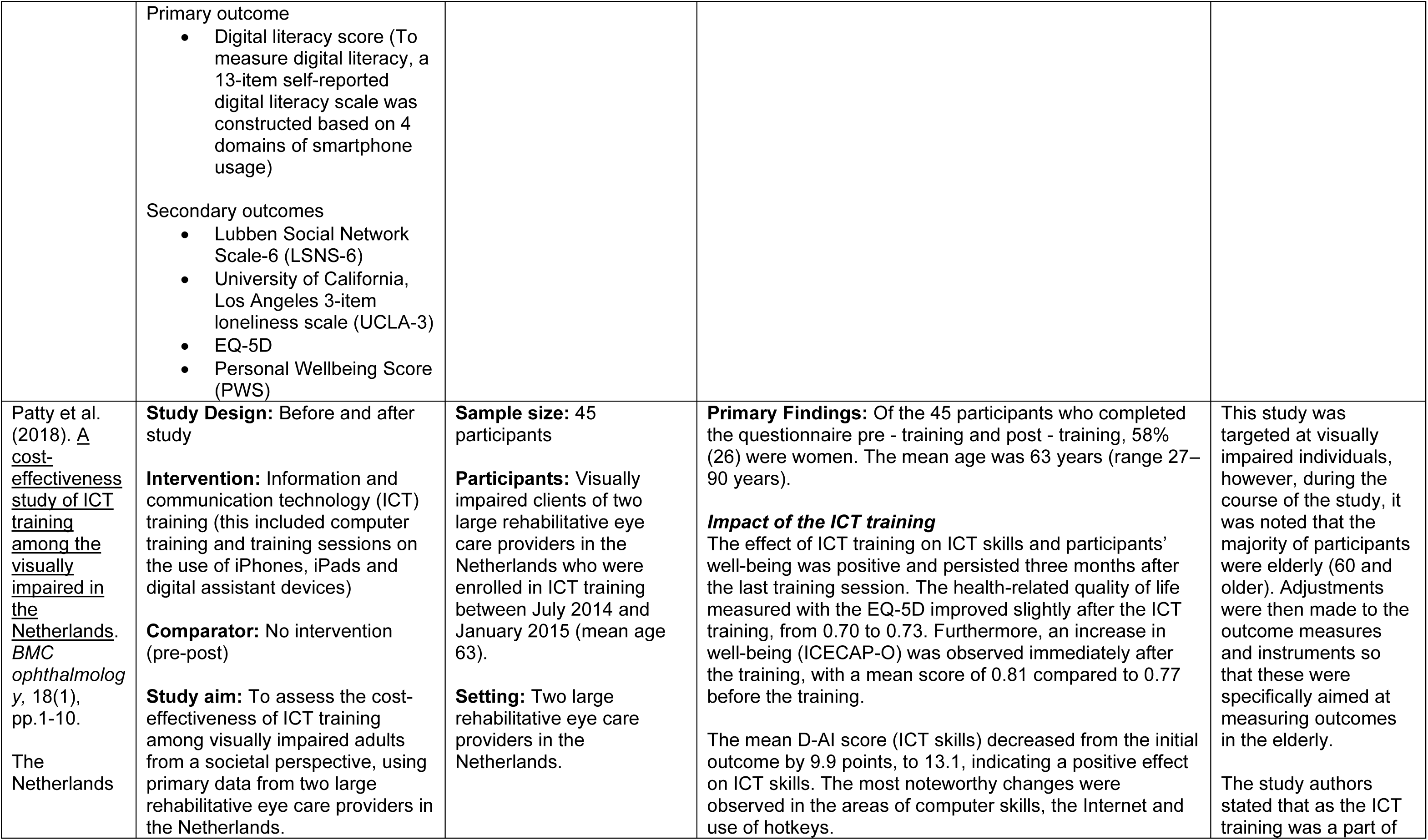
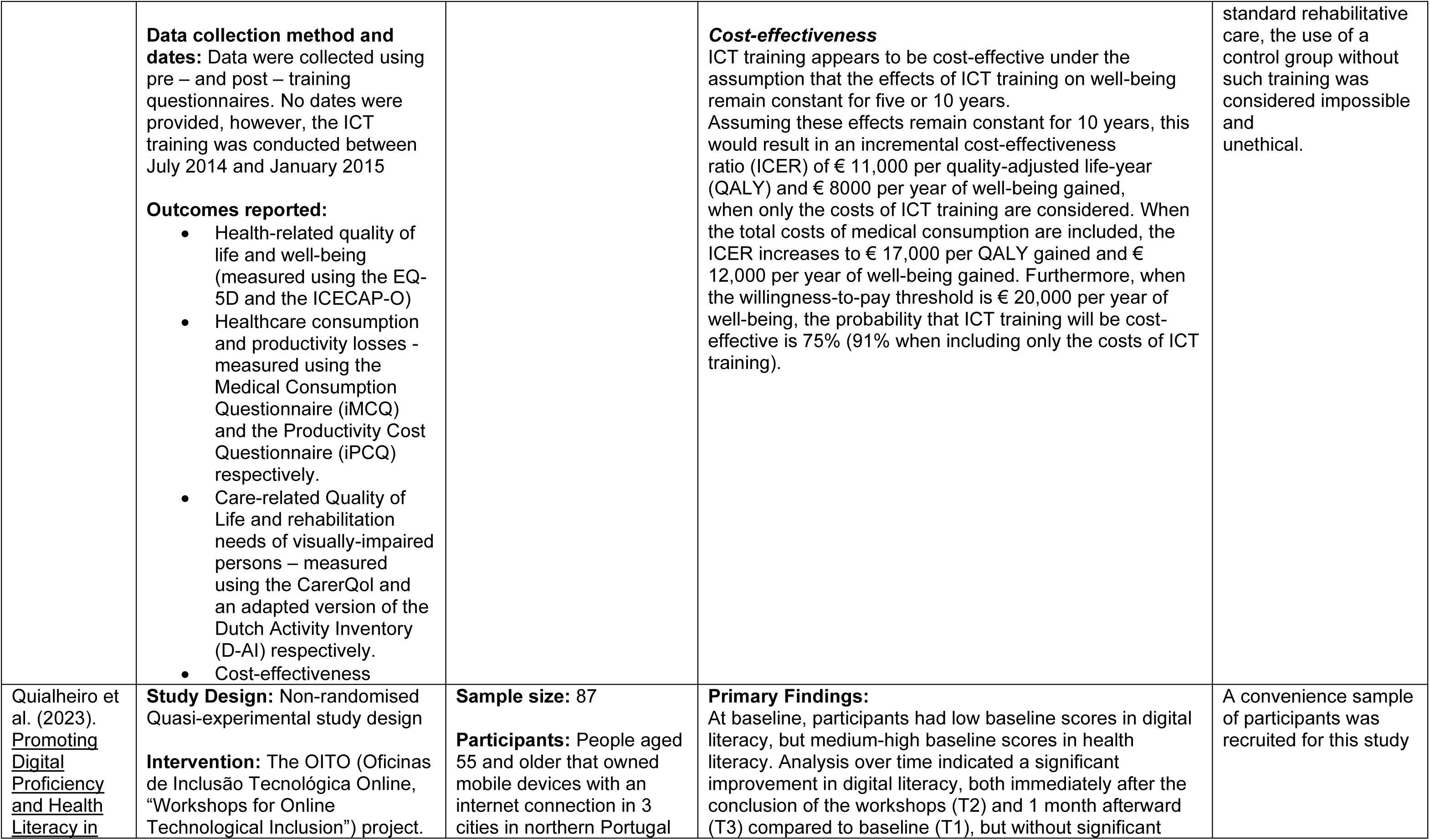
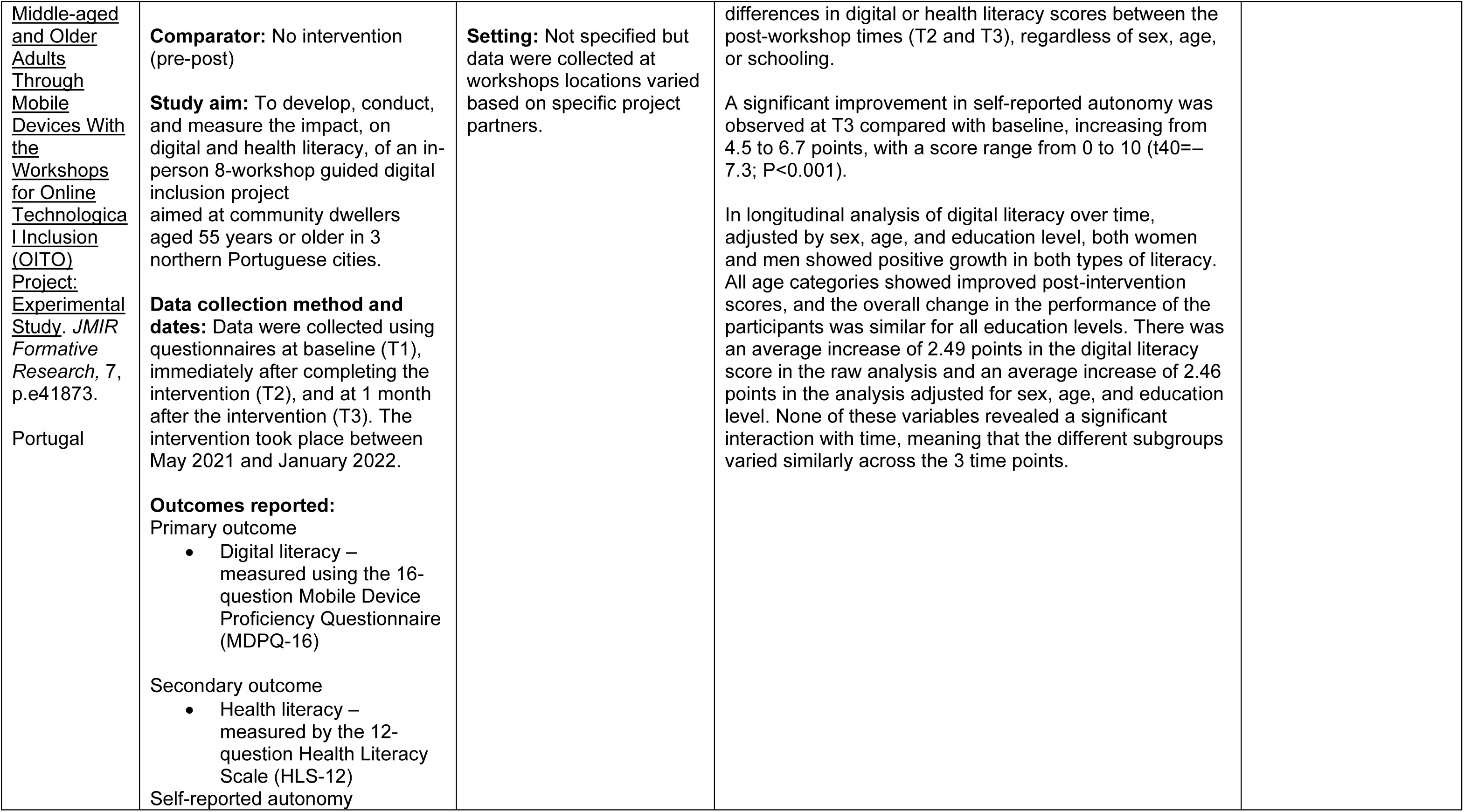
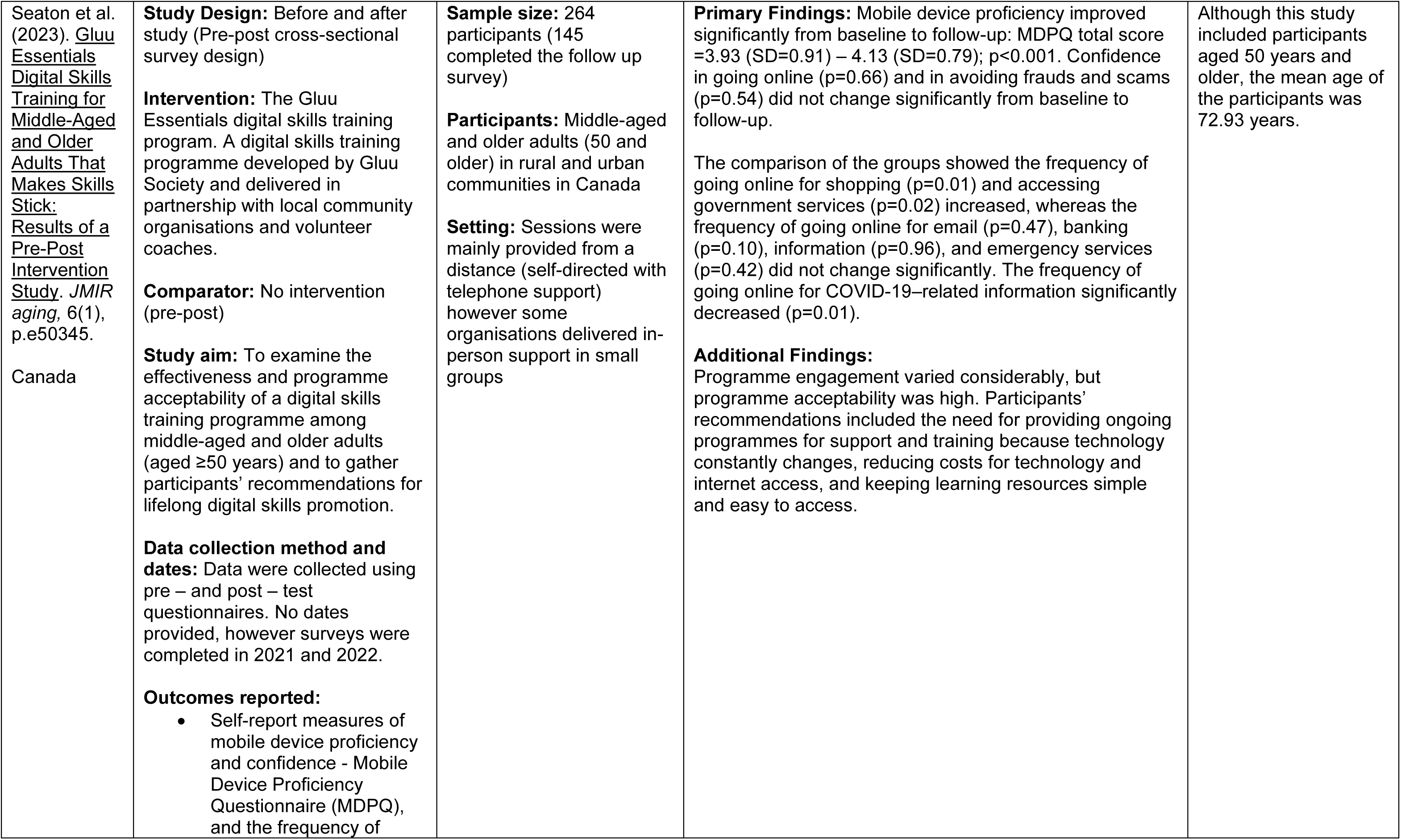
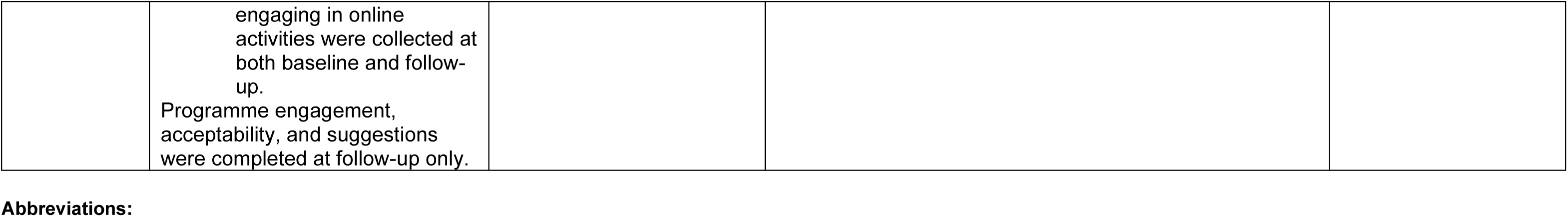
Summary of included studies.

### 6.3 Quality appraisal

**Table 6.** Quality appraisal results for quasi-experimental studies.

| Study | JBI Appraisal Items – Quasi-experimental studies |  |  |  |  |  |  |  |  | Overall quality |
| --- | --- | --- | --- | --- | --- | --- | --- | --- | --- | --- |
|  | Q1. | Q2. | Q3. | Q4. | Q5. | Q6. | Q7. | Q8. | Q9. |  |
| Castilla et al. (2018) | Y | Y | Y | N | N | N/A | Y | U | Y | Low |
| Choi and Park (2022) | Y | U | Y | Y | N | Y | Y | U | Y | Low |
| Elbaz et al. (2023) | Y | Y | Y | N | N | N/A | Y | U | Y | Low |
| Gadbois et al. (2022) | Y | Y | Y | N | N | N/A | Y | U | Y | Low |
| Garcia et al. (2022) | Y | U | Y | Y | N | N | Y | U | Y | Low |
| Holguin-Alvarez et al. (2020) | Y | Y | Y | Y | N | N | Y | U | U | Low |
| Lee and Kim (2019) | Y | Y | Y | N | N | N/A | Y | Y | Y | Low |
| Lee et al. (2022a) | Y | N | Y | Y | N | N | Y | U | Y | Low |
| Lee et al. (2022b) | Y | N | Y | Y | N | N | Y | Y | Y | Low |
| Ma et al. (2020) | Y | Y | Y | N | N | N/A | Y | Y | Y | Low |
| Martínez-Alcalá et al. (2018) | Y | Y | Y | Y | N | N | Y | U | Y | Low |
| Martínez-Alcalá et al. (2021) | Y | U | Y | N | Y | N | Y | U | Y | Low |
| McCosker et al. (2020) | Y | Y | Y | N | N | N/A | Y | U | Y | Low |
| Moore & Hancock (2022) | Y | U | Y | Y | N | Y | Y | U | Y | Low |
| Ngiam et al. (2022) | Y | Y | Y | Y | N | Y | Y | U | Y | Low |
| Patty et al. (2018) | Y | Y | Y | N | Y | N/A | Y | U | Y | Low |
| Quialheiro et al. (2023) | Y | Y | Y | N | Y | N/A | Y | Y | Y | Low |
| Seaton et al. (2023) | Y | Y | Y | N | N | N/A | Y | Y | Y | Low |

Key: Y=Yes, N=No, U=unclear, N/A=not applicable
1. Is it clear what is the cause and what is the effect? 2. Were the participants included in any comparisons similar? 3. Were the participants included in any comparisons receiving similar treatment/care, other than the exposure or intervention of interest? 4. Was there a control group? 5. Were there multiple measurements of the outcome both pre and post the intervention/ exposure? 6. Was follow up complete and if not, were differences between groups in terms of their follow up adequately described and analysed? 7. Were the outcomes of participants included in any comparisons measured in the same way? 8. Were outcomes measured in a reliable way? 9. Was appropriate statistical analysis used?

**Table 7.**
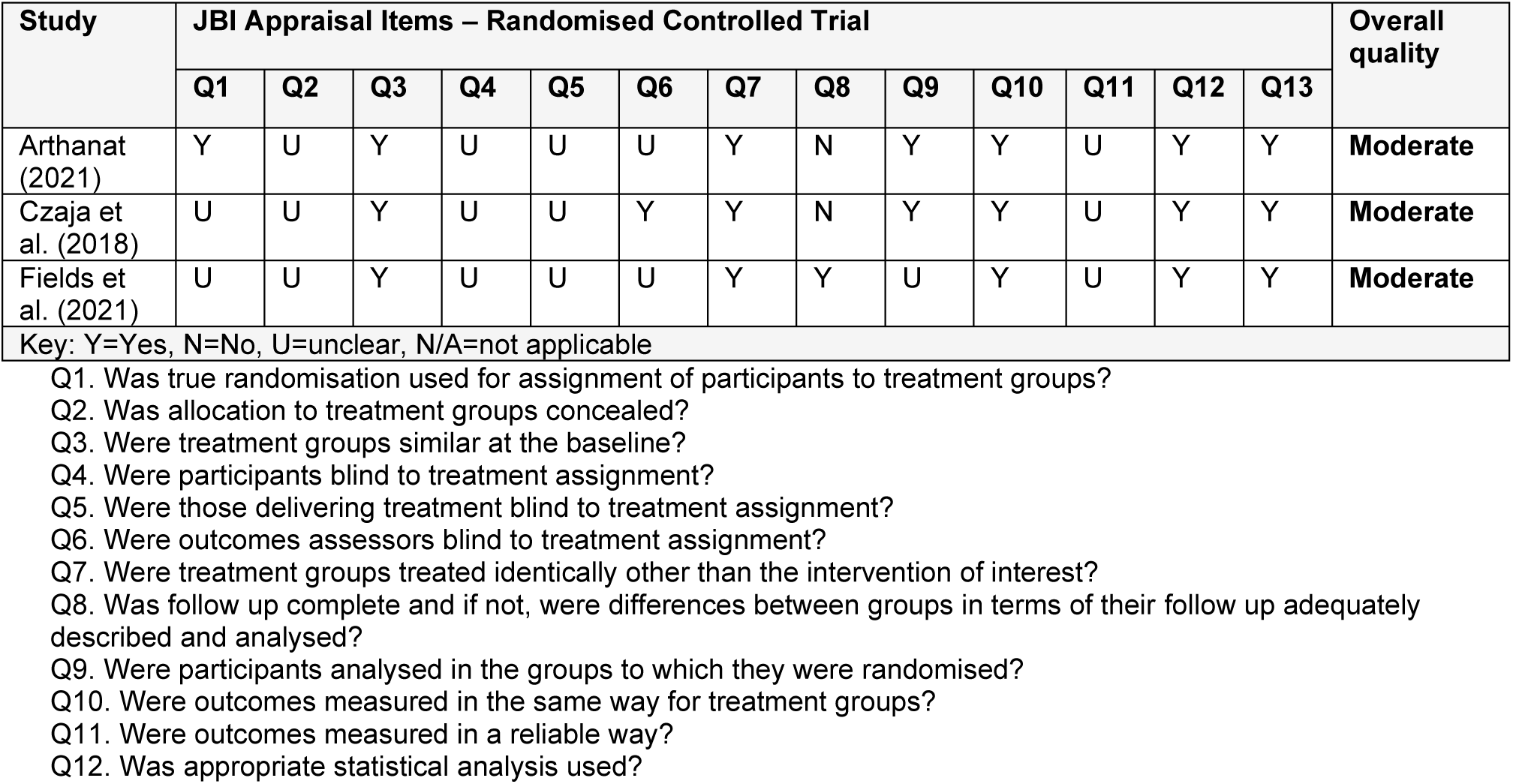
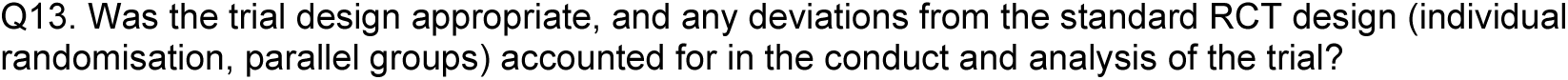
Quality appraisal results for randomised controlled trials.

| Study | JBI Appraisal Items – Randomised Controlled Trial |  |  |  |  |  |  |  |  |  |  |  |  | Overall quality |
| --- | --- | --- | --- | --- | --- | --- | --- | --- | --- | --- | --- | --- | --- | --- |
|  | Q1 | Q2 | Q3 | Q4 | Q5 | Q6 | Q7 | Q8 | Q9 | Q10 | Q11 | Q12 | Q13 |  |
| Arthanat (2021) | Y | U | Y | U | U | U | Y | N | Y | Y | U | Y | Y | Moderate |
| Czaja et al. (2018) | U | U | Y | U | U | Y | Y | N | Y | Y | U | Y | Y | Moderate |
| Fields et al. (2021) | U | U | Y | U | U | U | Y | Y | U | Y | U | Y | Y | Moderate |

Key: Y=Yes, N=No, U=unclear, N/A=not applicable
- Q1. Was true randomisation used for assignment of participants to treatment groups? - Q2. Was allocation to treatment groups concealed? - Q3. Were treatment groups similar at the baseline? - Q4. Were participants blind to treatment assignment? - Q5. Were those delivering treatment blind to treatment assignment? - Q6. Were outcomes assessors blind to treatment assignment? - Q7. Were treatment groups treated identically other than the intervention of interest? - Q8. Was follow up complete and if not, were differences between groups in terms of their follow up adequately described and analysed? - Q9. Were participants analysed in the groups to which they were randomised? - Q10. Were outcomes measured in the same way for treatment groups? - Q11. Were outcomes measured in a reliable way? - Q12. Was appropriate statistical analysis used?
Q13. Was the trial design appropriate, and any deviations from the standard RCT design (individual randomisation, parallel groups) accounted for in the conduct and analysis of the trial?

### 6.4 Information available on request

The following are available on request: protocol; search strategies.

## 7.2 Acknowledgements

The Public Health Wales team would like to thank Aimee Twinberrow, Beti Ingram, Eleanor Johnson, Emma Taylor-Collins, Josie Jackson, Melanie McAulay, Gareth Ashman and Stephen Thomas for their time, expertise and contributions during stakeholder meetings in guiding the focus of the review and interpretation of findings.

## 8. APPENDIX

### APPENDIX 1: Search strategy used for Social Policy and Practice

1. (“Digital exclusion” or “Digital inclusion” or “Digital divide*” or “Digital division*” or “Digital disparit*” or “Digital gap*” or “Digital inequalit*” or “digital training” or “Technostress” or “Digital literacy” or “Digital skills” or “Computer literacy” or “ICT literacy” or “computer skills” or “ICT skills” or “information and communicat* technolog*”).ti,ab. 1696
2. (elder* or old* or ageing or aging or senior or seniors or carer* or geriatric* or Centarian* or centenarian* or eldest or frail* or geriatri* or nonagenarian* or octagenarian* or octogenarian* or “old age*” or “older adult*” or “older age*” or “older patient*” or “older people” or “older person*” or “older population” or “older subject*” or oldest or senium or septuagenarian* or supercentenarian* or “older female*” or “older male*” or “older man” or “older men” or “older woman” or “older women”).ti,ab. 95831
3. 1 and 2 486
4. limit 3 to yr=“2018 - 2024” 165

